# Beyond the Percentile: CDC-referenced BMI-for-Age and Brain Structure Across Childhood and Adolescence

**DOI:** 10.64898/2026.10.18.26365816

**Authors:** Dani Beck, Lars T. Westlye, Christian K. Tamnes

**Author notes:** **Corresponding author**: Dani Beck. Co-author.

## Abstract

**Importance:** Pediatric obesity affects 1 in 5 US children and has been linked to brain structural differences. Most evidence is cross-sectional and models body mass index (BMI) linearly, with metrics that compress severe obesity.

**Objective:** To examine associations between brain structure and BMI-for-age across the weight spectrum and Centers for Disease Control and Prevention (CDC) weight status categories, distinguishing between- and within-person associations, and their age and sex moderation.

**Design, Setting, and Participants:** Longitudinal cohort study of youth aged 8–10 years at baseline in the US community-based Adolescent Brain Cognitive Development Study (4 waves, 2016–2025; ages 8–18 years).

**Exposures:** Extended CDC BMI-for-age z-score, grouped by wave-mean z-score: underweight (<5th percentile, n=297), healthy weight (5th–<85th, n=7,452), overweight (85th–<95th, n=1,919), and obesity (≥95th, n=1,842).

**Main Outcomes and Measures:** Cortical thickness (12 regions) and subcortical volume (7 regions) from T1-weighted magnetic resonance imaging (MRI), defined a priori.

**Results:** Among 11,510 participants contributing 32,212 observations (mean [SD] age, 12.6 [2.5] years; 52.2% male), higher between-person BMI-for-age z was associated with lower frontal and temporal thickness (6 regions) and larger subcortical volume (5 regions; all *q* < .001), but associations were nonlinear (17 of 19 regions). Youth with obesity had lower thickness than healthy weight youth in 8 frontal and temporal regions (largest, −0.21 SD; *q* < .001), with dorsal frontal differences decreasing across adolescence; underweight youth had smaller subcortical volumes. Higher BMI-for-age z was associated with larger subcortical volume (all 7 regions) within the healthy weight category and lower cortical thickness (11 of 12 regions; fusiform: β = −0.27; *q* < .001) within the obesity category, steeper than in healthy weight youth in 10 regions, but not in overweight or underweight youth. Within-person associations did not reproduce the obesity associations, reversed from negative at age 10 to positive at age 16, and were more negative in female than male participants.

**Conclusions and Relevance:** Youth with obesity had lower cortical thickness than healthy weight peers, but associations between brain structure and BMI-for-age differed between CDC weight status groups, suggesting a complex relationship between adiposity and brain structure.

**Key Points:** *Question:* How is BMI-for-age associated with brain structure across the weight spectrum, within CDC weight status categories, and within individuals over time in youth?

*Findings:* In this cohort study of 11,510 youth aged 8–18 years, those with obesity had lower frontal and temporal cortical thickness than healthy weight youth. Within categories, higher BMI-for-age was associated with lower thickness in obesity but larger subcortical volumes in healthy weight youth; within individuals over time, the obesity association was not reproduced.

*Meaning:* BMI–brain associations differ between CDC weight status categories and may be obscured by assuming a single linear association.

## Introduction

Pediatric obesity affects about 1 in 5 US children and adolescents^1,2^ and often persists into adulthood^3,4^. Beyond cardiometabolic risk, childhood obesity is associated with attention-deficit/hyperactivity disorder, depression, learning disabilities, and functional impairment^5,6^, with at least partly causal links to externalizing and internalizing symptoms^7^.

Higher body mass index (BMI) is also linked to differences in brain structure. In adults, higher adiposity and cardiometabolic risk are associated with smaller frontal and temporal gray matter volume, white matter differences, and older-appearing brains^8–13^. In youth, cross-sectional studies in the Adolescent Brain Cognitive Development (ABCD) Study^14,15^ have linked higher BMI to lower prefrontal and temporal cortical thickness at ages 9 to 10 years^16–18^, and independent pediatric samples report positive associations with accumbens, pallidum, and amygdala volume^19–21^. This fronto-temporal pattern extends from early adolescence to older adulthood^22^.

Yet most pediatric evidence is cross-sectional, leaving unclear whether a child’s brain changes as their own BMI changes; heavier children may differ in brain structure because of pre-existing neural differences^23^ or genetic or prenatal factors that shape both brain development and adiposity^24–26^, confounds that cross-sectional designs cannot resolve^27,28^. Most studies also model BMI as a single linear predictor^16–18^, which implies that a given BMI difference carries the same meaning in the healthy weight range as in obesity. Whether BMI–brain associations shift with age or differ by sex is also poorly understood, despite sex-by-obesity interactions on brain structure in adults^29,30^. Finally, raw BMI rises with growth and conventional BMI-for-age z-scores compress the upper tail where severe obesity falls^31,32^. The extended Centers for Disease Control and Prevention (CDC) BMI-for-age z-score^33^ preserves upper-tail differentiation and maps onto the 4 official CDC weight status categories, providing a clinically anchored framework for these questions.

Here, in 11,510 ABCD participants assessed up to 4 times between ages 8 and 18 years (Figure 1), we examined whether BMI-for-age is linearly associated with cortical thickness and subcortical volume across the weight spectrum; whether brain structure and its development differ between the 4 CDC weight status categories; and, within each category, how between- and within-person associations compare and whether within-person associations vary with age and sex.

**Figure 1.**
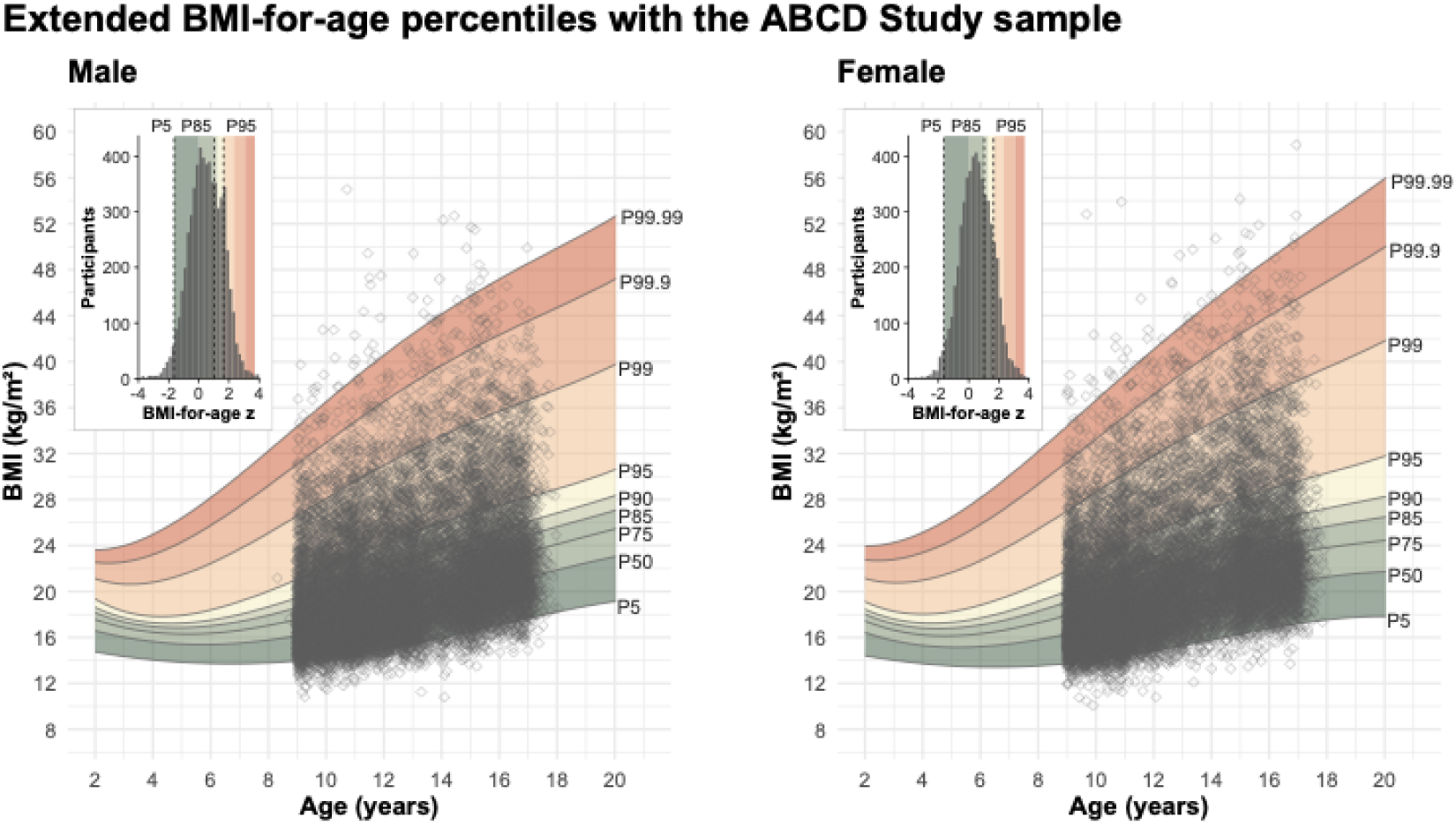
Extended CDC BMI-for-age percentiles with the ABCD sample. Body mass index (kg/m²) against age for male (left) and female (right) participants. Shaded bands denote CDC extended BMI-for-age percentile regions (5th to 99.99th percentile). Gray diamonds are individual observations across all waves. Insets show the distribution of participants’ wave-mean extended BMI-for-age z, the value used to assign CDC weight status groups (bin width, 0.2; shading as in the main panels). Dashed lines mark the group cut-offs at the 5th, 85th, and 95th percentiles (z = −1.645, 1.036, and 1.645), separating underweight, healthy weight, overweight, and obesity. Insets show z from −4 to 4; 23 participants with values outside this range are not shown.

## Methods

### Study Design, Setting, and Participants

This longitudinal cohort study used release 7.0 of the ABCD Study^14^ (2016–2025; NBDC Data Hub), a community-based sample of US children recruited at ages 8 to 10 years at 21 sites (variable sources in eMethods 1 in the Supplement). The study followed the STROBE guideline and received central institutional review board approval at the University of California, San Diego^34^, and Norwegian ethical approval (REK 2019/943); parents provided consent and children assent. Across 4 imaging waves (baseline and 2-, 4-, and 6-year follow-up), 33,456 observations from 11,794 participants had a T1-weighted magnetic resonance imaging (MRI) scan passing ABCD quality control^14^. The analytic sample comprised 32,212 observations from 11,510 participants with complete data on BMI-for-age z, covariates, family identifier, and outcomes (sample flow in eMethods 2; missingness in eFigure 1).

### MRI Acquisition and Processing

T1-weighted images were acquired on 3T scanners using a harmonized protocol^14,35^ and processed with FreeSurfer v7.1.1^36,37^. We used bilateral averages of Desikan–Killiany cortical thickness and subcortical volume. Scanner effects across 34 scanners were adjusted for with longitudinal ComBat^38^, preserving variance associated with age, sex, wave, raw BMI, ethnicity, and socioeconomic status (eMethods 2 and 3; eFigures 2 and 3).

**Figure 2.**
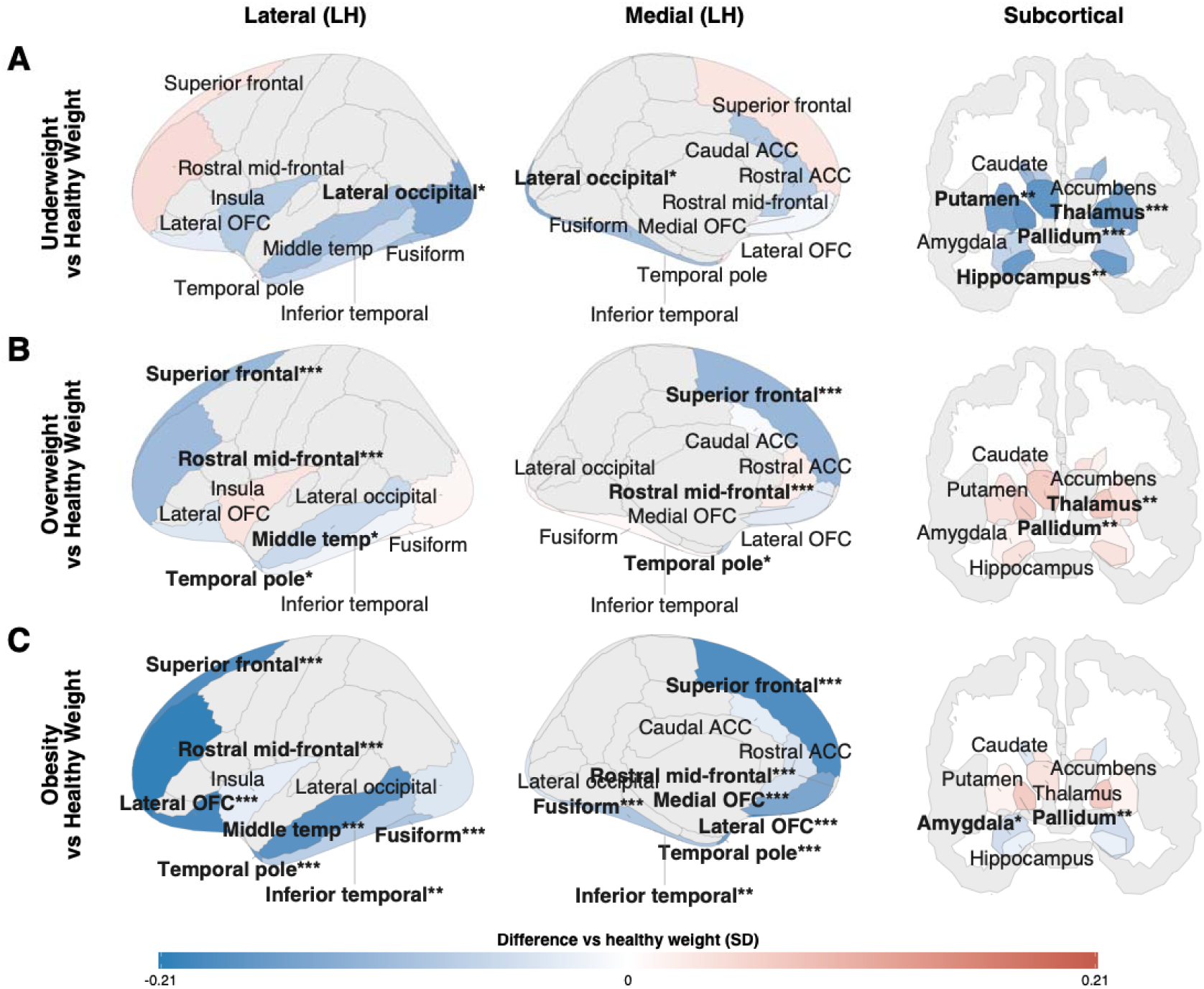
Differences in regional brain structure between CDC weight status groups. Adjusted differences in cortical thickness and subcortical volume between each group and the healthy weight group (5th to <85th percentile), in SD units, mapped to left-hemisphere cortical surfaces (lateral and medial views) and a subcortical coronal section: (A) underweight (<5th percentile); (B) overweight (85th to <95th percentile); (C) obesity (≥95th percentile). Groups were defined by each participant’s mean extended BMI-for-age z across included visits, and differences were estimated across all participants, adjusted for age, age², sex, pubertal development, socioeconomic status, race and ethnicity, and intracranial volume (subcortical). Blue indicates lower and red higher cortical thickness or subcortical volume than in the healthy weight group. All 19 analyzed regions are colored by the estimated difference; bold labels with asterisks denote FDR-significant differences (\**q* < .05, \*\**q* < .01, \*\*\**q* < .001), and regions not analyzed are shown in gray. The color scale is shared across panels (−0.21 to 0.21). Youth with obesity show lower thickness in 8 frontal and temporal regions, most markedly in rostral middle frontal cortex, larger pallidum volume, and smaller amygdala volume (C); overweight youth show lower thickness in 4 of these regions and larger thalamus and pallidum volume (B); and underweight youth show smaller volume in 4 subcortical regions and lower lateral occipital thickness (A). ACC indicates anterior cingulate cortex; FDR, false discovery rate; LH, left hemisphere; OFC, orbitofrontal cortex.

**Figure 3.**
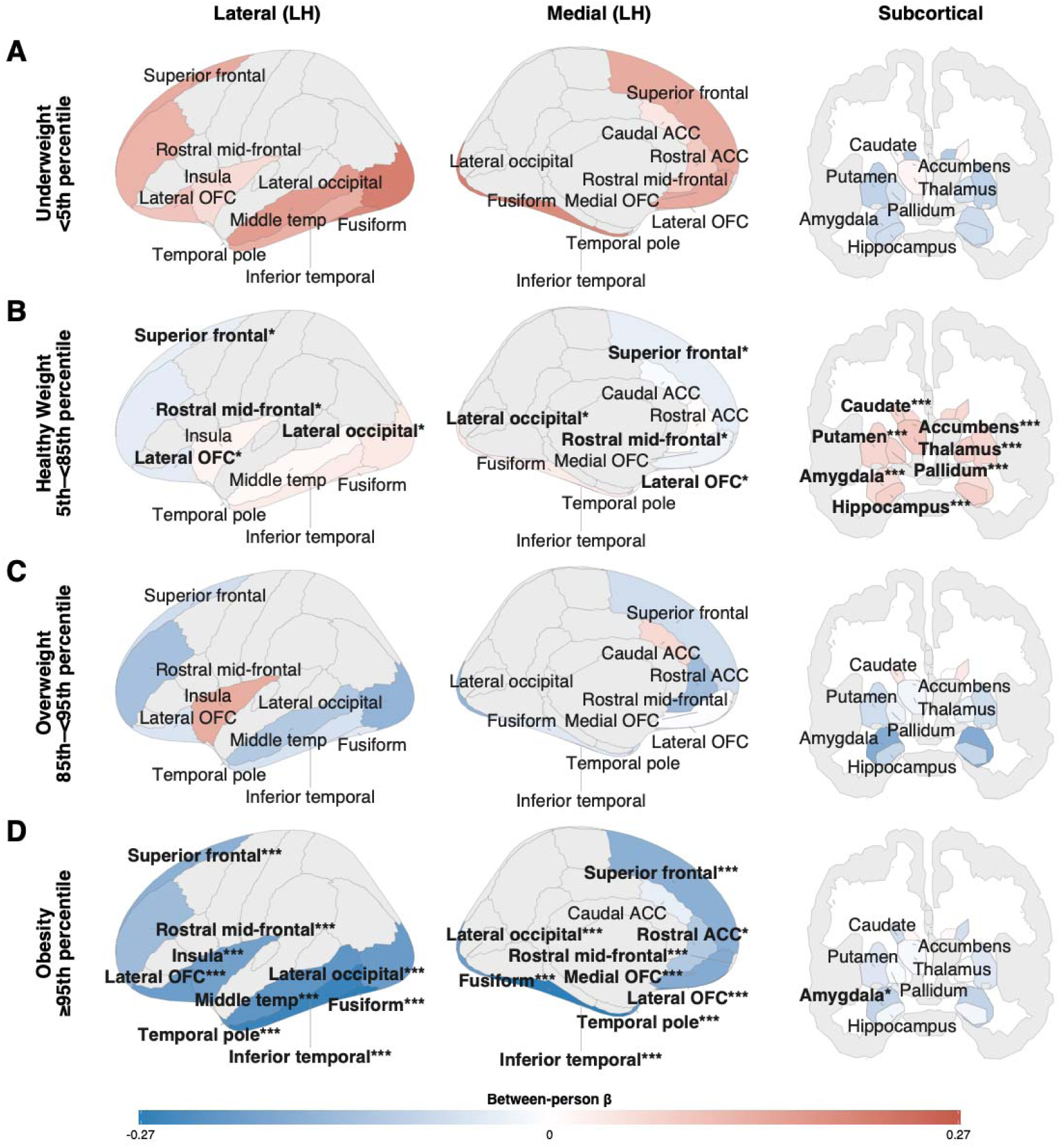
Cross-sectional (between-person) associations of extended BMI-for-age z with regional brain structure by CDC weight status group. Standardized β per 1-SD increase in extended BMI-for-age z mapped to left-hemisphere cortical surfaces (lateral and medial views) and a subcortical coronal section, separately for each group: (A) underweight (<5th percentile); (B) healthy weight (5th to <85th percentile); (C) overweight (85th to <95th percentile); (D) obesity (≥95th percentile). Models were adjusted for age, age², sex, pubertal development, socioeconomic status, race and ethnicity, and intracranial volume (subcortical). Blue indicates lower cortical thickness or subcortical volume at higher between-person BMI- for-age z; red indicates higher. All 19 analyzed regions are colored by β; bold labels with asterisks denote FDR-significant associations (\**q* < .05, \*\**q* < .01, \*\*\**q* < .001), and regions not analyzed are shown in gray. The color scale is shared across groups (−0.27 to 0.27). In healthy weight youth, higher BMI-for-age z is associated with larger volume in all 7 subcortical regions and with small, mixed-direction cortical associations in 4 regions (B); in youth with obesity, it is associated with lower thickness in 11 of 12 cortical regions and smaller amygdala volume (D). The overweight and underweight groups show no significant associations (A, C). ACC indicates anterior cingulate cortex; FDR, false discovery rate; LH, left hemisphere; OFC, orbitofrontal cortex.

### Study Measures

#### Exposure

The extended CDC BMI-for-age z-score was derived from measured height and weight with the cdcanthro R package^39^, relative to the CDC growth reference rather than the ABCD sample; unlike conventional z-scores, it preserves differentiation in severe obesity (eFigure 4).

**Figure 4.**
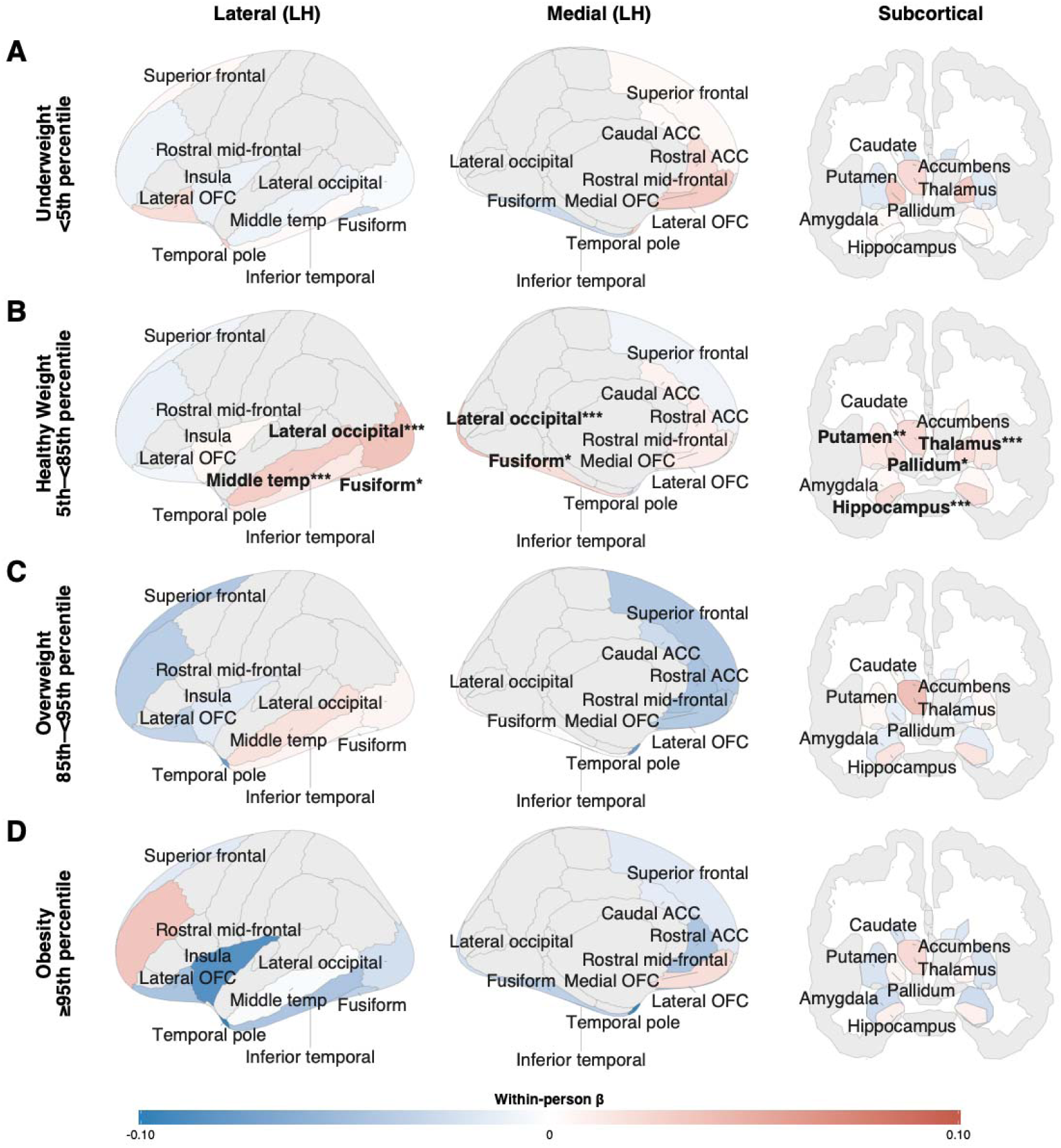
Within-person (longitudinal) associations of extended BMI-for-age z with regional brain structure by CDC weight status group. Standardized β per 1-SD increase in extended BMI-for-age z mapped to left-hemisphere cortical surfaces (lateral and medial views) and a subcortical coronal section, separately for each group: (A) underweight (<5th percentile); (B) healthy weight (5th to <85th percentile); (C) overweight (85th to <95th percentile); (D) obesity (≥95th percentile). Models were adjusted for age, age², sex, pubertal development, socioeconomic status, race and ethnicity, and intracranial volume (subcortical). Within-person associations reflect change within each participant across visits; because each participant serves as their own control, they are free of stable between-person confounding. Blue indicates lower cortical thickness or subcortical volume at visits when a youth’s BMI- for-age z was above their own mean; red indicates higher. All 19 analyzed regions are colored by β; bold labels with asterisks denote FDR-significant associations (\**q* < .05, \*\**q* < .01, \*\*\**q* < .001), and regions not analyzed are shown in gray. The color scale is shared across groups (−0.10 to 0.10). Healthy weight youth show positive within-person associations in 3 cortical regions (lateral occipital, middle temporal, and fusiform) and 4 subcortical regions (thalamus, hippocampus, pallidum, and putamen) (B); no within-person association survived FDR correction in the underweight, overweight, or obesity groups (A, C, D). ACC indicates anterior cingulate cortex; FDR, false discovery rate; LH, left hemisphere; OFC, orbitofrontal cortex.

#### Outcomes

Nineteen bilateral brain regions were selected a priori (rationale in eMethods 2): thickness of 12 cortical regions (medial and lateral orbitofrontal, rostral and caudal anterior cingulate, superior frontal, rostral middle frontal, temporal pole, insula, fusiform, inferior temporal, middle temporal, and lateral occipital cortex) and volume of 7 subcortical structures (nucleus accumbens, amygdala, caudate, hippocampus, pallidum, putamen, and thalamus). Outcomes and continuous covariates were standardized in the analytic sample, so coefficients express SD change in the outcome per 1-unit increase in BMI-for-age z (1 SD of the CDC reference), and group differences are in outcome SD units.

#### Covariates

Models included age (linear and quadratic), sex, pubertal status (parent-reported Pubertal Development Scale), a socioeconomic status composite (mean of parental education and household income), and caregiver-reported race and ethnicity (White, Hispanic, Black, Asian, Other; included to account for sociodemographic differences in BMI), plus estimated intracranial volume for subcortical outcomes (correlations in eFigure 5; sample characteristics in eFigures 6–9).

### Statistical Analysis

Analyses used R version 4.6.1^40^ and nlme^41^ for linear mixed-effects models with nested random intercepts for family and participant, Wald tests for fixed effects, and likelihood ratio tests for nested models (estimation details in eMethods 4). Within each analysis and group, P values were corrected across 19 regions with the Benjamini–Hochberg false discovery rate (FDR) procedure^42^; FDR-corrected P values (q) below .05 were considered significant. Brain maps were rendered with ggseg^43^.

Participants were assigned to the 4 CDC categories by their wave-mean extended BMI-for-age z across included visits (Figure 1): underweight (z < −1.645; <5th percentile), healthy weight (−1.645 to <1.036; 5th to <85th), overweight (1.036 to <1.645; 85th to <95th), and obesity (≥1.645; ≥95th). Within-between (hybrid) models decomposed BMI-for-age z into the wave-mean (between-person component) and each visit’s deviation from it (within-person component). The between-person coefficient compares youth whose typical BMI-for-age z differs; the within-person coefficient indicates whether brain measures were higher at visits when BMI-for-age z was above that youth’s own average and is free of stable confounding.

Analyses proceeded in 4 steps. First, between- and within-person associations and their difference (contextual effect) were estimated across the full BMI range, with quadratic terms for each component testing nonlinearity. Second, brain structure was compared between CDC groups (healthy weight as reference) with likelihood ratio tests and contrasts, and group × age and group × sex interactions tested whether these group differences varied with age (estimated at ages 10, 13, and 16 years) or sex. Third, models were fitted within each group, with a quadratic within-person term testing nonlinearity (eTable 1); BMI-for-age z × group interactions for both components, fitted in all participants, tested whether slopes differed from those in healthy weight youth. Fourth, in all participants and within each group, we tested whether within-person associations themselves varied with age (within-person BMI-for-age z × age interaction, likelihood ratio test; simple slopes at ages 10, 13, and 16 years for significant regions) or sex (within-person × sex interactions, male participants as reference; sex-specific slopes). Sensitivity analyses added a random slope for age, excluded biologically implausible values (eMethods 2), and examined variance inflation factors and residual kurtosis (eMethods 4).

## Results

### Sample

The analytic sample comprised 11,510 participants (32,212 observations; ages 8.3 to 18.0 years; mean [SD] age, 12.6 [2.5] years; Figure 1; Table 1). Between-person variation was constrained by category width, most in the narrow overweight band (between-person SD, 0.18 vs 0.65 in healthy weight; eFigures 10 and 11).

**Table 1.**
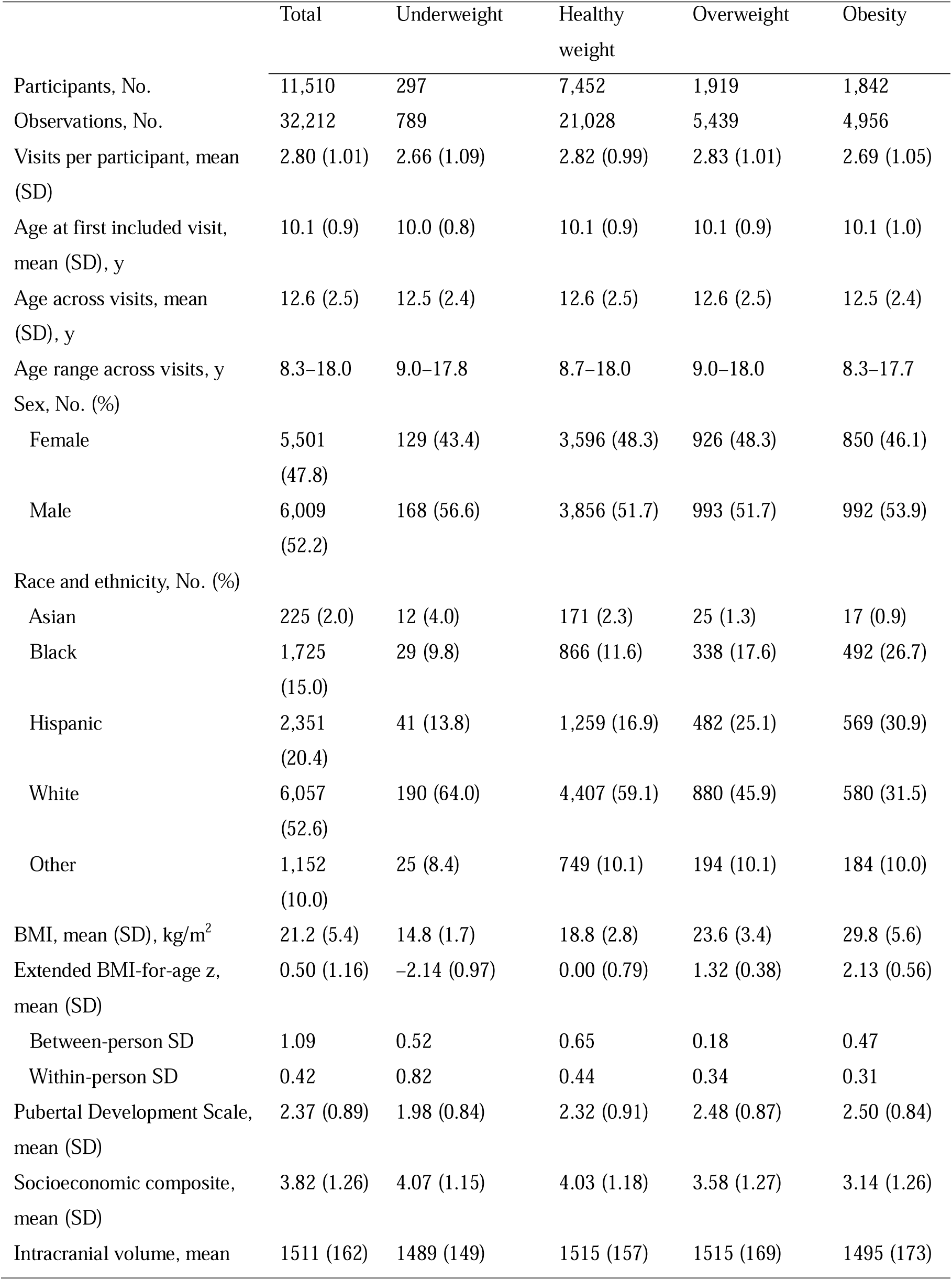
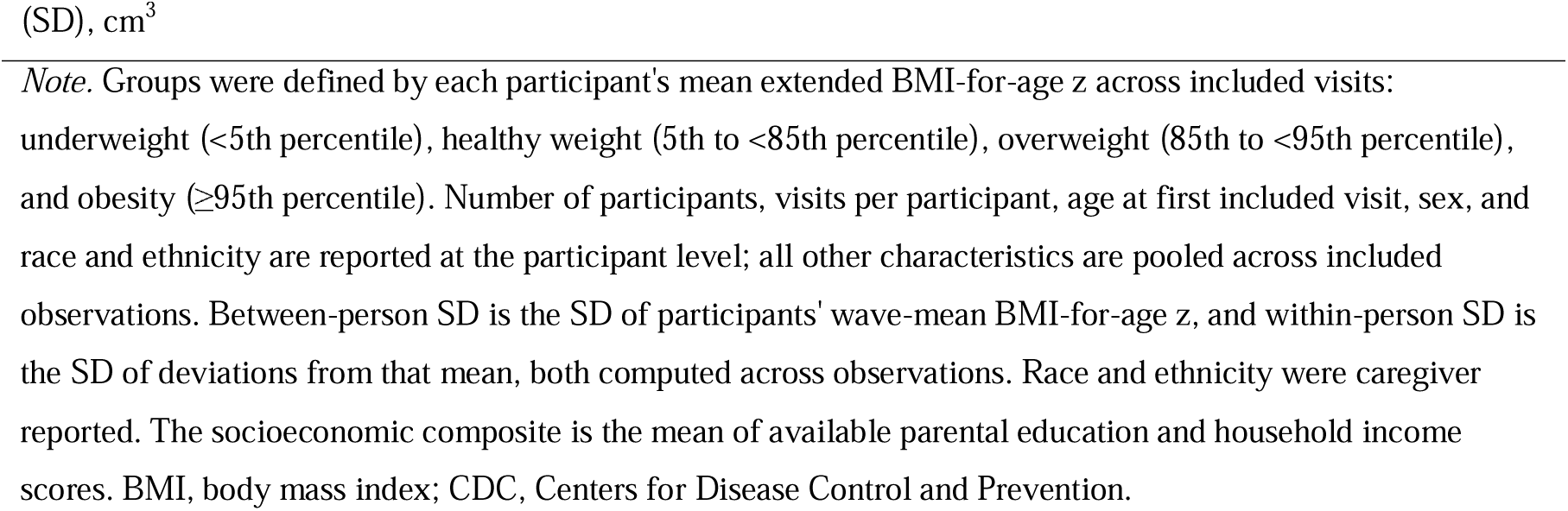
Characteristics of the Analytic Sample by CDC Weight Status Group.

|  | Total | Underweight | Healthy<br>weight | Overweight | Obesity |
| --- | --- | --- | --- | --- | --- |
| Participants, No. | 11,510 | 297 | 7,452 | 1,919 | 1,842 |
| Observations, No. | 32,212 | 789 | 21,028 | 5,439 | 4,956 |
| Visits per participant, mean<br>(SD) | 2.80 (1.01) | 2.66 (1.09) | 2.82 (0.99) | 2.83 (1.01) | 2.69 (1.05) |
| Age at first included visit,<br>mean (SD), y | 10.1 (0.9) | 10.0 (0.8) | 10.1 (0.9) | 10.1 (0.9) | 10.1 (1.0) |
| Age across visits, mean<br>(SD), y | 12.6 (2.5) | 12.5 (2.4) | 12.6 (2.5) | 12.6 (2.5) | 12.5 (2.4) |
| Age range across visits, y | 8.3–18.0 | 9.0–17.8 | 8.7–18.0 | 9.0–18.0 | 8.3–17.7 |
| Sex, No. (%) |  |  |  |  |  |
| Female | 5,501<br>(47.8) | 129 (43.4) | 3,596 (48.3) | 926 (48.3) | 850 (46.1) |
| Male | 6,009<br>(52.2) | 168 (56.6) | 3,856 (51.7) | 993 (51.7) | 992 (53.9) |
| Race and ethnicity, No. (%) |  |  |  |  |  |
| Asian | 225 (2.0) | 12 (4.0) | 171 (2.3) | 25 (1.3) | 17 (0.9) |
| Black | 1,725<br>(15.0) | 29 (9.8) | 866 (11.6) | 338 (17.6) | 492 (26.7) |
| Hispanic | 2,351<br>(20.4) | 41 (13.8) | 1,259 (16.9) | 482 (25.1) | 569 (30.9) |
| White | 6,057<br>(52.6) | 190 (64.0) | 4,407 (59.1) | 880 (45.9) | 580 (31.5) |
| Other | 1,152<br>(10.0) | 25 (8.4) | 749 (10.1) | 194 (10.1) | 184 (10.0) |
| BMI, mean (SD), kg/m<sup>2</sup> | 21.2 (5.4) | 14.8 (1.7) | 18.8 (2.8) | 23.6 (3.4) | 29.8 (5.6) |
| Extended BMI-for-age z,<br>mean (SD) | 0.50 (1.16) | –2.14 (0.97) | 0.00 (0.79) | 1.32 (0.38) | 2.13 (0.56) |
| Between-person SD | 1.09 | 0.52 | 0.65 | 0.18 | 0.47 |
| Within-person SD | 0.42 | 0.82 | 0.44 | 0.34 | 0.31 |
| Pubertal Development Scale,<br>mean (SD) | 2.37 (0.89) | 1.98 (0.84) | 2.32 (0.91) | 2.48 (0.87) | 2.50 (0.84) |
| Socioeconomic composite,<br>mean (SD) | 3.82 (1.26) | 4.07 (1.15) | 4.03 (1.18) | 3.58 (1.27) | 3.14 (1.26) |
| Intracranial volume, mean | 1511 (162) | 1489 (149) | 1515 (157) | 1515 (169) | 1495 (173) |

*Note.* Groups were defined by each participant's mean extended BMI-for-age z across included visits: underweight (<5th percentile), healthy weight (5th to <85th percentile), overweight (85th to <95th percentile), and obesity (≥95th percentile). Number of participants, visits per participant, age at first included visit, sex, and race and ethnicity are reported at the participant level; all other characteristics are pooled across included observations. Between-person SD is the SD of participants' wave-mean BMI-for-age z, and within-person SD is the SD of deviations from that mean, both computed across observations. Race and ethnicity were caregiver reported. The socioeconomic composite is the mean of available parental education and household income scores. BMI, body mass index; CDC, Centers for Disease Control and Prevention.

### Associations across the full BMI range

Higher between-person BMI-for-age z was associated with lower thickness in 6 frontal and temporal regions (β = −0.042 to −0.072) and larger volume in 5 subcortical regions (β = 0.026 to 0.049; all *q* < .001; eTable 2; eFigure 12). These associations were nonlinear, with negative between-person quadratic terms in 17 of 19 regions (*q* ≤ .035) indicating more negative cortical and weaker subcortical associations at higher BMI-for-age z (eTable 3). Within-person associations were small and linear: visits with higher-than-usual BMI-for-age z were accompanied by greater lateral occipital and middle temporal thickness, lower temporal pole thickness, and larger hippocampus and thalamus volume (β = −0.028 to 0.023; *q* ≤ .014). Between-person associations exceeded within-person associations in 5 cortical and 4 subcortical regions (contextual tests, *q* ≤ .009). Within-person associations varied with age in 7 cortical regions (*q* ≤ .038), shifting from negative at age 10 to positive at age 16 (eTables 4 and 5), and were more negative in female than in male participants in 14 regions (*q* ≤ .021; eTables 6 and 7).

### Differences between CDC weight status groups

Brain structure differed between groups in 18 of 19 regions (*q* ≤ .047; Figure 2; eTables 8 and 9). Compared with healthy weight youth, youth with obesity had lower thickness in 8 frontal and temporal regions (−0.077 to −0.206 SD; largest in rostral middle frontal cortex), larger pallidum volume, and smaller amygdala volume. Overweight youth had lower thickness in 4 of these regions and larger thalamus and pallidum volume, and underweight youth had smaller volume in 4 subcortical regions and lower lateral occipital thickness (all *q* < .05). Age-related change differed between groups in 7 regions (eTable 10). Only youth with obesity differed from healthy weight youth, with slower thinning in 4 frontal and cingulate regions (by 0.009 to 0.024 SD per year) and slower decline in accumbens volume (eTables 11 and 12). Consequently, superior and rostral middle frontal differences were less than half as large at age 16 as at age 10 (eg, rostral middle frontal, −0.261 vs −0.122 SD; both *q* < .001), whereas temporal differences did not decrease (eTable 13). Group differences did not vary by sex except in the accumbens (eTables 14–16).

### Cross-sectional associations within CDC weight status groups

Between-person associations differed qualitatively between groups (Figure 3). In the healthy weight group, higher BMI-for-age z was associated with larger volume in all 7 subcortical regions (β = 0.054 to 0.089; all *q* < .001), with small, mixed-direction cortical associations (β = −0.039 to 0.036; significant in 4 regions). In the obesity group, it was associated with lower thickness in 11 of 12 cortical regions (β = −0.110 to −0.270; all *q* < .05; strongest in fusiform cortex) and smaller amygdala volume (β = −0.086; *q* = .024). Obesity slopes were more negative than healthy weight slopes in 10 cortical and 4 subcortical regions (largest: fusiform, Δβ = −0.278; *q* < .001); the overweight and underweight groups showed no significant associations and did not differ from healthy weight youth (eTable 17, Section A; eTable 18; eFigures 13–15).

### Longitudinal associations within CDC weight status groups

Within-person associations also differed between groups (Figure 4). In the healthy weight group, visits with higher-than-usual BMI-for-age z were accompanied by greater lateral occipital, middle temporal, and fusiform thickness (β = 0.020 to 0.036) and larger thalamus, hippocampus, pallidum, and putamen volume (β = 0.013 to 0.022; all *q* < .05). No within-person association was significant in the other groups, although obesity slopes were lower than healthy weight slopes in 5 cortical and 2 subcortical regions (eTable 18). Associations were linear in all groups (eTable 1). Between-person associations exceeded within-person associations in 9 cortical regions in the obesity group and all 7 subcortical regions in the healthy weight group (contextual tests, *q* < .05; eTable 17, Sections B and C).

### Age moderation of within-person associations

Within-person associations varied with age in 4 cortical regions in the healthy weight group, 3 in the overweight group, and 1 in the obesity group (likelihood ratio χ² = 7.14 to 18.54; *q* ≤ .048; eFigure 16; eTables 19 and 20). At age 10, frontal associations were negative in each of these groups (superior frontal: healthy weight, β = −0.051; obesity, β = −0.146; rostral middle frontal: overweight, β = −0.118; *q* ≤ .004); at age 16, associations were significantly positive in 7 of the 8 regions (β = 0.040 to 0.132). In the overweight group, these opposite-signed associations averaged to near zero. No age moderation occurred in the underweight group or in any subcortical region.

### Sex moderation of within-person associations

In the healthy weight group, within-person × sex interactions were significant in 13 of 19 regions, all indicating more negative slopes in female participants: male participants showed positive temporal and subcortical associations (eg, middle temporal, β = 0.060), whereas female participants showed negative frontal associations (eg, superior frontal, β = −0.054; both *q* < .001) (eFigure 17; eTables 21 and 22). Interactions were significant in 3 subcortical regions in the overweight group, with positive slopes in male participants, and in no region in the obesity or underweight groups.

### Sensitivity analyses

Within-person findings were robust to a random slope for age (coefficient changes ≤0.009) and to excluding 6 biologically implausible observations (≤0.021), with no significant finding lost and 2 obesity-group associations (insula, temporal pole) becoming significant (*q* ≤ .025). Variance inflation factors were below 5, and residual kurtosis was highest in caudate models (eTables 23–25).

## Discussion

Brain structure and its development differed across CDC weight status categories. Compared with healthy weight youth, youth with obesity had lower frontal and temporal cortical thickness, consistent with prior work^16–18,22^, but slower frontal and cingulate thinning, so that dorsal frontal differences narrowed across adolescence. Overweight youth showed attenuated thickness differences, and underweight youth had smaller subcortical volumes. BMI-for-age associations within categories also differed qualitatively: within the obesity group, higher between-person BMI-for-age z was associated with lower thickness in 11 of 12 regions, a pattern not reproduced by within-person change, whereas within the healthy weight group it was associated with larger volume in all 7 subcortical regions.

Group differences and within-group associations were concordant in direction but differed in regional emphasis: group differences were most marked in frontal cortex, whereas within the obesity group BMI-for-age z was most strongly associated with temporo-occipital thickness. Slower frontal thinning in obesity accords with decelerated prefrontal development at higher BMI in ABCD^18^ and, with frontal cortex already thinner at age 10, could reflect earlier maturation not captured by pubertal stage, or partial convergence. Subcortically, smaller volumes in underweight youth paralleled the positive association within the healthy weight range.

Within the healthy weight range, BMI differences may partly reflect normal variation in lean mass and somatic growth rather than excess adiposity. Positive subcortical associations in mixed-weight pediatric samples^19–21^ could therefore reflect such variation, because here these associations were absent above the healthy weight range and independent of intracranial volume. Together with the nonlinearity across the full range, this suggests that associations in healthy weight and obesity may not lie on a single continuum.

In obesity, lower thickness at higher BMI-for-age z is consistent with evidence that obesity-associated inflammation partially mediates BMI-related cortical thinning in adolescents^44^ and that insulin resistance is associated with thinner medial orbitofrontal cortex in female adolescents^45^. A single linear slope across the full range underestimated these effects (fusiform β = −0.012 vs −0.270 in obesity; eFigure 18).

Between- and within-person associations diverged in both groups, subcortically in healthy weight youth and cortically in youth with obesity, where lower thickness in 9 regions was not reproduced by within-person change. Consistent with bidirectional prospective evidence^28^, prenatal programming models^24,25^, and our prior cardiometabolic work in ABCD^27^, these differences appear to reflect largely stable traits, possibly early-emerging phenotypes that predispose to weight gain, rather than concurrent coupling. Modest within-person coupling cannot be excluded, however: within-person slopes in obesity were lower than in healthy weight youth in 5 cortical regions, and the insula and temporal pole associations reached significance in sensitivity analyses.

The null between-person findings in the overweight group likely reflect the narrow category, with approximately 14-fold smaller between-person variance than in healthy weight youth (eFigure 11), rather than an absence of brain differences: overweight youth differed from healthy weight youth in fronto-temporal thickness and subcortical volume, and their within-person associations varied with age. Within-person fluctuation was large relative to the 0.61-unit band, so overweight classification is likely to change between visits, a consideration for single-visit screening.

The within-person reversal from negative frontal associations at age 10 to positive associations by age 16 may reconcile shorter-window studies that captured only one side of this trajectory^46,47^; visceral fat accumulation in later adolescence, which, unlike BMI, was associated with greater cortical thickness^48^, offers one mechanism for the late-adolescent slope.

The male-positive, female-negative within-person divergence, present across the full BMI range and in 13 regions in healthy weight youth with pubertal stage as a covariate, may be an early manifestation of the sex-differentiated body–brain coupling documented in adults^29,30^. Its narrowing in the overweight and obesity groups may partly reflect their smaller size; disrupted sex-specific hormonal profiles in pubertal obesity^49^ offer a biological explanation warranting future research.

### Strengths and Limitations

BMI-for-age z does not distinguish fat from lean mass or visceral from subcutaneous fat, which relate to brain structure independently of BMI, at least in adults^10,11^. Changes in z-scores do not map uniformly onto adiposity change^31^, and within-person variation was smallest in the obesity group (SD, 0.31 vs 0.44 in healthy weight), so within-person null findings there warrant caution. Although the ABCD Study was designed to reflect the US population’s ethnic and sociodemographic composition^15^, findings may not generalize to other countries. Group differences and between-person estimates remain open to stable confounding, and within-person estimates remove stable but not time-varying confounding; reverse causation, whereby brain phenotypes influencing eating behavior or metabolism increase obesity risk^28^, cannot be excluded.

Defining groups by wave-mean BMI-for-age z restricts between-person variance, most in the overweight band, and participants near boundaries may have visits in adjacent categories. Group sizes differed (297 to 7,452 participants), so differences in the number of significant findings partly reflect power; between-group differences in associations are best judged from the interaction tests, and underweight findings should be interpreted with caution. Finally, the primary within-person estimate averages over ages at which slopes have opposite signs, and sex moderation was estimated at average age.

Strengths include the extended CDC BMI-for-age z-score^33^, the within-between effects decomposition, direct comparisons of brain structure and BMI slopes across groups, and, to our knowledge, the broadest developmental window of any longitudinal ABCD BMI–brain study.

## Conclusion

Brain structure and its associations with BMI-for-age differed across CDC weight status categories, and obesity-related cortical differences were already evident by late childhood and did not track concurrent weight change. Because pediatric obesity tends to persist and carries cardiometabolic and mental health risks, longitudinal studies beginning earlier in childhood are needed to establish when these brain differences emerge and whether they relate to later health outcomes. Such studies should not assume a single linear BMI–brain association across the weight spectrum.

## Data Availability

All data produced were obtained from the Adolescent Brain Cognitive Development (ABCD) Study, held in the NIH Brain Development Cohorts Data Sharing Platform.

## Acknowledgments

Data used in the preparation of this article were obtained from the <u>Adolescent Brain</u> <u>Cognitive Development™ (ABCD) Study</u>, held in the <u>NIH Brain Development Cohorts Data</u> <u>Sharing Platform</u>. This is a multisite, longitudinal study designed to recruit more than 10,000 children aged 9-10 and follow them over 10 years into early adulthood. The ABCD Study® is supported by the National Institutes of Health and additional federal partners under award numbers: U01DA041048, U01DA050989, U01DA051016, U01DA041022, U01DA051018, U01DA051037, U01DA050987, U01DA041174, U01DA041106, U01DA041117, U01DA041028, U01DA041134, U01DA050988, U01DA051039, U01DA041156, U01DA041025, U01DA041120, U01DA051038, U01DA041148, U01DA041093, U01DA041089, U24DA041123, U24DA041147. A full list of supporters is available at <u>Federal Partners - ABCD Study</u>. ABCD Consortium investigators designed and implemented the study and/or provided data but did not necessarily participate in the analysis or writing of this report. This manuscript reflects the views of the authors and may not reflect the opinions or views of the NIH or ABCD Consortium investigators. Data were handled inside Service for Sensitive Data (TSD), a platform owned by the University of Oslo, operated and developed by the TSD service group at the University of Oslo IT-Department (USIT). Computations were also performed using resources provided by UNINETT Sigma2 – the National Infrastructure for High-Performance Computing and Data Storage in Norway (NS9666S).

## Data Sharing Statement

The data that support the findings of this study are available from the Adolescent Brain Cognitive Development™ (ABCD) Study via the NIH Brain Development Cohorts (NBDC) Data Hub. The specific dataset used was ABCD Study Release 7.0 (DOI: https://doi.org/10.82525/jy7n-g441). Access to these data is subject to approval through the NIH Data Use Certification (DUC) process, and the data are available to qualified researchers upon reasonable request and with appropriate authorization via the NBDC Data Hub (https://nbdc-datahub.org). Statistical analysis code is available from the corresponding author upon reasonable request.

## CRediT author contribution statement

Author Contributions: Dr Beck had full access to all of the data in the study and takes responsibility for the integrity of the data and the accuracy of the data analysis. *Concept and design:* All authors. *Acquisition, analysis, or interpretation of data:* Beck. *Drafting of the manuscript:* Beck. *Critical review of the manuscript for important intellectual content:* All authors. *Statistical analysis:* Beck. *Obtained funding:* Westlye, Tamnes. *Administrative, technical, or material support:* Tamnes.

## References

1. Kelly AS, Armstrong SC, Michalsky MP, Fox CK. Obesity in Adolescents: A Review. JAMA. Published online August 5, 2024. doi:10.1001/jama.2024.11809

2. Skinner AC, Ravanbakht SN, Skelton JA, Perrin EM, Armstrong SC. Prevalence of Obesity and Severe Obesity in US Children, 1999–2016. Pediatrics. 2018;141(3):e20173459. doi:10.1542/peds.2017-3459

3. Geserick M, Vogel M, Gausche R, et al. Acceleration of BMI in Early Childhood and Risk of Sustained Obesity. N Engl J Med. 2018;379(14):1303–1312. doi:10.1056/NEJMoa1803527

4. Simmonds M, Llewellyn A, Owen CG, Woolacott N. Predicting adult obesity from childhood obesity: a systematic review and meta-analysis. Obes Rev Off J Int Assoc Study Obes. 2016;17(2):95–107. doi:10.1111/obr.12334

5. Halfon N, Larson K, Slusser W. Associations Between Obesity and Comorbid Mental Health, Developmental, and Physical Health Conditions in a Nationally Representative Sample of US Children Aged 10 to 17. Acad Pediatr. 2013;13(1):6–13. doi:10.1016/j.acap.2012.10.007

6. Karhunen V, Bond TA, Zuber V, et al. The link between attention deficit hyperactivity disorder (ADHD) symptoms and obesity-related traits: genetic and prenatal explanations. Transl Psychiatry. 2021;11(1):455. doi:10.1038/s41398-021-01584-4

7. Hughes AM, Sanderson E, Morris T, et al. Body mass index and childhood symptoms of depression, anxiety, and attention-deficit hyperactivity disorder: A within-family Mendelian randomization study. eLife. 2022;11:e74320. doi:10.7554/eLife.74320

8. Cox SR, Lyall DM, Ritchie SJ, et al. Associations between vascular risk factors and brain MRI indices in UK Biobank. Eur Heart J. 2019;40(28):2290–2300. doi:10.1093/eurheartj/ehz100

9. Beck D, de Lange AMG, Pedersen ML, et al. Cardiometabolic risk factors associated with brain age and accelerate brain ageing. Hum Brain Mapp. 2022;43(2):700–720. doi:10.1002/hbm.25680

10. Beck D, de Lange AMG, Alnæs D, et al. Adipose tissue distribution from body MRI is associated with cross-sectional and longitudinal brain age in adults. NeuroImage Clin. 2022;33:102949. doi:10.1016/j.nicl.2022.102949

11. Beck D, de Lange AMG, Gurholt TP, et al. Dissecting unique and common variance across body and brain health indicators using age prediction. Hum Brain Mapp. 2024;45(6):e26685. doi:10.1002/hbm.26685

12. Willette AA, Kapogiannis D. Does the brain shrink as the waist expands? Ageing Res Rev. 2015;20:86–97. doi:10.1016/j.arr.2014.03.007

13. Gómez-Apo E, Mondragón-Maya A, Ferrari-Díaz M, Silva-Pereyra J. Structural Brain Changes Associated with Overweight and Obesity. J Obes. 2021;2021(1):6613385. doi:10.1155/2021/6613385

14. Casey BJ, Cannonier T, Conley MI, et al. The Adolescent Brain Cognitive Development (ABCD) study: Imaging acquisition across 21 sites. Dev Cogn Neurosci. 2018;32:43–54. doi:10.1016/j.dcn.2018.03.001

15. Karcher NR, Barch DM. The ABCD study: understanding the development of risk for mental and physical health outcomes. Neuropsychopharmacology. 2021;46(1):131–142. doi:10.1038/s41386-020-0736-6

16. Laurent JS, Watts R, Adise S, et al. Associations Among Body Mass Index, Cortical Thickness, and Executive Function in Children. JAMA Pediatr. 2020;174(2):170–177. doi:10.1001/jamapediatrics.2019.4708

17. Ronan L, Alexander-Bloch A, Fletcher PC. Childhood Obesity, Cortical Structure, and Executive Function in Healthy Children. Cereb Cortex. 2020;30(4):2519–2528. doi:10.1093/cercor/bhz257

18. Kaltenhauser S, Weber CF, Lin H, et al. Association of Body Mass Index and Waist Circumference With Imaging Metrics of Brain Integrity and Functional Connectivity in Children Aged 9 to 10 Years in the US, 2016-2018. JAMA Netw Open. 2023;6(5):e2314193. doi:10.1001/jamanetworkopen.2023.14193

19. Rapuano KM, Zieselman AL, Kelley WM, Sargent JD, Heatherton TF, Gilbert-Diamond D. Genetic risk for obesity predicts nucleus accumbens size and responsivity to real-world food cues. Proc Natl Acad Sci. 2017;114(1):160–165. doi:10.1073/pnas.1605548113

20. de Groot CJ, van den Akker ELT, Rings EHHM, Delemarre-van de Waal HA, van der Grond J. Brain structure, executive function and appetitive traits in adolescent obesity. Pediatr Obes. 2017;12(4):e33–e36. doi:10.1111/ijpo.12149

21. Perlaki G, Molnar D, Smeets PAM, et al. Volumetric gray matter measures of amygdala and accumbens in childhood overweight/obesity. PLOS ONE. 2018;13(10):e0205331. doi:10.1371/journal.pone.0205331

22. Morys F, Tremblay C, Rahayel S, et al. Neural correlates of obesity across the lifespan. Commun Biol. 2024;7(1):656. doi:10.1038/s42003-024-06361-9

23. Adise S, Allgaier N, Laurent J, et al. Multimodal brain predictors of current weight and weight gain in children enrolled in the ABCD study ®. Dev Cogn Neurosci. 2021;49:100948. doi:10.1016/j.dcn.2021.100948

24. Norr ME, Hect JL, Lenniger CJ, Van den Heuvel M, Thomason ME. An examination of maternal prenatal BMI and human fetal brain development. J Child Psychol Psychiatry. 2021;62(4):458–469. doi:10.1111/jcpp.13301

25. Skowronski AA, Leibel RL, LeDuc CA. Neurodevelopmental Programming of Adiposity: Contributions to Obesity Risk. Endocr Rev. 2024;45(2):253–280. doi:10.1210/endrev/bnad031

26. Kentistou KA, Sundfjord J, Karimi R, et al. Genome-wide mapping of common and rare variant effects on adiposity across childhood. Nat Genet. Published online September 25, 2026:1–10. doi:10.1038/s41588-026-02772-y

27. Beck D, Westlye LT, Tamnes CK. Cardiometabolic Risk and Structural Brain Development in a Large Community-Based US Cohort. Hum Brain Mapp. 2026;47(8):e70575. doi:10.1002/hbm.70575

28. Sakib MN, Best JR, Hall PA. Bidirectional Associations Between Adiposity and Cognitive Function and Mediation by Brain Morphology in the ABCD Study. JAMA Netw Open. 2023;6(2):e2255631. doi:10.1001/jamanetworkopen.2022.55631

29. Gurholt TP, Beck D, Voldsbekk I, et al. Linking obesity with white matter microstructure highlights the importance of brainstem tracts and sex differences. Brain Commun. 2026;8(1):fcag026. doi:10.1093/braincomms/fcag026

30. Mueller K, Anwander A, Möller HE, et al. Sex-Dependent Influences of Obesity on Cerebral White Matter Investigated by Diffusion-Tensor Imaging. PLOS ONE. 2011;6(4):e18544. doi:10.1371/journal.pone.0018544

31. Adise S, Rhee KE, Laurent J, et al. Limitations of BMI z scores for assessing weight change: A clinical tool versus individual risk. Obesity. 2024;32(3):445–449. doi:10.1002/oby.23957

32. Freedman DS, Butte NF, Taveras EM, et al. BMI z-Scores are a poor indicator of adiposity among 2- to 19-year-olds with very high BMIs, NHANES 1999-2000 to 2013-2014. Obesity. 2017;25(4):739–746. doi:10.1002/oby.21782

33. CDC. Growth Charts - CDC Extended BMI-for-Age Growth Charts. October 15, 2024. Accessed June 22, 2026. https://www.cdc.gov/growthcharts/extended-bmi.htm

34. Auchter AM, Hernandez Mejia M, Heyser CJ, et al. A description of the ABCD organizational structure and communication framework. Dev Cogn Neurosci. 2018;32:8–15. doi:10.1016/j.dcn.2018.04.003

35. Hagler DJ, Hatton SeanN, Cornejo MD, et al. Image processing and analysis methods for the Adolescent Brain Cognitive Development Study. NeuroImage. 2019;202:116091. doi:10.1016/j.neuroimage.2019.116091

36. Fischl B, Salat DH, Busa E, et al. Whole Brain Segmentation. Neuron. 2002;33(3):341–355. doi:10.1016/S0896-6273(02)00569-X

37. Dale AM, Fischl B, Sereno MI. Cortical Surface-Based Analysis: I. Segmentation and Surface Reconstruction. NeuroImage. 1999;9(2):179–194. doi:10.1006/nimg.1998.0395

38. Beer JC, Tustison NJ, Cook PA, et al. Longitudinal ComBat: A method for harmonizing longitudinal multi-scanner imaging data. NeuroImage. 2020;220:117129. doi:10.1016/j.neuroimage.2020.117129

39. Freedman D. cdcanthro: Sex- and Age-Standardized Metrics from the Centers for Disease Control and Prevention (CDC) Growth Charts. Published online May 22, 2026. Accessed June 22, 2026. https://cran.r-project.org/web/packages/cdcanthro/index.html

40. R Core Team. R: A Language and Environment for Statistical Computing. 2025. Accessed June 22, 2026. https://www.r-project.org/

41. Pinheiro J, Bates D, R-core. Nlme: Linear and Nonlinear Mixed Effects Models. 2022. https://svn.r-project.org/R-packages/trunk/nlme/

42. Benjamini Y, Hochberg Y. Controlling the False Discovery Rate: A Practical and Powerful Approach to Multiple Testing. J R Stat Soc Ser B Methodol. 1995;57(1):289–300. doi:10.1111/j.2517-6161.1995.tb02031.x

43. Mowinckel AM, Vidal-Piñeiro D. Visualization of Brain Statistics With R Packages ggseg and ggseg3d. Adv Methods Pract Psychol Sci. 2020;3(4):466–483. doi:10.1177/2515245920928009

44. Prats-Soteras X, Jurado MA, Ottino-González J, et al. Inflammatory agents partially explain associations between cortical thickness, surface area, and body mass in adolescents and young adulthood. Int J Obes. 2020;44(7):1487–1496. doi:10.1038/s41366-020-0582-y

45. Gabay A, London S, Yates KF, Convit A. Does obesity-associated insulin resistance affect brain structure and function of adolescents differentially by sex? Psychiatry Res Neuroimaging. 2022;319:111417. doi:10.1016/j.pscychresns.2021.111417

46. Jiang F, Li G, Ji W, et al. Obesity is associated with decreased gray matter volume in children: a longitudinal study. Cereb Cortex N Y NY. 2022;33(7):3674–3682. doi:10.1093/cercor/bhac300

47. Adise S, Marshall AT, Hahn S, et al. Longitudinal assessment of brain structure and behaviour in youth with rapid weight gain: Potential contributing causes and consequences. Pediatr Obes. 2023;18(2):e12985. doi:10.1111/ijpo.12985

48. Saute RL, Soder RB, Alves Filho JO, Baldisserotto M, Franco AR. Increased brain cortical thickness associated with visceral fat in adolescents. Pediatr Obes. 2018;13(1):74–77. doi:10.1111/ijpo.12190

49. Nokoff N, Thurston J, Hilkin A, et al. Sex Differences in Effects of Obesity on Reproductive Hormones and Glucose Metabolism in Early Puberty. J Clin Endocrinol Metab. 2019;104(10):4390–4397. doi:10.1210/jc.2018-02747

