## Supplementary Information for "Beyond the Percentile: CDC-referenced BMI-for-Age and Brain Structure Across Childhood and Adolescence"

**Contents**

eMethods 1. Data Sources and Linkage
eMethods 2. Data Cleaning and Quality Assurance
eMethods 3. LongCombat Harmonization

eMethods 4. Statistical Analysis Details
eFigure 1. Missingness Across Variables and Waves
eFigure 2. Longitudinal ComBat Harmonization of Cortical Thickness: Before and After, by Scanner
eFigure 3. Longitudinal ComBat Harmonization of Subcortical Volume: Before and After, by Scanner
eFigure 4. Conventional vs Extended CDC BMI-for-Age z in the Analytic Sample
eFigure 5. Correlation Matrix of Raw BMI, Extended BMI-for-Age z, and Model Covariates
eFigure 6. BMI Distribution Across Waves in the Analytic Sample
eFigure 7. Race and Ethnicity of the Analytic Sample
eFigure 8. Socioeconomic Status Distribution of the Analytic Sample
eFigure 9. Pubertal Development by Sex in the Analytic Sample
eTable 1. Test of Quadratic Within-Person BMI-for-Age z Term as a Pre-Specified Linearity Check of Longitudinal BMI–Brain Associations Within Each CDC Weight Status Group
eFigure 10. Individual BMI-for-Age z Trajectories Across Adolescence by CDC Weight Status Group
eFigure 11. BMI-for-Age z Variance Decomposition by CDC Weight Status Group
eTable 2. Full Range: Between-Person, Within-Person, and Contextual Associations of Extended BMI-for-Age z With Regional Brain Structure
eFigure 12. Between- and Within-Person Associations of Extended BMI-for-Age z With Regional Brain Structure Across the Full BMI Range
eTable 3. Full Range: Test of Quadratic Between-Person and Within-Person BMI-for-Age z Terms
eTable 4. Likelihood Ratio Tests for Age Moderation of Within-Person BMI-for-Age z Associations With Regional Brain Structure Across the Full BMI Range
eTable 5. Simple Slopes of Within-Person BMI-for-Age z Associations With Cortical Thickness at Ages 10, 13, and 16 Years Across the Full BMI Range
eTable 6. Within-Person × Sex Interaction Terms for BMI-for-Age z Associations With Brain Structure Across the Full BMI Range
eTable 7. Sex-Specific Within-Person BMI-for-Age z Associations With Brain Structure Across the Full BMI Range
eTable 8. Likelihood Ratio Tests for Differences in Regional Brain Structure Between CDC Weight Status Groups
eTable 9. Differences in Regional Brain Structure Between Each CDC Weight Status Group and the Healthy Weight Group
eTable 10. Likelihood Ratio Tests for Differences in Age-Related Change in Regional Brain Structure Between CDC Weight Status Groups
eTable 11. Age Slopes of Regional Brain Structure Within Each CDC Weight Status Group
eTable 12. Differences in Age Slopes of Regional Brain Structure Between Each CDC Weight Status Group and the Healthy Weight Group
eTable 13. Differences in Regional Brain Structure Between Each CDC Weight Status Group and the Healthy Weight Group at Ages 10, 13, and 16 Years
eTable 14. Likelihood Ratio Tests for Weight Status Group × Sex Interactions on Regional Brain Structure
eTable 15. Weight Status Group × Sex Interaction Terms for Regional Brain Structure
eTable 16. Sex-Specific Differences in Regional Brain Structure Between Each CDC Weight Status Group and the Healthy Weight Group
eTable 17. Stratified Between-Person, Within-Person, and Contextual Associations of Extended BMI-for-Age z With Regional Brain Structure by CDC Weight Status Group
eTable 18. Formal Group Comparison: Interaction Tests for Between-Person and Within-Person BMI-for-Age z Associations Versus the Healthy Weight Reference Group
eFigure 13. LOESS Associations Between Extended BMI-for-Age z and Cortical Thickness by CDC Weight Status Group
eFigure 14. LOESS Associations Between Extended BMI-for-Age z and Subcortical Volume by CDC Weight Status Group
eFigure 15. Between-Person BMI-for-Age z Associations With Brain Structure Across All CDC Weight Status Groups
eFigure 16. Age Moderation of Within-Person BMI-for-Age z Associations With Cortical Thickness Across Adolescence by CDC Weight Status Group
eTable 19. Likelihood Ratio Tests for Age Moderation of Within-Person BMI-for-Age z Associations With Regional Brain Structure by CDC Weight Status Group
eTable 20. Simple Slopes of Within-Person BMI-for-Age z Associations With Cortical Thickness at Ages 10, 13, and 16 Years for Regions Showing Significant Age Moderation, by CDC Weight Status Group
eFigure 17. Sex Moderation of Within-Person BMI-for-Age z Associations With Brain Structure by CDC Weight Status Group
eTable 21. Within-Person × Sex Interaction Terms for BMI-for-Age z Associations With Brain Structure by CDC Weight Status Group
eTable 22. Sex-Specific Within-Person BMI-for-Age z Associations With Brain Structure by CDC Weight Status Group
eTable 23. Sensitivity Analysis: Within-Person Estimates With Random Slope for Age, by CDC Weight Status Group
eTable 24. Sensitivity Analysis: Within-Person Estimates After Excluding Biologically Implausible BMI Values, by CDC Weight Status Group
eTable 25. Model Diagnostics: Maximum Variance Inflation Factor and Residual Kurtosis by CDC Weight Status Group
eFigure 18. Comparison of Full Range (pooled) vs Obesity-Specific Between-Person BMI-for-Age z Associations With Brain Structure
eReferences

**eMethods 1. Data Sources and Linkage.**

All measures were drawn from the ABCD Study Release 7.0 tabulated phenotype files and linked by participant and session identifiers. Chronological age, parental education, and household income were taken from the general dynamic file (ab_g_dyn); sex and race and ethnicity from the static background file (ab_g_stc); measured height and weight from the youth anthropometric file (ph_y_anthr); pubertal status from the parent-report Pubertal Development Scale file (ph_p_pds); family relatedness from the genetic-relatedness file (gn_y_genrel); cortical thickness from the Desikan–Killiany parcellation file (mr_y_smri__thk__dsk); subcortical volume from the aseg parcellation file (mr_y_smri__vol__aseg); T1 quality-control inclusion flags from mr_y_qc__incl; and scanner site identity from the MRI administrative file (mr_y_adm__info). All files were merged on the combination of participant_id and session_id. Only observations with T1 inclusion flag equal to 1 were retained.

**eMethods 2**. Data Cleaning and Quality Assurance.

Data were drawn from Release 7.0, which contained 42,180 observations from 11,860 participants across four waves. Of these, 33,456 observations from 11,794 participants had valid T1-weighted images passing the ABCD quality-control inclusion criterion. Harmonization required non-missing BMI and socioeconomic status, leaving 33,257 observations from 11,785 participants. The analytic sample was then restricted to observations with complete data on every model variable: BMI-for-age z after the anthropometric plausibility screen (33,209 observations, 11,778 participants), pubertal status (32,650; 11,693), and family identifier, required for the family-level random intercept (32,212; 11,510). No further observations were lost for socioeconomic status, race and ethnicity, sex, intracranial volume, or brain outcomes. The analytic sample therefore comprised 32,212 observations from 11,510 participants, and the within-between decomposition and CDC weight status groups were derived on this sample. The pattern of missingness in the imaging sample is shown in eFigure 1.

**Body Mass Index (BMI)**. Anthropometric plausibility was assessed in two stages. First, observations with physically implausible raw values (height < 110 cm or > 200 cm, weight < 18 kg or > 150 kg, or BMI < 10 or > 60 kg/m²) were set to missing at that wave, retaining the participant's remaining observations for analysis. These cutoffs were chosen to identify likely data-entry or unit-conversion errors – for example, height or weight inadvertently recorded in imperial rather than metric units – rather than genuine extreme physiological values. Where height or weight was missing at an intermediate wave but available at adjacent waves, the missing value was recovered by within-person linear interpolation between the two flanking measurements, without extrapolation beyond a participant's observed range; this preserved BMI data for waves where one anthropometric component was unavailable. BMI distribution across waves is shown in eFigure 6.

Second, biologically implausible BMI values (BIVs) were identified using the modified z-score (mod_bmiz) provided by the cdcanthro R package^1^, with thresholds of mod_bmiz < −4 or > +8 following Centers for Disease Control and Prevention (CDC) guidance^2^. BIV-flagged observations were retained in the primary analysis for two reasons: (1) the extended CDC BMI-for-age z-score used as the exposure was designed to accommodate the upper tail of the BMI distribution without compression, so high values carry meaningful signal rather than distorting the z-score scale; and (2) a CDC validation study of 8.8 million children found that many flagged values were unlikely to represent data errors and that excluding them underestimated obesity prevalence^3^. In the analytic sample, 6 observations were flagged (2 high values in the obesity group and 4 low values in the underweight group). These were removed only in a sensitivity analysis, where their exclusion did not change the direction or significance of any significant estimate, although 2 obesity-group associations (insula and temporal pole) became significant (eTable 24).

**Structural MRI**. The ABCD Study consortium's centralized quality control pipeline screens for motion artifacts, signal dropout, and reconstruction failures using both automated metrics and visual inspection. No additional manual quality control was applied beyond the standard T1 inclusion flag (mr_y_qc__incl; t1_indicator = 1). The T1 inclusion criterion was applied prior to data merging, so participants contributing imaging data at one wave but failing quality control at another contributed only their passing waves to the analysis.

Following quality control, scanner-related variance was removed using longCombat (see eMethods 3). The harmonization model preserved age, sex, study wave, raw BMI, socioeconomic status, and race and ethnicity as covariates of interest, and included a participant-level random intercept to account for the repeated-measures structure of the data and prevent removal of within-person longitudinal signal. LongCombat was applied across the 34 scanners contributing data to ABCD Release 7.0, as identified by the scanner site identity variable in the MRI administrative file (mr_y_adm__info). Following harmonization, left- and right-hemisphere regional values were averaged to bilateral measures for analysis. Visual inspection of before-and-after longCombat distributions confirmed that scanner-related clustering was substantially reduced while the age-related developmental trajectories of brain structure were preserved.

Nineteen bilateral brain structure metrics were chosen for the study, including thickness in 12 cortical regions (medial and lateral orbitofrontal, rostral and caudal anterior cingulate, superior frontal, rostral middle frontal, temporal pole, insula, fusiform, inferior temporal, middle temporal, and lateral occipital cortex) and volume of 7 subcortical regions (nucleus accumbens, amygdala, caudate, hippocampus, pallidum, putamen, and thalamus). These measures were selected a priori based on prior literature consistently linking higher adiposity to lower cortical thickness in fronto-temporal and orbitofrontal cortex and larger volume in subcortical reward- and homeostasis-related structures in pediatric and adult samples^4–12^; cortical thickness was prioritized over surface area as the measure most sensitive to neurodevelopmental change across adolescence and most consistently associated with BMI in prior ABCD-based work^6,8,13^.

**Covariates.** Pubertal Development Scale. Pubertal status was derived from the parent-report Pubertal Development Scale (PDS) using the ABCD Release 7.0 validated summary scores (ph_p_pds__m_mean for male and ph_p_pds__f_mean for female participants). These pre-computed means are validated by the ABCD consortium as requiring no more than one missing item across the five PDS questions and handle the sex-specific menarche item mapping internally, avoiding the errors and biases introduced by hand-scoring the raw (now character-coded) items. The two sex-specific means are mutually exclusive by participant sex, so a single PDS variable was created by coalescing the two columns. Coverage was verified at each wave; parent-reported PDS missingness was low (1%–2% per wave in the imaging sample, consistent with published ABCD estimates^14^), and observations missing PDS were excluded from models incorporating it as a covariate under the complete-case approach. The distribution of pubertal development by sex is shown in eFigure 9.

Socioeconomic Status. Socioeconomic status was operationalized as a composite of parental education and household income (their mean when both were available, or the single available value otherwise), both drawn from the general dynamic file (ab_g_dyn). When a participant had no composite value at any follow-up imaging wave (2, 4, or 6 years) but had one at an intermediate annual visit (1 or 3 years), that value was used for the missing follow-up waves, since socioeconomic status was treated as a slowly changing background characteristic rather than a time-varying exposure. This composite ranged from 1 to 5.5 in the analytic sample. The SES distribution of the analytic sample is shown in eFigure 8.

Race and Ethnicity. Caregiver-reported race and ethnicity (White, Hispanic, Black, Asian, Other) was drawn from the static background file (ab_g_stc) and treated as time-invariant. This variable is recorded at enrollment and not repeated; it was missing for 1 observation in the imaging sample, which was also missing other model variables, so missingness was zero in the analytic sample. The race and ethnicity composition of the analytic sample is shown in eFigure 7.

Age. Chronological age at each visit was taken from the general dynamic file (ab_g_dyn__visit_age) and was complete for all retained observations (0% missing across waves), as age is derived from the visit date and date of birth rather than participant report.

Intracranial Volume. Total intracranial volume (ICV), used as a covariate in subcortical volume models, was taken from the aseg parcellation file after longCombat harmonization. ICV is a structural constant that scales with head size and was not subject to separate quality control beyond the T1 inclusion flag applied to all MRI measures.

**eMethods 3.** LongCombat Harmonization

The ABCD Study collects MRI data across 21 sites using 34 distinct scanners from three manufacturers (Siemens Prisma, General Electric 750, and Philips, all 3T). Technical variability across scanners introduces additive and multiplicative batch effects unrelated to biology. Longitudinal ComBat (longCombat^15^) was applied separately to cortical thickness and subcortical volume to remove scanner-related variance while preserving within-person longitudinal signal – an approach shown to provide superior type I error control compared with unharmonized models that include scanner as a covariate. Scanner identity was used as the batch variable, capturing scanner model and head coil differences across sites. The harmonization model included age, sex, study wave, raw BMI, race and ethnicity and socioeconomic status as covariates, with a participant-level random intercept to model the repeated-measures structure and prevent removal of within-person signal. Visual inspection confirmed that scanner-related clustering was substantially reduced after harmonization for both cortical thickness (eFigure 2) and subcortical volume (eFigure 3), while overall distributions and developmental trajectories were preserved.

**eMethods 4. Statistical Analysis Details**

Linear mixed-effects models were fitted with the nlme R package, with nested random intercepts for family and for participant within family. P values for fixed effects were obtained from Wald tests (z = β/SE) with a normal reference distribution, because containment-based denominator degrees of freedom are not well defined when most families include a single child. Likelihood ratio tests compared nested models fitted by maximum likelihood (1 df for age moderation of within-person associations; 3 df for weight status group, group × age, and group × sex); all reported coefficients were estimated by restricted maximum likelihood. The contextual effect (between- minus within-person coefficient) was tested as a linear contrast from the model covariance matrix. Sex-specific slopes were obtained by refitting each sex moderation model with each sex as the reference in turn. In the group × age models, weight status group interacted with linear age, with age² included as a common term; age slopes were converted to SD per year by dividing coefficients per SD of age by the age SD (2.47 years), and group differences at ages 10, 13, and 16 years were estimated as linear combinations of the group and group × age coefficients. Benjamini–Hochberg FDR correction was applied across the 19 regions within each analysis, group, and estimate type, with 2 exceptions: simple slopes were corrected across the regions shown, and sex-specific within-person slopes within weight status groups (eTable 22) were corrected across both sexes (38 tests).

Sensitivity analyses refitted the primary within-group models (1) with an added participant-level random slope for age and (2) after excluding the 6 observations flagged as biologically implausible (CDC modified BMI z-score < −4 or > 8; eMethods 2), recomputing the within- and between-person components while retaining primary group membership. Variance inflation factors were computed from linear-model approximations of the fixed effects of each primary model; for race and ethnicity, the generalized variance inflation factor was converted to the variance inflation factor scale as (GVIF^(1/(2·df)))². Excess residual kurtosis was computed from normalized residuals of the primary mixed-effects models.

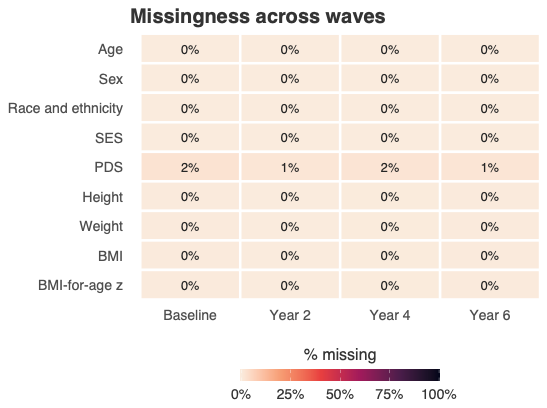

**eFigure 1.** Missingness across variables and waves. Heatmap showing the percentage of missing values for each variable (rows) at each of the 4 imaging waves (columns) in the imaging sample, defined as observations passing T1 quality control with harmonized MRI data (33,257 observations from 11,785 participants), before complete-case exclusion. Missing data were low (≤2% per variable per wave; highest for the Pubertal Development Scale), supporting complete-case analysis.

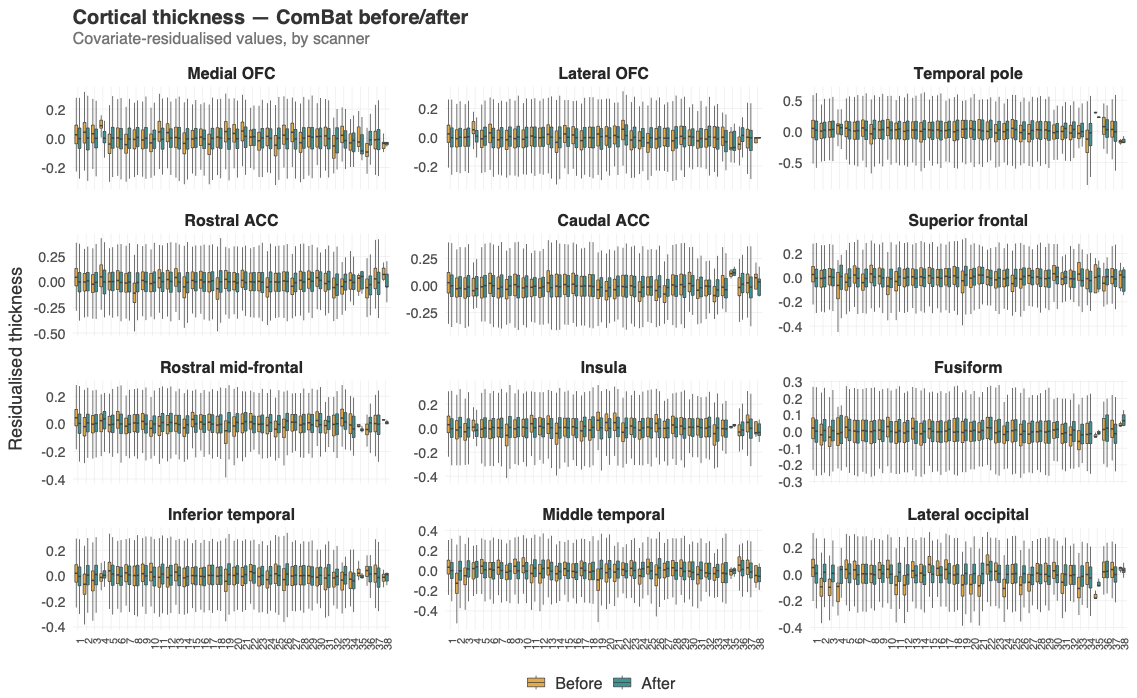

**eFigure 2.** Longitudinal ComBat harmonization of cortical thickness: before and after, by scanner. Box plots show covariate-residualized cortical thickness values for each of the 12 a priori cortical regions across the 34 scanners (x-axis), before (orange) and after (blue) longCombat harmonization. Scanner-related clustering in medians and spread is substantially reduced after harmonization while the overall distribution is preserved.

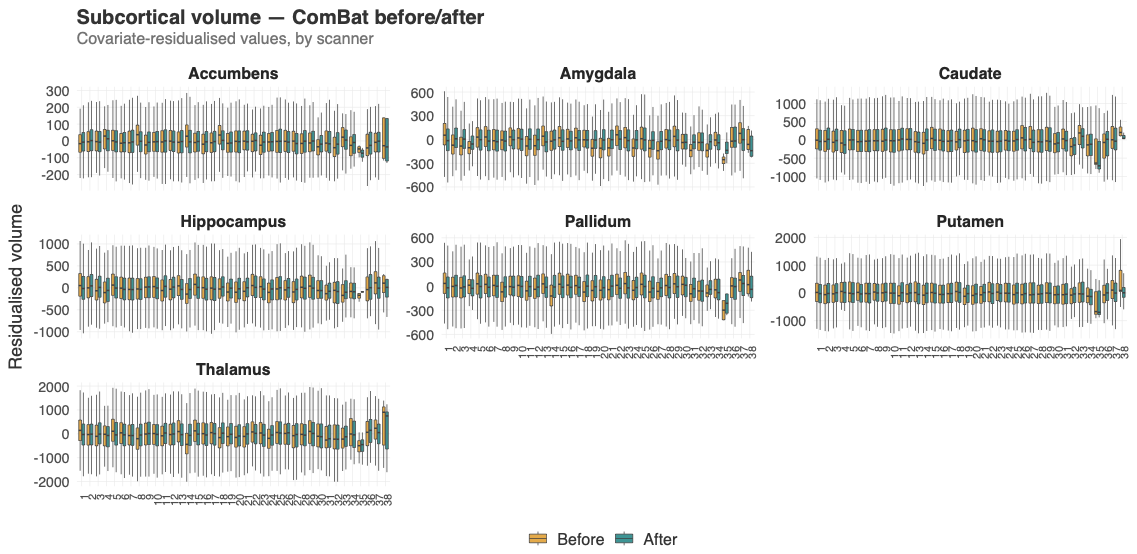

**eFigure 3.** Longitudinal ComBat harmonization of subcortical volume: before and after, by scanner. Box plots show covariate-residualized subcortical volume values (mm³) for each of the 7 a priori subcortical regions across the 34 scanners (x-axis), before (orange) and after (blue) longCombat harmonization. Scanner-related clustering is substantially attenuated after harmonization across all regions.

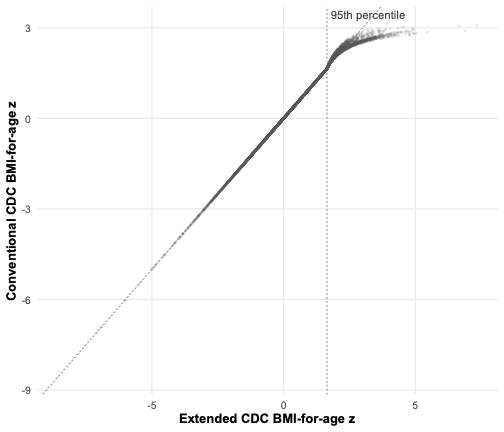

**eFigure 4. Conventional vs extended CDC BMI-for-age z in the analytic sample.** Conventional CDC BMI-for-age z (y-axis) plotted against extended CDC BMI-for-age z (x-axis) for all observations in the analytic sample (32,212 observations). The dashed diagonal line marks identity and the dashed vertical line the 95th percentile (z = 1.645). Below the 95th percentile, the 2 scores are identical by construction. At or above the 95th percentile (5,853 observations from 2,900 participants), conventional z-scores approach an upper limit (range, 1.65 to 3.12), whereas extended z-scores continue to separate higher BMI values (range, 1.65 to 7.32). The correlation between the 2 scores was r = 0.995 (Spearman ρ > 0.999) across all observations and r = 0.904 (ρ = 0.993) at or above the 95th percentile.

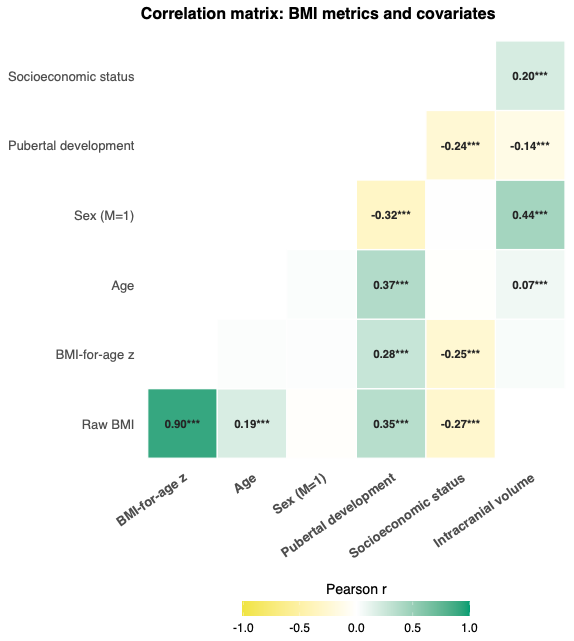

**eFigure 5.** Correlation matrix of raw BMI, extended BMI-for-age z, and model covariates. Lower-triangle Pearson correlations among raw BMI (kg/m²), extended BMI-for-age z, age, sex, pubertal development scale (PDS), socioeconomic status (SES), and intracranial volume (ICV), using one observation per participant (first included visit; n = 11,510). Tiles are colored by correlation magnitude and direction (yellow, negative; green, positive); correlations with |r| < .05 are not labeled. Raw BMI and extended BMI-for-age z are strongly correlated (r = 0.90). Unlike raw BMI, which correlates with age (r = 0.19), BMI-for-age z shows negligible correlation with age and sex (|r| < .05), reflecting CDC standardization, but correlates with PDS (r = 0.28) and SES (r = −0.25), supporting their inclusion as covariates. *P < .05; **P < .01; ***P < .001.

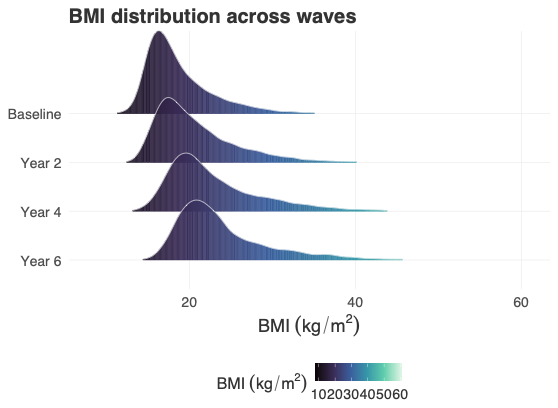

**eFigure 6.** BMI distribution across waves in the analytic sample. Ridge plot of raw BMI (kg/m²) at each imaging wave (baseline, year 2, year 4, and year 6; 32,212 observations), colored by BMI value. The distribution shifts rightward with age, reflecting normal growth across adolescence.

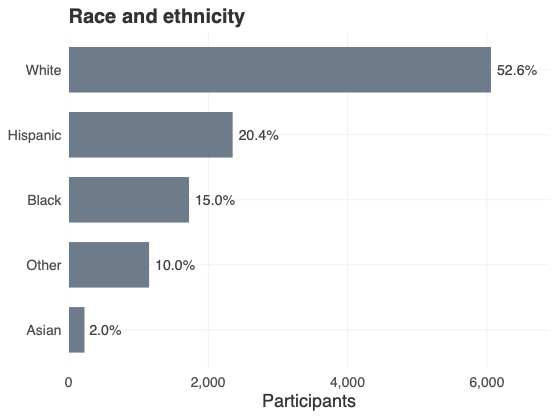

**eFigure 7.** Race and ethnicity of the analytic sample. Horizontal bar chart of the number of participants (n = 11,510) in each caregiver-reported race and ethnicity category, ordered by frequency, with the percentage of the analytic sample labeled on each bar.

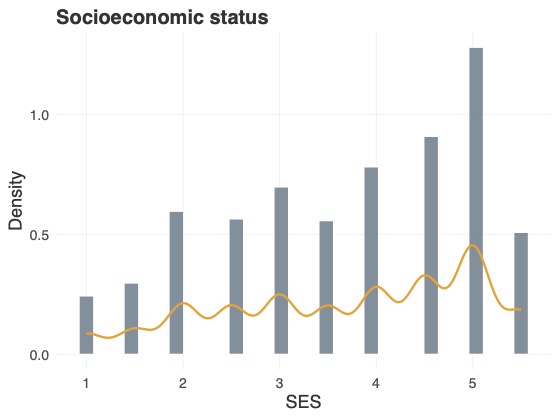

**eFigure 8.** Socioeconomic status distribution of the analytic sample. Histogram with overlaid density curve of the socioeconomic composite (mean of available parental education and household income scores) at each participant's first included visit (n = 11,510).

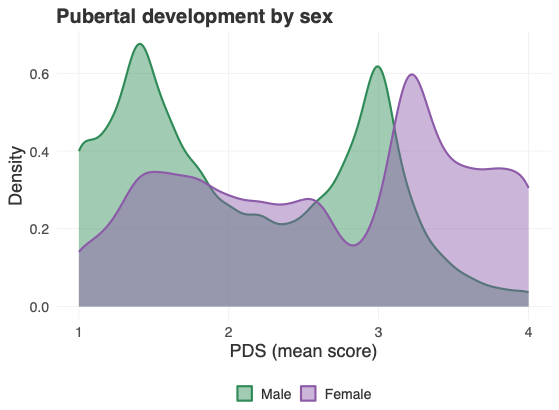

**eFigure 9.** Pubertal development by sex in the analytic sample. Overlaid density plot of mean Pubertal Development Scale score for male and female participants, pooled across all included observations (32,212 observations).

| **eTable 1.** Test of Quadratic Within-Person BMI-for-Age z Term for Nonlinearity of Longitudinal BMI–Brain Associations Within Each CDC Weight Status Group. | | | | | | | |
| --- | --- | --- | --- | --- | --- | --- | --- |
| group | region | β | SE | z | p | q | sig |
| Healthy weight | Rostral mid-frontal | -0.023 | 0.01 | -2.264 | 0.024 | 0.258 |  |
| Healthy weight | Caudal ACC | -0.016 | 0.008 | -1.919 | 0.055 | 0.261 |  |
| Healthy weight | Lateral OFC | -0.013 | 0.011 | -1.172 | 0.241 | 0.796 |  |
| Healthy weight | Temporal pole | -0.007 | 0.014 | -0.541 | 0.588 | 0.796 |  |
| Healthy weight | Rostral ACC | -0.01 | 0.012 | -0.843 | 0.399 | 0.796 |  |
| Healthy weight | Insula | 0.006 | 0.013 | 0.483 | 0.629 | 0.796 |  |
| Healthy weight | Inferior temporal | -0.005 | 0.01 | -0.523 | 0.601 | 0.796 |  |
| Healthy weight | Middle temporal | 0.006 | 0.01 | 0.605 | 0.545 | 0.796 |  |
| Healthy weight | Lateral occipital | 0.006 | 0.01 | 0.584 | 0.559 | 0.796 |  |
| Healthy weight | Fusiform | -0.004 | 0.01 | -0.355 | 0.723 | 0.858 |  |
| Healthy weight | Superior frontal | -0.002 | 0.01 | -0.237 | 0.812 | 0.863 |  |
| Healthy weight | Medial OFC | -0.001 | 0.011 | -0.079 | 0.937 | 0.937 |  |
| Healthy weight | Amygdala | -0.016 | 0.008 | -2.046 | 0.041 | 0.258 |  |
| Healthy weight | Pallidum | -0.021 | 0.01 | -2.153 | 0.031 | 0.258 |  |
| Healthy weight | Accumbens | -0.01 | 0.01 | -1.061 | 0.289 | 0.796 |  |
| Healthy weight | Caudate | -0.004 | 0.004 | -0.817 | 0.414 | 0.796 |  |
| Healthy weight | Hippocampus | -0.003 | 0.005 | -0.501 | 0.617 | 0.796 |  |
| Healthy weight | Putamen | -0.004 | 0.006 | -0.634 | 0.526 | 0.796 |  |
| Healthy weight | Thalamus | 0.002 | 0.007 | 0.23 | 0.818 | 0.863 |  |
| Overweight | Superior frontal | 0.08 | 0.045 | 1.756 | 0.079 | 0.501 |  |
| Overweight | Rostral mid-frontal | 0.096 | 0.046 | 2.07 | 0.038 | 0.501 |  |
| Overweight | Medial OFC | 0.074 | 0.049 | 1.525 | 0.127 | 0.605 |  |
| Overweight | Rostral ACC | -0.041 | 0.05 | -0.81 | 0.418 | 0.738 |  |
| Overweight | Caudal ACC | -0.042 | 0.037 | -1.142 | 0.253 | 0.738 |  |
| Overweight | Insula | -0.055 | 0.053 | -1.04 | 0.298 | 0.738 |  |
| Overweight | Lateral occipital | 0.034 | 0.042 | 0.794 | 0.427 | 0.738 |  |
| Overweight | Inferior temporal | 0.02 | 0.046 | 0.444 | 0.657 | 0.833 |  |
| Overweight | Temporal pole | 0.018 | 0.06 | 0.306 | 0.76 | 0.857 |  |
| Overweight | Middle temporal | -0.014 | 0.046 | -0.296 | 0.767 | 0.857 |  |
| Overweight | Lateral OFC | 0.011 | 0.048 | 0.227 | 0.821 | 0.866 |  |
| Overweight | Fusiform | 0.006 | 0.045 | 0.131 | 0.896 | 0.896 |  |
| Overweight | Putamen | -0.042 | 0.024 | -1.758 | 0.079 | 0.501 |  |
| Overweight | Accumbens | -0.034 | 0.042 | -0.827 | 0.408 | 0.738 |  |
| Overweight | Amygdala | -0.031 | 0.035 | -0.886 | 0.376 | 0.738 |  |
| Overweight | Caudate | -0.017 | 0.019 | -0.878 | 0.38 | 0.738 |  |
| Overweight | Hippocampus | 0.01 | 0.023 | 0.464 | 0.643 | 0.833 |  |
| Overweight | Pallidum | -0.02 | 0.041 | -0.484 | 0.629 | 0.833 |  |
| Overweight | Thalamus | -0.019 | 0.032 | -0.604 | 0.546 | 0.833 |  |
| Obese | Temporal pole | 0.102 | 0.051 | 2.023 | 0.043 | 0.319 |  |
| Obese | Caudal ACC | -0.047 | 0.028 | -1.642 | 0.101 | 0.319 |  |
| Obese | Rostral mid-frontal | 0.063 | 0.037 | 1.677 | 0.094 | 0.319 |  |
| Obese | Superior frontal | 0.056 | 0.036 | 1.547 | 0.122 | 0.331 |  |
| Obese | Lateral OFC | 0.05 | 0.04 | 1.244 | 0.214 | 0.477 |  |
| Obese | Rostral ACC | 0.026 | 0.039 | 0.661 | 0.509 | 0.698 |  |
| Obese | Fusiform | 0.029 | 0.038 | 0.76 | 0.447 | 0.698 |  |
| Obese | Inferior temporal | 0.025 | 0.038 | 0.652 | 0.514 | 0.698 |  |
| Obese | Middle temporal | 0.029 | 0.037 | 0.791 | 0.429 | 0.698 |  |
| Obese | Lateral occipital | 0.019 | 0.034 | 0.571 | 0.568 | 0.72 |  |
| Obese | Medial OFC | -0.005 | 0.039 | -0.123 | 0.902 | 1 |  |
| Obese | Insula | -0.005 | 0.042 | -0.124 | 0.901 | 1 |  |
| Obese | Amygdala | 0.052 | 0.027 | 1.91 | 0.056 | 0.319 |  |
| Obese | Hippocampus | -0.041 | 0.019 | -2.211 | 0.027 | 0.319 |  |
| Obese | Thalamus | -0.043 | 0.025 | -1.745 | 0.081 | 0.319 |  |
| Obese | Accumbens | 0.04 | 0.033 | 1.211 | 0.226 | 0.477 |  |
| Obese | Caudate | 0.017 | 0.016 | 1.078 | 0.281 | 0.534 |  |
| Obese | Pallidum | 0.002 | 0.035 | 0.064 | 0.949 | 1 |  |
| Obese | Putamen | 0 | 0.02 | 0 | 1 | 1 |  |
| Underweight | Medial OFC | 0.019 | 0.014 | 1.376 | 0.169 | 0.927 |  |
| Underweight | Lateral OFC | -0.006 | 0.014 | -0.445 | 0.656 | 0.927 |  |
| Underweight | Rostral ACC | 0.027 | 0.015 | 1.823 | 0.068 | 0.927 |  |
| Underweight | Superior frontal | 0.004 | 0.012 | 0.345 | 0.73 | 0.927 |  |
| Underweight | Rostral mid-frontal | 0.012 | 0.012 | 1.033 | 0.302 | 0.927 |  |
| Underweight | Insula | 0.022 | 0.016 | 1.36 | 0.174 | 0.927 |  |
| Underweight | Fusiform | 0.007 | 0.013 | 0.502 | 0.616 | 0.927 |  |
| Underweight | Inferior temporal | 0.01 | 0.013 | 0.807 | 0.42 | 0.927 |  |
| Underweight | Lateral occipital | -0.003 | 0.012 | -0.278 | 0.781 | 0.927 |  |
| Underweight | Temporal pole | -0.001 | 0.017 | -0.082 | 0.935 | 0.935 |  |
| Underweight | Caudal ACC | 0.002 | 0.011 | 0.144 | 0.885 | 0.935 |  |
| Underweight | Middle temporal | -0.002 | 0.013 | -0.124 | 0.901 | 0.935 |  |
| Underweight | Accumbens | -0.012 | 0.012 | -0.932 | 0.351 | 0.927 |  |
| Underweight | Amygdala | -0.004 | 0.011 | -0.406 | 0.685 | 0.927 |  |
| Underweight | Caudate | -0.006 | 0.006 | -1.118 | 0.264 | 0.927 |  |
| Underweight | Hippocampus | -0.005 | 0.006 | -0.857 | 0.391 | 0.927 |  |
| Underweight | Pallidum | -0.004 | 0.013 | -0.342 | 0.732 | 0.927 |  |
| Underweight | Putamen | -0.004 | 0.008 | -0.475 | 0.635 | 0.927 |  |
| Underweight | Thalamus | -0.004 | 0.009 | -0.4 | 0.689 | 0.927 |  |
| Note. A quadratic within-person BMI-for-age z term was added to the primary within-between model in each CDC weight status group; a significant term would indicate nonlinearity of the longitudinal BMI–brain association. Models were adjusted for age, age², sex, pubertal development, socioeconomic status, race and ethnicity, and intracranial volume (subcortical outcomes), with random intercepts for family and participant. β is the standardized coefficient for the quadratic term. P values are from Wald tests (z = β/SE); FDR correction was applied within each group across 19 regions, and significance markers are based on unrounded *q* values. BMI, body mass index; CDC, Centers for Disease Control and Prevention; FDR, false discovery rate; SE, standard error; *q, FDR-corrected P value*. **q* < .05; ***q* < .01; ****q* < .001. | | | | | | | |

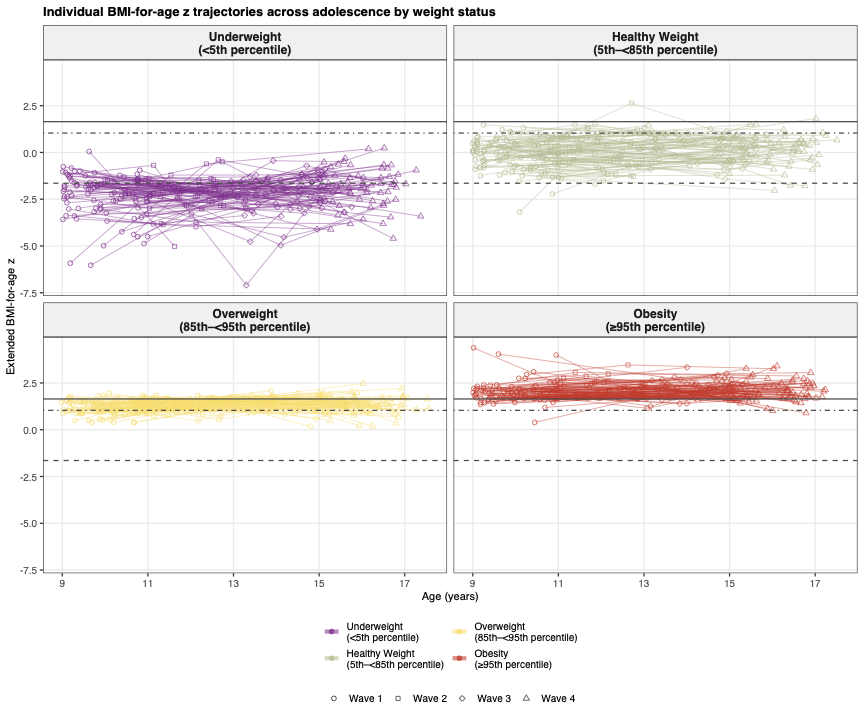

**eFigure 10.** Individual BMI-for-age z trajectories across adolescence by CDC weight status group. Extended BMI-for-age z (y-axis) plotted against age (x-axis) for a random subsample of 100 participants with at least 2 observations in each group (panels) of the analytic sample. Each line represents one participant; point shapes indicate study wave. Horizontal reference lines mark the CDC thresholds defining group boundaries (dashed, z = −1.645; dot-dashed, z = 1.036; solid, z = 1.645). Because the overweight band is narrow, within-person fluctuation of similar magnitude to that in other groups frequently carries individual observations across its boundaries (eFigure 11).

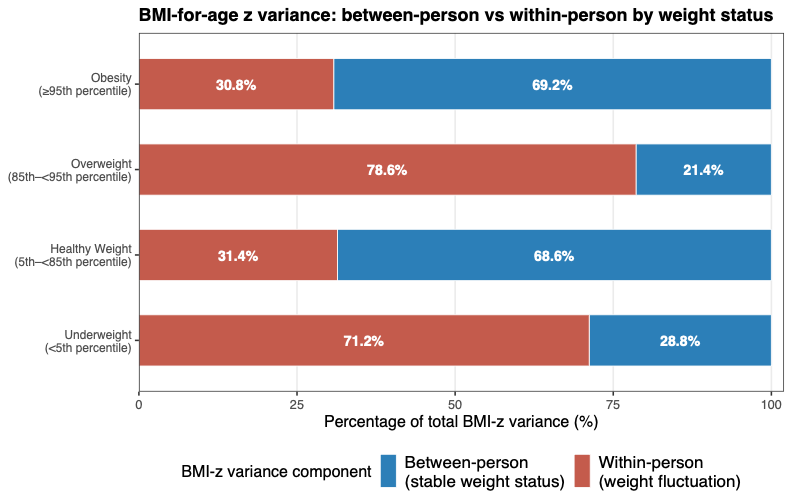
**eFigure 11.** BMI-for-age z variance decomposition by CDC weight status group. Stacked bar chart of the percentage of total extended BMI-for-age z variance attributable to between-person (stable individual differences, blue) and within-person (fluctuation across visits, red) sources in each group. Within-person variation accounted for 31.4% of variance in the healthy weight group, 78.6% in the overweight group, 30.8% in the obesity group, and 71.2% in the underweight group. The high within-person share in the overweight group reflects restricted between-person variance within this narrow category (0.031 vs 0.426 in the healthy weight group) rather than greater fluctuation: absolute within-person variance was 0.195 (healthy weight), 0.115 (overweight), 0.096 (obesity), and 0.667 (underweight).

| **eTable 2. Full Range: Between-Person, Within-Person, and Contextual Associations of Extended BMI-for-Age z With Regional Brain Structure.** | | | | | | | |
| --- | --- | --- | --- | --- | --- | --- | --- |
| estimate type | region | β | SE | z | p | q | sig |
| Between-person | Medial OFC | -0.043 | 0.007 | -6.143 | 0 | 0 | *** |
| Between-person | Lateral OFC | -0.06 | 0.007 | -8.468 | 0 | 0 | *** |
| Between-person | Temporal pole | -0.054 | 0.008 | -7.144 | 0 | 0 | *** |
| Between-person | Superior frontal | -0.068 | 0.007 | -9.094 | 0 | 0 | *** |
| Between-person | Rostral mid-frontal | -0.072 | 0.007 | -10.422 | 0 | 0 | *** |
| Between-person | Middle temporal | -0.042 | 0.008 | -5.512 | 0 | 0 | *** |
| Between-person | Inferior temporal | -0.016 | 0.008 | -2.077 | 0.038 | 0.06 |  |
| Between-person | Fusiform | -0.012 | 0.008 | -1.601 | 0.109 | 0.148 |  |
| Between-person | Rostral ACC | -0.004 | 0.008 | -0.545 | 0.586 | 0.619 |  |
| Between-person | Caudal ACC | -0.005 | 0.008 | -0.588 | 0.556 | 0.619 |  |
| Between-person | Lateral occipital | 0.005 | 0.007 | 0.636 | 0.525 | 0.619 |  |
| Between-person | Insula | 0.001 | 0.007 | 0.119 | 0.906 | 0.906 |  |
| Between-person | Accumbens | 0.03 | 0.007 | 4.37 | 0 | 0 | *** |
| Between-person | Hippocampus | 0.026 | 0.007 | 3.809 | 0 | 0 | *** |
| Between-person | Pallidum | 0.049 | 0.007 | 7.519 | 0 | 0 | *** |
| Between-person | Putamen | 0.03 | 0.007 | 4.051 | 0 | 0 | *** |
| Between-person | Thalamus | 0.048 | 0.006 | 7.654 | 0 | 0 | *** |
| Between-person | Caudate | 0.016 | 0.008 | 1.996 | 0.046 | 0.067 |  |
| Between-person | Amygdala | 0.005 | 0.007 | 0.692 | 0.489 | 0.619 |  |
| Within-person | Lateral occipital | 0.022 | 0.006 | 3.583 | 0 | 0.002 | ** |
| Within-person | Middle temporal | 0.021 | 0.006 | 3.173 | 0.002 | 0.007 | ** |
| Within-person | Temporal pole | -0.028 | 0.01 | -2.899 | 0.004 | 0.014 | * |
| Within-person | Superior frontal | -0.014 | 0.007 | -2.059 | 0.04 | 0.107 |  |
| Within-person | Rostral mid-frontal | -0.013 | 0.007 | -1.865 | 0.062 | 0.131 |  |
| Within-person | Lateral OFC | -0.012 | 0.007 | -1.655 | 0.098 | 0.186 |  |
| Within-person | Fusiform | 0.008 | 0.007 | 1.198 | 0.231 | 0.399 |  |
| Within-person | Inferior temporal | 0.006 | 0.007 | 0.941 | 0.347 | 0.549 |  |
| Within-person | Medial OFC | 0.006 | 0.007 | 0.793 | 0.428 | 0.552 |  |
| Within-person | Insula | -0.007 | 0.009 | -0.821 | 0.412 | 0.552 |  |
| Within-person | Caudal ACC | 0.001 | 0.005 | 0.22 | 0.826 | 0.923 |  |
| Within-person | Rostral ACC | 0 | 0.008 | 0.023 | 0.981 | 0.981 |  |
| Within-person | Hippocampus | 0.017 | 0.003 | 5.266 | 0 | 0 | *** |
| Within-person | Thalamus | 0.023 | 0.005 | 5.1 | 0 | 0 | *** |
| Within-person | Pallidum | 0.015 | 0.006 | 2.407 | 0.016 | 0.051 |  |
| Within-person | Putamen | 0.007 | 0.003 | 1.943 | 0.052 | 0.124 |  |
| Within-person | Accumbens | -0.005 | 0.006 | -0.779 | 0.436 | 0.552 |  |
| Within-person | Caudate | 0.001 | 0.003 | 0.389 | 0.698 | 0.828 |  |
| Within-person | Amygdala | 0 | 0.005 | -0.073 | 0.942 | 0.981 |  |
| Contextual | Medial OFC | -0.049 | 0.01 | -4.827 | 0 | 0 | *** |
| Contextual | Lateral OFC | -0.048 | 0.01 | -4.758 | 0 | 0 | *** |
| Contextual | Superior frontal | -0.054 | 0.01 | -5.468 | 0 | 0 | *** |
| Contextual | Rostral mid-frontal | -0.059 | 0.01 | -6.159 | 0 | 0 | *** |
| Contextual | Middle temporal | -0.063 | 0.01 | -6.295 | 0 | 0 | *** |
| Contextual | Inferior temporal | -0.022 | 0.01 | -2.201 | 0.028 | 0.053 |  |
| Contextual | Temporal pole | -0.026 | 0.012 | -2.147 | 0.032 | 0.055 |  |
| Contextual | Fusiform | -0.02 | 0.01 | -2.008 | 0.045 | 0.071 |  |
| Contextual | Lateral occipital | -0.017 | 0.01 | -1.804 | 0.071 | 0.104 |  |
| Contextual | Insula | 0.008 | 0.011 | 0.706 | 0.48 | 0.57 |  |
| Contextual | Caudal ACC | -0.006 | 0.01 | -0.617 | 0.537 | 0.578 |  |
| Contextual | Rostral ACC | -0.004 | 0.011 | -0.4 | 0.689 | 0.689 |  |
| Contextual | Accumbens | 0.035 | 0.009 | 3.789 | 0 | 0 | *** |
| Contextual | Pallidum | 0.033 | 0.009 | 3.682 | 0 | 0.001 | *** |
| Contextual | Thalamus | 0.024 | 0.008 | 3.141 | 0.002 | 0.004 | ** |
| Contextual | Putamen | 0.023 | 0.008 | 2.856 | 0.004 | 0.009 | ** |
| Contextual | Caudate | 0.015 | 0.008 | 1.766 | 0.077 | 0.105 |  |
| Contextual | Hippocampus | 0.009 | 0.008 | 1.218 | 0.223 | 0.283 |  |
| Contextual | Amygdala | 0.005 | 0.008 | 0.601 | 0.548 | 0.578 |  |
| Note. The within-between model was fitted across all participants regardless of CDC weight status group. Between-person and within-person rows give the respective coefficients; contextual rows give the between- minus within-person difference, estimated as a linear contrast from the model covariance matrix. Models were adjusted for age, age², sex, pubertal development, socioeconomic status, race and ethnicity, and intracranial volume (subcortical outcomes), with random intercepts for family and participant. β is the standardized coefficient (SD change in the outcome per 1-SD increase in BMI-for-age z). Because between-person associations were nonlinear (eTable 3), these linear estimates average across the weight range; group-specific estimates are in eTable 17. P values are from Wald tests (z = β/SE); FDR correction was applied within each estimate type across 19 regions, and significance markers are based on unrounded q values. BMI, body mass index; CDC, Centers for Disease Control and Prevention; FDR, false discovery rate; SE, standard error; q, FDR-corrected P value. *q < .05; **q < .01; ***q < .001. | | | | | | | |

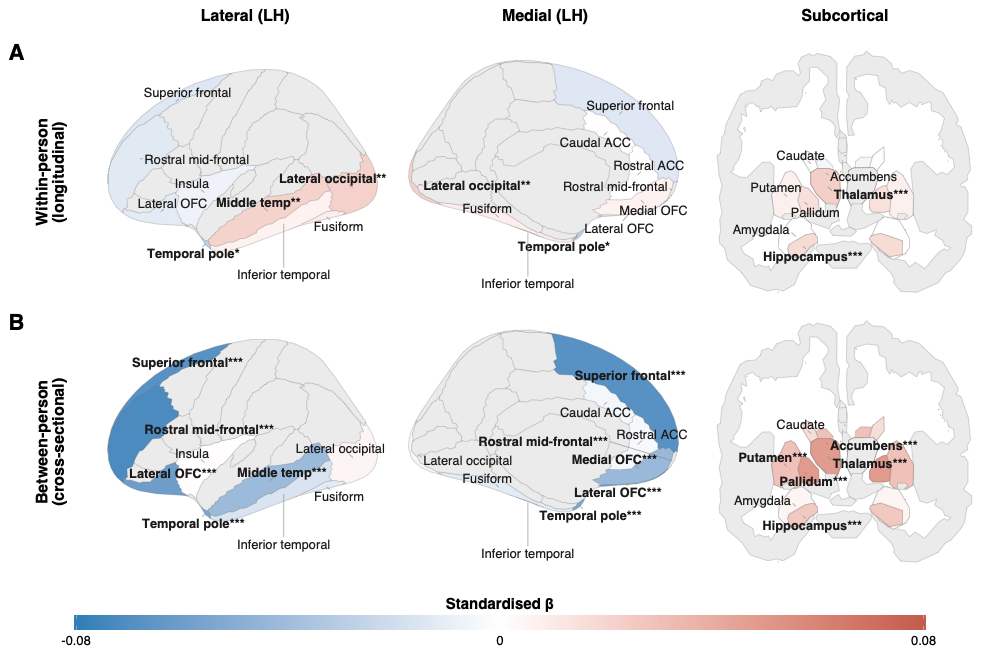
**eFigure 12**. Between- and within-person associations of extended BMI-for-age z with regional brain structure across the full BMI range. Standardized β coefficients per 1-SD increase in extended BMI-for-age z mapped to left-hemisphere cortical surfaces (lateral and medial views) and a subcortical coronal section, from the within-between model fitted across all participants: (A) within-person (longitudinal) associations; (B) between-person (cross-sectional) associations. Models were adjusted for age, age², sex, pubertal development, socioeconomic status, race and ethnicity, and intracranial volume (subcortical outcomes). Blue indicates lower cortical thickness or subcortical volume at higher BMI-for-age z; red indicates higher. All 19 analyzed regions are colored by β; bold labels with asterisks denote FDR-significant associations (*q < .05, **q < .01, ***q < .001), and regions not analyzed are shown in gray. The color scale (−0.08 to 0.08) is derived from these estimates and differs from the main-text figures. Because associations were nonlinear (eTable 3), these linear full-range estimates should be read alongside the group-specific estimates in Figures 3 and 4; full estimates are in eTable 2.

| **eTable 3. Full Range: Test of Quadratic Between-Person and Within-Person BMI-for-Age z Terms.** | | | | | | | |
| --- | --- | --- | --- | --- | --- | --- | --- |
| estimate type | region | β | SE | z | p | q | sig |
| Between-person quadratic | Medial OFC | -0.02 | 0.004 | -4.995 | 0 | 0 | *** |
| Between-person quadratic | Lateral OFC | -0.024 | 0.004 | -6.14 | 0 | 0 | *** |
| Between-person quadratic | Temporal pole | -0.021 | 0.004 | -4.923 | 0 | 0 | *** |
| Between-person quadratic | Rostral ACC | -0.016 | 0.004 | -3.755 | 0 | 0 | *** |
| Between-person quadratic | Superior frontal | -0.018 | 0.004 | -4.247 | 0 | 0 | *** |
| Between-person quadratic | Rostral mid-frontal | -0.017 | 0.004 | -4.491 | 0 | 0 | *** |
| Between-person quadratic | Insula | -0.018 | 0.004 | -4.488 | 0 | 0 | *** |
| Between-person quadratic | Fusiform | -0.032 | 0.004 | -7.525 | 0 | 0 | *** |
| Between-person quadratic | Inferior temporal | -0.027 | 0.004 | -6.341 | 0 | 0 | *** |
| Between-person quadratic | Middle temporal | -0.031 | 0.004 | -7.39 | 0 | 0 | *** |
| Between-person quadratic | Lateral occipital | -0.026 | 0.004 | -6.252 | 0 | 0 | *** |
| Between-person quadratic | Caudal ACC | -0.008 | 0.005 | -1.674 | 0.094 | 0.099 |  |
| Between-person quadratic | Amygdala | -0.015 | 0.004 | -4.094 | 0 | 0 | *** |
| Between-person quadratic | Hippocampus | -0.015 | 0.004 | -4.014 | 0 | 0 | *** |
| Between-person quadratic | Thalamus | -0.014 | 0.003 | -4.109 | 0 | 0 | *** |
| Between-person quadratic | Caudate | -0.015 | 0.004 | -3.451 | 0.001 | 0.001 | *** |
| Between-person quadratic | Putamen | -0.011 | 0.004 | -2.813 | 0.005 | 0.006 | ** |
| Between-person quadratic | Pallidum | -0.008 | 0.004 | -2.154 | 0.031 | 0.035 | * |
| Between-person quadratic | Accumbens | -0.004 | 0.004 | -1.181 | 0.238 | 0.238 |  |
| Within-person quadratic | Lateral OFC | -0.014 | 0.008 | -1.724 | 0.085 | 0.53 |  |
| Within-person quadratic | Caudal ACC | -0.011 | 0.006 | -1.897 | 0.058 | 0.53 |  |
| Within-person quadratic | Medial OFC | 0.002 | 0.008 | 0.288 | 0.773 | 0.858 |  |
| Within-person quadratic | Temporal pole | -0.009 | 0.01 | -0.883 | 0.377 | 0.858 |  |
| Within-person quadratic | Rostral ACC | -0.002 | 0.008 | -0.255 | 0.798 | 0.858 |  |
| Within-person quadratic | Rostral mid-frontal | -0.004 | 0.007 | -0.515 | 0.606 | 0.858 |  |
| Within-person quadratic | Insula | 0.004 | 0.009 | 0.411 | 0.681 | 0.858 |  |
| Within-person quadratic | Fusiform | 0.002 | 0.007 | 0.274 | 0.784 | 0.858 |  |
| Within-person quadratic | Inferior temporal | -0.002 | 0.007 | -0.237 | 0.813 | 0.858 |  |
| Within-person quadratic | Middle temporal | 0.004 | 0.007 | 0.605 | 0.545 | 0.858 |  |
| Within-person quadratic | Lateral occipital | 0.002 | 0.007 | 0.343 | 0.731 | 0.858 |  |
| Within-person quadratic | Superior frontal | -0.001 | 0.007 | -0.068 | 0.945 | 0.945 |  |
| Within-person quadratic | Amygdala | -0.009 | 0.006 | -1.591 | 0.112 | 0.53 |  |
| Within-person quadratic | Pallidum | -0.015 | 0.007 | -2.113 | 0.035 | 0.53 |  |
| Within-person quadratic | Caudate | -0.004 | 0.003 | -1.265 | 0.206 | 0.782 |  |
| Within-person quadratic | Accumbens | -0.007 | 0.007 | -1.063 | 0.288 | 0.858 |  |
| Within-person quadratic | Hippocampus | -0.003 | 0.004 | -0.9 | 0.368 | 0.858 |  |
| Within-person quadratic | Putamen | -0.002 | 0.004 | -0.62 | 0.536 | 0.858 |  |
| Within-person quadratic | Thalamus | -0.003 | 0.005 | -0.529 | 0.597 | 0.858 |  |
| Note. Quadratic between-person (wave-mean squared) and within-person (deviation squared) BMI-for-age z terms were added separately to the full-range model described in eTable 2, with the same covariates and random effects. A significant negative quadratic between-person term indicates that the cross-sectional association becomes more negative (cortical thickness) or weaker (subcortical volume) at higher BMI-for-age z, consistent with the group-specific associations in eTable 17. β is the standardized coefficient for the quadratic term. P values are from Wald tests (z = β/SE); FDR correction was applied within each term across 19 regions, and significance markers are based on unrounded q values. BMI, body mass index; CDC, Centers for Disease Control and Prevention; FDR, false discovery rate; SE, standard error; q, FDR-corrected P value. *q < .05; **q < .01; ***q < .001. | | | | | | | |

| **eTable 4. Likelihood Ratio Tests for Age Moderation of Within-Person BMI-for-Age z Associations With Regional Brain Structure Across the Full BMI Range.** | | | | | | |
| --- | --- | --- | --- | --- | --- | --- |
| measure | region | LR χ² | df | p | q | sig |
| Cortical thickness | Superior frontal | 22.97 | 1 | 0 | 0 | *** |
| Cortical thickness | Middle temporal | 29.62 | 1 | 0 | 0 | *** |
| Cortical thickness | Rostral mid-frontal | 13.21 | 1 | 0 | 0.001 | ** |
| Cortical thickness | Inferior temporal | 12.88 | 1 | 0 | 0.001 | ** |
| Cortical thickness | Lateral occipital | 13.27 | 1 | 0 | 0.001 | ** |
| Cortical thickness | Medial OFC | 8.95 | 1 | 0.003 | 0.009 | ** |
| Cortical thickness | Fusiform | 6.05 | 1 | 0.014 | 0.038 | * |
| Cortical thickness | Caudal ACC | 4.89 | 1 | 0.027 | 0.064 |  |
| Cortical thickness | Lateral OFC | 2.36 | 1 | 0.124 | 0.219 |  |
| Cortical thickness | Rostral ACC | 0.84 | 1 | 0.36 | 0.488 |  |
| Cortical thickness | Temporal pole | 0.6 | 1 | 0.438 | 0.521 |  |
| Cortical thickness | Insula | 0.03 | 1 | 0.859 | 0.876 |  |
| Subcortical volume | Hippocampus | 3.94 | 1 | 0.047 | 0.099 |  |
| Subcortical volume | Caudate | 2.33 | 1 | 0.127 | 0.219 |  |
| Subcortical volume | Thalamus | 1.85 | 1 | 0.173 | 0.275 |  |
| Subcortical volume | Pallidum | 1.41 | 1 | 0.234 | 0.343 |  |
| Subcortical volume | Putamen | 0.71 | 1 | 0.398 | 0.504 |  |
| Subcortical volume | Accumbens | 0.34 | 1 | 0.559 | 0.624 |  |
| Subcortical volume | Amygdala | 0.02 | 1 | 0.876 | 0.876 |  |
| Note. Age moderation was tested in all participants by comparing models with and without a within-person BMI-for-age z × age interaction using likelihood ratio tests (1 df), specified as in the within-group models in eTable 19. Both models were fitted by maximum likelihood, included the between-person component, and were adjusted for age², sex, pubertal development, socioeconomic status, race and ethnicity, and intracranial volume (subcortical outcomes), with random intercepts for family and participant. LR χ² is the likelihood ratio statistic. FDR correction was applied across 19 regions, and significance markers are based on unrounded q values. BMI, body mass index; FDR, false discovery rate; LR, likelihood ratio; q, FDR-corrected P value. *q < .05; **q < .01; ***q < .001. | | | | | | |

| **eTable 5. Simple Slopes of Within-Person BMI-for-Age z Associations With Cortical Thickness at Ages 10, 13, and 16 Years Across the Full BMI Range.** | | | | | | | | |
| --- | --- | --- | --- | --- | --- | --- | --- | --- |
| group | region | age | β | SE | z | p | q | sig |
| All participants | Fusiform | 10 | -0.015 | 0.012 | -1.334 | 0.182 | 0.212 |  |
| All participants | Fusiform | 13 | 0.01 | 0.007 | 1.471 | 0.141 | 0.208 |  |
| All participants | Fusiform | 16 | 0.035 | 0.013 | 2.725 | 0.006 | 0.007 | ** |
| All participants | Inferior temporal | 10 | -0.028 | 0.012 | -2.41 | 0.016 | 0.028 | * |
| All participants | Inferior temporal | 13 | 0.009 | 0.007 | 1.345 | 0.179 | 0.208 |  |
| All participants | Inferior temporal | 16 | 0.045 | 0.013 | 3.562 | 0 | 0.001 | *** |
| All participants | Lateral occipital | 10 | -0.01 | 0.011 | -0.956 | 0.339 | 0.339 |  |
| All participants | Lateral occipital | 13 | 0.025 | 0.006 | 3.978 | 0 | 0 | *** |
| All participants | Lateral occipital | 16 | 0.06 | 0.012 | 4.964 | 0 | 0 | *** |
| All participants | Medial OFC | 10 | -0.025 | 0.013 | -1.974 | 0.048 | 0.068 |  |
| All participants | Medial OFC | 13 | 0.008 | 0.007 | 1.122 | 0.262 | 0.262 |  |
| All participants | Medial OFC | 16 | 0.042 | 0.014 | 2.963 | 0.003 | 0.004 | ** |
| All participants | Middle temporal | 10 | -0.03 | 0.011 | -2.662 | 0.008 | 0.018 | * |
| All participants | Middle temporal | 13 | 0.025 | 0.007 | 3.777 | 0 | 0.001 | *** |
| All participants | Middle temporal | 16 | 0.08 | 0.013 | 6.301 | 0 | 0 | *** |
| All participants | Rostral mid-frontal | 10 | -0.048 | 0.012 | -4.044 | 0 | 0 | *** |
| All participants | Rostral mid-frontal | 13 | -0.01 | 0.007 | -1.444 | 0.149 | 0.208 |  |
| All participants | Rostral mid-frontal | 16 | 0.028 | 0.013 | 2.134 | 0.033 | 0.033 | * |
| All participants | Superior frontal | 10 | -0.059 | 0.012 | -5.11 | 0 | 0 | *** |
| All participants | Superior frontal | 13 | -0.01 | 0.007 | -1.5 | 0.134 | 0.208 |  |
| All participants | Superior frontal | 16 | 0.039 | 0.013 | 3.047 | 0.002 | 0.004 | ** |
| Note. Simple slopes are within-person BMI-for-age z associations at ages 10, 13, and 16 years in all participants, estimated from the age moderation models (restricted maximum likelihood) as linear combinations of the within-person and interaction coefficients. Only regions with significant age moderation in eTable 4 (q < .05) are shown; all were cortical. Models were adjusted as described in eTable 4. β is the standardized within-person coefficient at the indicated age. P values are from Wald tests (z = β/SE); FDR correction was applied within each age across the regions shown, and significance markers are based on unrounded q values. BMI, body mass index; FDR, false discovery rate; SE, standard error; q, FDR-corrected P value. *q < .05; **q < .01; ***q < .001. | | | | | | | | |

| **eTable 6. Within-Person × Sex Interaction Terms for BMI-for-Age z Associations With Brain Structure Across the Full BMI Range.** | | | | | | | |
| --- | --- | --- | --- | --- | --- | --- | --- |
| group | region | β | SE | z | p | q | sig |
| All participants | Superior frontal | -0.063 | 0.013 | -4.767 | 0 | 0 | *** |
| All participants | Fusiform | -0.08 | 0.013 | -6.078 | 0 | 0 | *** |
| All participants | Inferior temporal | -0.066 | 0.013 | -5.048 | 0 | 0 | *** |
| All participants | Middle temporal | -0.065 | 0.013 | -5.049 | 0 | 0 | *** |
| All participants | Lateral occipital | -0.045 | 0.012 | -3.711 | 0 | 0 | *** |
| All participants | Caudal ACC | -0.037 | 0.01 | -3.589 | 0 | 0.001 | *** |
| All participants | Rostral mid-frontal | -0.043 | 0.014 | -3.145 | 0.002 | 0.003 | ** |
| All participants | Insula | -0.045 | 0.017 | -2.638 | 0.008 | 0.012 | * |
| All participants | Lateral OFC | -0.03 | 0.014 | -2.059 | 0.04 | 0.05 |  |
| All participants | Medial OFC | -0.03 | 0.015 | -2.013 | 0.044 | 0.052 |  |
| All participants | Temporal pole | -0.037 | 0.019 | -1.914 | 0.056 | 0.062 |  |
| All participants | Rostral ACC | -0.015 | 0.015 | -0.94 | 0.347 | 0.367 |  |
| All participants | Caudate | -0.026 | 0.005 | -4.857 | 0 | 0 | *** |
| All participants | Hippocampus | -0.048 | 0.006 | -7.557 | 0 | 0 | *** |
| All participants | Putamen | -0.032 | 0.007 | -4.662 | 0 | 0 | *** |
| All participants | Amygdala | -0.036 | 0.01 | -3.617 | 0 | 0.001 | *** |
| All participants | Accumbens | -0.037 | 0.012 | -2.985 | 0.003 | 0.004 | ** |
| All participants | Thalamus | -0.022 | 0.009 | -2.422 | 0.015 | 0.021 | * |
| All participants | Pallidum | -0.009 | 0.013 | -0.729 | 0.466 | 0.466 |  |
| Note. Within-person BMI-for-age z × sex interactions (male participants as reference) were added to the full-range within-between model in all participants, specified as in the within-group models in eTable 21. Models were adjusted for age, age², pubertal development, socioeconomic status, race and ethnicity, and intracranial volume (subcortical outcomes), with random intercepts for family and participant. Negative coefficients indicate a more negative within-person slope in female than in male participants. P values are from Wald tests (z = β/SE); FDR correction was applied across 19 regions, and significance markers are based on unrounded q values. BMI, body mass index; FDR, false discovery rate; SE, standard error; q, FDR-corrected P value. *q < .05; **q < .01; ***q < .001. | | | | | | | |

| **eTable 7. Sex-Specific Within-Person BMI-for-Age z Associations With Brain Structure Across the Full BMI Range.** | | | | | | | | |
| --- | --- | --- | --- | --- | --- | --- | --- | --- |
| contrast | sex | region | β | SE | z | p | q | sig |
| Within-person slope | Female | Superior frontal | -0.05 | 0.01 | -4.954 | 0 | 0 | *** |
| Within-person slope | Female | Fusiform | -0.038 | 0.01 | -3.816 | 0 | 0.001 | ** |
| Within-person slope | Female | Rostral mid-frontal | -0.037 | 0.01 | -3.6 | 0 | 0.002 | ** |
| Within-person slope | Female | Temporal pole | -0.049 | 0.015 | -3.344 | 0.001 | 0.003 | ** |
| Within-person slope | Female | Inferior temporal | -0.032 | 0.01 | -3.205 | 0.001 | 0.004 | ** |
| Within-person slope | Female | Lateral OFC | -0.029 | 0.011 | -2.64 | 0.008 | 0.018 | * |
| Within-person slope | Female | Caudal ACC | -0.02 | 0.008 | -2.573 | 0.01 | 0.019 | * |
| Within-person slope | Female | Insula | -0.033 | 0.013 | -2.532 | 0.011 | 0.02 | * |
| Within-person slope | Female | Middle temporal | -0.017 | 0.01 | -1.746 | 0.081 | 0.11 |  |
| Within-person slope | Female | Medial OFC | -0.011 | 0.011 | -1.004 | 0.315 | 0.352 |  |
| Within-person slope | Female | Rostral ACC | -0.008 | 0.012 | -0.696 | 0.487 | 0.514 |  |
| Within-person slope | Female | Lateral occipital | -0.004 | 0.009 | -0.466 | 0.641 | 0.641 |  |
| Within-person slope | Female | Caudate | -0.014 | 0.004 | -3.423 | 0.001 | 0.003 | ** |
| Within-person slope | Female | Accumbens | -0.026 | 0.009 | -2.768 | 0.006 | 0.013 | * |
| Within-person slope | Female | Amygdala | -0.021 | 0.008 | -2.785 | 0.005 | 0.013 | * |
| Within-person slope | Female | Hippocampus | -0.011 | 0.005 | -2.273 | 0.023 | 0.035 | * |
| Within-person slope | Female | Putamen | -0.012 | 0.005 | -2.258 | 0.024 | 0.035 | * |
| Within-person slope | Female | Thalamus | 0.011 | 0.007 | 1.501 | 0.133 | 0.169 |  |
| Within-person slope | Female | Pallidum | 0.01 | 0.01 | 1.022 | 0.307 | 0.352 |  |
| Within-person slope | Male | Fusiform | 0.041 | 0.009 | 4.813 | 0 | 0 | *** |
| Within-person slope | Male | Inferior temporal | 0.034 | 0.009 | 3.955 | 0 | 0 | *** |
| Within-person slope | Male | Middle temporal | 0.048 | 0.008 | 5.672 | 0 | 0 | *** |
| Within-person slope | Male | Lateral occipital | 0.041 | 0.008 | 5.129 | 0 | 0 | *** |
| Within-person slope | Male | Caudal ACC | 0.017 | 0.007 | 2.465 | 0.014 | 0.029 | * |
| Within-person slope | Male | Medial OFC | 0.018 | 0.01 | 1.899 | 0.058 | 0.091 |  |
| Within-person slope | Male | Superior frontal | 0.013 | 0.009 | 1.47 | 0.142 | 0.207 |  |
| Within-person slope | Male | Insula | 0.012 | 0.011 | 1.061 | 0.289 | 0.366 |  |
| Within-person slope | Male | Temporal pole | -0.012 | 0.013 | -0.997 | 0.319 | 0.379 |  |
| Within-person slope | Male | Rostral ACC | 0.006 | 0.01 | 0.62 | 0.535 | 0.592 |  |
| Within-person slope | Male | Rostral mid-frontal | 0.005 | 0.009 | 0.582 | 0.561 | 0.592 |  |
| Within-person slope | Male | Lateral OFC | 0 | 0.009 | 0.048 | 0.962 | 0.962 |  |
| Within-person slope | Male | Hippocampus | 0.037 | 0.004 | 8.891 | 0 | 0 | *** |
| Within-person slope | Male | Putamen | 0.02 | 0.004 | 4.478 | 0 | 0 | *** |
| Within-person slope | Male | Thalamus | 0.033 | 0.006 | 5.469 | 0 | 0 | *** |
| Within-person slope | Male | Caudate | 0.012 | 0.003 | 3.406 | 0.001 | 0.002 | ** |
| Within-person slope | Male | Pallidum | 0.019 | 0.008 | 2.315 | 0.021 | 0.039 | * |
| Within-person slope | Male | Amygdala | 0.015 | 0.007 | 2.26 | 0.024 | 0.041 | * |
| Within-person slope | Male | Accumbens | 0.011 | 0.008 | 1.314 | 0.189 | 0.256 |  |
| Note. Sex-specific within-person slopes were estimated by refitting each sex moderation model (eTable 6) with each sex as the reference in turn. β is the standardized within-person coefficient within the indicated sex. P values are from Wald tests (z = β/SE); FDR correction was applied within each sex across 19 regions, and significance markers are based on unrounded q values. BMI, body mass index; FDR, false discovery rate; SE, standard error; q, FDR-corrected P value. *q < .05; **q < .01; ***q < .001. | | | | | | | | |

| **eTable 8. Likelihood Ratio Tests for Differences in Regional Brain Structure Between CDC Weight Status Groups.** | | | | | | |
| --- | --- | --- | --- | --- | --- | --- |
| test | region | LR χ² | df | p | q | sig |
| Group | Medial OFC | 45.32 | 3 | 0 | 0 | *** |
| Group | Lateral OFC | 80.7 | 3 | 0 | 0 | *** |
| Group | Temporal pole | 48.77 | 3 | 0 | 0 | *** |
| Group | Superior frontal | 81.18 | 3 | 0 | 0 | *** |
| Group | Rostral mid-frontal | 110.62 | 3 | 0 | 0 | *** |
| Group | Fusiform | 20.97 | 3 | 0 | 0 | *** |
| Group | Middle temporal | 61.75 | 3 | 0 | 0 | *** |
| Group | Inferior temporal | 12.35 | 3 | 0.006 | 0.01 | ** |
| Group | Lateral occipital | 11.89 | 3 | 0.008 | 0.011 | * |
| Group | Insula | 9.02 | 3 | 0.029 | 0.037 | * |
| Group | Rostral ACC | 8.29 | 3 | 0.04 | 0.045 | * |
| Group | Caudal ACC | 3.71 | 3 | 0.294 | 0.294 |  |
| Group | Pallidum | 37 | 3 | 0 | 0 | *** |
| Group | Thalamus | 36.19 | 3 | 0 | 0 | *** |
| Group | Hippocampus | 19.06 | 3 | 0 | 0.001 | *** |
| Group | Putamen | 14.37 | 3 | 0.002 | 0.004 | ** |
| Group | Amygdala | 10.36 | 3 | 0.016 | 0.021 | * |
| Group | Accumbens | 8.66 | 3 | 0.034 | 0.041 | * |
| Group | Caudate | 8.1 | 3 | 0.044 | 0.047 | * |
| Note. Differences between the 4 CDC weight status groups were tested across all participants by comparing models with and without weight status group (3 df) using likelihood ratio tests, with both models fitted by maximum likelihood. Groups were defined by each participant’s mean extended BMI-for-age z across included visits. Models were adjusted for age, age², sex, pubertal development, socioeconomic status, race and ethnicity, and intracranial volume (subcortical outcomes), with random intercepts for family and participant. LR χ² is the likelihood ratio statistic. FDR correction was applied across 19 regions, and significance markers are based on unrounded q values. BMI, body mass index; CDC, Centers for Disease Control and Prevention; FDR, false discovery rate; LR, likelihood ratio; q, FDR-corrected P value. *q < .05; **q < .01; ***q < .001. | | | | | | |

| **eTable 9. Differences in Regional Brain Structure Between Each CDC Weight Status Group and the Healthy Weight Group.** | | | | | | | |
| --- | --- | --- | --- | --- | --- | --- | --- |
| group | region | difference | SE | z | p | q | sig |
| Overweight | Superior frontal | -0.114 | 0.021 | -5.373 | 0 | 0 | *** |
| Overweight | Rostral mid-frontal | -0.108 | 0.02 | -5.495 | 0 | 0 | *** |
| Overweight | Temporal pole | -0.06 | 0.021 | -2.776 | 0.006 | 0.021 | * |
| Overweight | Middle temporal | -0.059 | 0.022 | -2.716 | 0.007 | 0.021 | * |
| Overweight | Lateral OFC | -0.038 | 0.02 | -1.863 | 0.062 | 0.145 |  |
| Overweight | Insula | 0.034 | 0.021 | 1.661 | 0.097 | 0.167 |  |
| Overweight | Medial OFC | -0.032 | 0.02 | -1.598 | 0.11 | 0.174 |  |
| Overweight | Rostral ACC | 0.023 | 0.022 | 1.082 | 0.279 | 0.408 |  |
| Overweight | Inferior temporal | -0.017 | 0.022 | -0.809 | 0.419 | 0.539 |  |
| Overweight | Lateral occipital | 0.012 | 0.021 | 0.569 | 0.569 | 0.636 |  |
| Overweight | Caudal ACC | -0.009 | 0.023 | -0.379 | 0.705 | 0.713 |  |
| Overweight | Fusiform | 0.008 | 0.021 | 0.368 | 0.713 | 0.713 |  |
| Overweight | Thalamus | 0.065 | 0.018 | 3.712 | 0 | 0.001 | ** |
| Overweight | Pallidum | 0.064 | 0.018 | 3.468 | 0.001 | 0.002 | ** |
| Overweight | Accumbens | 0.038 | 0.019 | 1.948 | 0.051 | 0.139 |  |
| Overweight | Hippocampus | 0.035 | 0.02 | 1.771 | 0.076 | 0.145 |  |
| Overweight | Putamen | 0.037 | 0.021 | 1.787 | 0.074 | 0.145 |  |
| Overweight | Caudate | 0.018 | 0.022 | 0.797 | 0.426 | 0.539 |  |
| Overweight | Amygdala | 0.013 | 0.019 | 0.7 | 0.484 | 0.575 |  |
| Obese | Medial OFC | -0.143 | 0.021 | -6.738 | 0 | 0 | *** |
| Obese | Lateral OFC | -0.192 | 0.021 | -8.99 | 0 | 0 | *** |
| Obese | Temporal pole | -0.155 | 0.023 | -6.811 | 0 | 0 | *** |
| Obese | Superior frontal | -0.185 | 0.023 | -8.201 | 0 | 0 | *** |
| Obese | Rostral mid-frontal | -0.206 | 0.021 | -9.873 | 0 | 0 | *** |
| Obese | Fusiform | -0.09 | 0.023 | -3.931 | 0 | 0 | *** |
| Obese | Middle temporal | -0.177 | 0.023 | -7.679 | 0 | 0 | *** |
| Obese | Inferior temporal | -0.077 | 0.023 | -3.376 | 0.001 | 0.002 | ** |
| Obese | Rostral ACC | -0.043 | 0.023 | -1.871 | 0.061 | 0.102 |  |
| Obese | Lateral occipital | -0.038 | 0.022 | -1.667 | 0.095 | 0.139 |  |
| Obese | Insula | -0.028 | 0.022 | -1.27 | 0.204 | 0.242 |  |
| Obese | Caudal ACC | -0.03 | 0.025 | -1.215 | 0.224 | 0.251 |  |
| Obese | Pallidum | 0.064 | 0.02 | 3.238 | 0.001 | 0.003 | ** |
| Obese | Amygdala | -0.051 | 0.02 | -2.52 | 0.012 | 0.022 | * |
| Obese | Thalamus | 0.035 | 0.019 | 1.847 | 0.065 | 0.102 |  |
| Obese | Accumbens | 0.028 | 0.021 | 1.363 | 0.173 | 0.219 |  |
| Obese | Caudate | -0.033 | 0.024 | -1.369 | 0.171 | 0.219 |  |
| Obese | Hippocampus | -0.022 | 0.021 | -1.074 | 0.283 | 0.298 |  |
| Obese | Putamen | 0.014 | 0.022 | 0.632 | 0.528 | 0.528 |  |
| Underweight | Lateral occipital | -0.139 | 0.048 | -2.886 | 0.004 | 0.015 | * |
| Underweight | Fusiform | -0.103 | 0.049 | -2.096 | 0.036 | 0.093 |  |
| Underweight | Middle temporal | -0.103 | 0.05 | -2.064 | 0.039 | 0.093 |  |
| Underweight | Insula | -0.084 | 0.048 | -1.764 | 0.078 | 0.148 |  |
| Underweight | Rostral ACC | -0.077 | 0.05 | -1.552 | 0.121 | 0.177 |  |
| Underweight | Caudal ACC | -0.085 | 0.054 | -1.585 | 0.113 | 0.177 |  |
| Underweight | Inferior temporal | -0.061 | 0.05 | -1.221 | 0.222 | 0.301 |  |
| Underweight | Rostral mid-frontal | 0.045 | 0.045 | 0.993 | 0.321 | 0.406 |  |
| Underweight | Superior frontal | 0.033 | 0.048 | 0.69 | 0.49 | 0.582 |  |
| Underweight | Lateral OFC | -0.029 | 0.046 | -0.627 | 0.531 | 0.593 |  |
| Underweight | Temporal pole | 0.024 | 0.05 | 0.491 | 0.623 | 0.658 |  |
| Underweight | Medial OFC | -0.012 | 0.046 | -0.259 | 0.795 | 0.795 |  |
| Underweight | Thalamus | -0.172 | 0.04 | -4.293 | 0 | 0 | *** |
| Underweight | Pallidum | -0.166 | 0.042 | -3.958 | 0 | 0.001 | *** |
| Underweight | Hippocampus | -0.16 | 0.044 | -3.606 | 0 | 0.002 | ** |
| Underweight | Putamen | -0.149 | 0.047 | -3.187 | 0.001 | 0.007 | ** |
| Underweight | Caudate | -0.112 | 0.05 | -2.22 | 0.026 | 0.084 |  |
| Underweight | Accumbens | -0.08 | 0.044 | -1.806 | 0.071 | 0.148 |  |
| Underweight | Amygdala | -0.067 | 0.043 | -1.55 | 0.121 | 0.177 |  |
| Note. Adjusted differences between each CDC weight status group and the healthy weight reference group were estimated across all participants from the group model described in eTable 8 (restricted maximum likelihood), in SD units of each outcome; these estimates are mapped in Figure 2. P values are from Wald tests (z = difference/SE); FDR correction was applied within each group across 19 regions, and significance markers are based on unrounded q values. BMI, body mass index; CDC, Centers for Disease Control and Prevention; FDR, false discovery rate; SE, standard error; q, FDR-corrected P value. *q < .05; **q < .01; ***q < .001. | | | | | | | |

| **eTable 10. Likelihood Ratio Tests for Differences in Age-Related Change in Regional Brain Structure Between CDC Weight Status Groups.** | | | | | | |
| --- | --- | --- | --- | --- | --- | --- |
| test | region | LR χ² | df | p | q | sig |
| Group x age | Superior frontal | 49.53 | 3 | 0 | 0 | *** |
| Group x age | Rostral mid-frontal | 42.01 | 3 | 0 | 0 | *** |
| Group x age | Medial OFC | 13 | 3 | 0.005 | 0.02 | * |
| Group x age | Caudal ACC | 10.35 | 3 | 0.016 | 0.05 | * |
| Group x age | Fusiform | 8.14 | 3 | 0.043 | 0.103 |  |
| Group x age | Temporal pole | 5.47 | 3 | 0.141 | 0.243 |  |
| Group x age | Insula | 3.53 | 3 | 0.316 | 0.462 |  |
| Group x age | Lateral OFC | 3.35 | 3 | 0.341 | 0.463 |  |
| Group x age | Lateral occipital | 2.91 | 3 | 0.406 | 0.515 |  |
| Group x age | Rostral ACC | 1.69 | 3 | 0.639 | 0.714 |  |
| Group x age | Middle temporal | 1.19 | 3 | 0.756 | 0.798 |  |
| Group x age | Inferior temporal | 0.03 | 3 | 0.999 | 0.999 |  |
| Group x age | Accumbens | 23.69 | 3 | 0 | 0 | *** |
| Group x age | Hippocampus | 12.71 | 3 | 0.005 | 0.02 | * |
| Group x age | Putamen | 10.03 | 3 | 0.018 | 0.05 | * |
| Group x age | Amygdala | 7.07 | 3 | 0.07 | 0.147 |  |
| Group x age | Thalamus | 6.57 | 3 | 0.087 | 0.165 |  |
| Group x age | Caudate | 4.9 | 3 | 0.179 | 0.284 |  |
| Group x age | Pallidum | 2.35 | 3 | 0.504 | 0.598 |  |
| Note. Group differences in age-related change were tested across all participants by comparing the group model described in eTable 8 with and without weight status group × age interactions (linear age; 3 df) using likelihood ratio tests, with both models fitted by maximum likelihood; age² was included as a common term. Models were adjusted for age, age², sex, pubertal development, socioeconomic status, race and ethnicity, and intracranial volume (subcortical outcomes), with random intercepts for family and participant. LR χ² is the likelihood ratio statistic. FDR correction was applied across 19 regions, and significance markers are based on unrounded q values. BMI, body mass index; CDC, Centers for Disease Control and Prevention; FDR, false discovery rate; LR, likelihood ratio; q, FDR-corrected P value. *q < .05; **q < .01; ***q < .001. | | | | | | |

| **eTable 11. Age Slopes of Regional Brain Structure Within Each CDC Weight Status Group.** | | | | | | | |
| --- | --- | --- | --- | --- | --- | --- | --- |
| group | region | slope per year | SE | z | p | q | sig |
| Healthy weight | Caudal ACC | -0.1062 | 0.0023 | -45.591 | 0 | 0 | *** |
| Healthy weight | Fusiform | -0.1309 | 0.0029 | -45.435 | 0 | 0 | *** |
| Healthy weight | Inferior temporal | -0.1298 | 0.0029 | -45.128 | 0 | 0 | *** |
| Healthy weight | Insula | -0.1063 | 0.0036 | -29.598 | 0 | 0 | *** |
| Healthy weight | Lateral OFC | -0.1669 | 0.0031 | -53.479 | 0 | 0 | *** |
| Healthy weight | Lateral occipital | -0.1292 | 0.0027 | -47.72 | 0 | 0 | *** |
| Healthy weight | Medial OFC | -0.1784 | 0.0032 | -56.203 | 0 | 0 | *** |
| Healthy weight | Middle temporal | -0.1232 | 0.0029 | -43.196 | 0 | 0 | *** |
| Healthy weight | Rostral ACC | -0.1174 | 0.0033 | -35.259 | 0 | 0 | *** |
| Healthy weight | Rostral mid-frontal | -0.1807 | 0.0029 | -61.538 | 0 | 0 | *** |
| Healthy weight | Superior frontal | -0.1493 | 0.0029 | -51.707 | 0 | 0 | *** |
| Healthy weight | Temporal pole | -0.0055 | 0.004 | -1.377 | 0.169 | 0.169 |  |
| Healthy weight | Accumbens | -0.0463 | 0.0027 | -17.123 | 0 | 0 | *** |
| Healthy weight | Amygdala | 0.0157 | 0.0022 | 7.059 | 0 | 0 | *** |
| Healthy weight | Caudate | -0.031 | 0.0012 | -25.118 | 0 | 0 | *** |
| Healthy weight | Hippocampus | 0.0303 | 0.0015 | 20.61 | 0 | 0 | *** |
| Healthy weight | Pallidum | 0.0353 | 0.0028 | 12.694 | 0 | 0 | *** |
| Healthy weight | Putamen | -0.0173 | 0.0016 | -11.094 | 0 | 0 | *** |
| Healthy weight | Thalamus | 0.0396 | 0.002 | 19.388 | 0 | 0 | *** |
| Overweight | Caudal ACC | -0.1055 | 0.003 | -35.279 | 0 | 0 | *** |
| Overweight | Fusiform | -0.1386 | 0.0037 | -37.165 | 0 | 0 | *** |
| Overweight | Inferior temporal | -0.13 | 0.0037 | -34.951 | 0 | 0 | *** |
| Overweight | Insula | -0.1078 | 0.0047 | -22.921 | 0 | 0 | *** |
| Overweight | Lateral OFC | -0.1688 | 0.0041 | -41.562 | 0 | 0 | *** |
| Overweight | Lateral occipital | -0.1257 | 0.0035 | -35.959 | 0 | 0 | *** |
| Overweight | Medial OFC | -0.178 | 0.0041 | -43.012 | 0 | 0 | *** |
| Overweight | Middle temporal | -0.1253 | 0.0037 | -33.98 | 0 | 0 | *** |
| Overweight | Rostral ACC | -0.1215 | 0.0043 | -28.03 | 0 | 0 | *** |
| Overweight | Rostral mid-frontal | -0.1783 | 0.0038 | -46.732 | 0 | 0 | *** |
| Overweight | Superior frontal | -0.1445 | 0.0037 | -38.646 | 0 | 0 | *** |
| Overweight | Temporal pole | -0.0063 | 0.0052 | -1.207 | 0.228 | 0.228 |  |
| Overweight | Accumbens | -0.0426 | 0.0035 | -12.164 | 0 | 0 | *** |
| Overweight | Amygdala | 0.0184 | 0.0029 | 6.447 | 0 | 0 | *** |
| Overweight | Caudate | -0.0295 | 0.0016 | -18.714 | 0 | 0 | *** |
| Overweight | Hippocampus | 0.0274 | 0.0019 | 14.596 | 0 | 0 | *** |
| Overweight | Pallidum | 0.0363 | 0.0036 | 10.063 | 0 | 0 | *** |
| Overweight | Putamen | -0.0213 | 0.002 | -10.732 | 0 | 0 | *** |
| Overweight | Thalamus | 0.0354 | 0.0026 | 13.467 | 0 | 0 | *** |
| Obese | Caudal ACC | -0.0973 | 0.003 | -32.131 | 0 | 0 | *** |
| Obese | Fusiform | -0.1376 | 0.0038 | -36.405 | 0 | 0 | *** |
| Obese | Inferior temporal | -0.1295 | 0.0038 | -34.346 | 0 | 0 | *** |
| Obese | Insula | -0.1086 | 0.0048 | -22.773 | 0 | 0 | *** |
| Obese | Lateral OFC | -0.1625 | 0.0041 | -39.457 | 0 | 0 | *** |
| Obese | Lateral occipital | -0.1269 | 0.0035 | -35.805 | 0 | 0 | *** |
| Obese | Medial OFC | -0.1683 | 0.0042 | -40.093 | 0 | 0 | *** |
| Obese | Middle temporal | -0.122 | 0.0037 | -32.661 | 0 | 0 | *** |
| Obese | Rostral ACC | -0.1176 | 0.0044 | -26.757 | 0 | 0 | *** |
| Obese | Rostral mid-frontal | -0.1576 | 0.0039 | -40.742 | 0 | 0 | *** |
| Obese | Superior frontal | -0.1257 | 0.0038 | -33.166 | 0 | 0 | *** |
| Obese | Temporal pole | -0.015 | 0.0053 | -2.815 | 0.005 | 0.005 | ** |
| Obese | Accumbens | -0.0312 | 0.0036 | -8.778 | 0 | 0 | *** |
| Obese | Amygdala | 0.0224 | 0.0029 | 7.719 | 0 | 0 | *** |
| Obese | Caudate | -0.0284 | 0.0016 | -17.806 | 0 | 0 | *** |
| Obese | Hippocampus | 0.0327 | 0.0019 | 17.201 | 0 | 0 | *** |
| Obese | Pallidum | 0.0306 | 0.0037 | 8.357 | 0 | 0 | *** |
| Obese | Putamen | -0.0152 | 0.002 | -7.522 | 0 | 0 | *** |
| Obese | Thalamus | 0.0395 | 0.0027 | 14.815 | 0 | 0 | *** |
| Underweight | Caudal ACC | -0.1051 | 0.0065 | -16.153 | 0 | 0 | *** |
| Underweight | Fusiform | -0.1394 | 0.0082 | -17.086 | 0 | 0 | *** |
| Underweight | Inferior temporal | -0.1288 | 0.0081 | -15.826 | 0 | 0 | *** |
| Underweight | Insula | -0.1249 | 0.0104 | -12.034 | 0 | 0 | *** |
| Underweight | Lateral OFC | -0.1769 | 0.0089 | -19.821 | 0 | 0 | *** |
| Underweight | Lateral occipital | -0.1195 | 0.0076 | -15.644 | 0 | 0 | *** |
| Underweight | Medial OFC | -0.1986 | 0.0091 | -21.806 | 0 | 0 | *** |
| Underweight | Middle temporal | -0.1175 | 0.0081 | -14.583 | 0 | 0 | *** |
| Underweight | Rostral ACC | -0.1113 | 0.0095 | -11.689 | 0 | 0 | *** |
| Underweight | Rostral mid-frontal | -0.1738 | 0.0084 | -20.766 | 0 | 0 | *** |
| Underweight | Superior frontal | -0.1296 | 0.0082 | -15.838 | 0 | 0 | *** |
| Underweight | Temporal pole | -0.0222 | 0.0116 | -1.913 | 0.056 | 0.056 |  |
| Underweight | Accumbens | -0.0321 | 0.0077 | -4.183 | 0 | 0 | *** |
| Underweight | Amygdala | 0.0135 | 0.0062 | 2.158 | 0.031 | 0.033 | * |
| Underweight | Caudate | -0.0336 | 0.0034 | -9.871 | 0 | 0 | *** |
| Underweight | Hippocampus | 0.0401 | 0.0041 | 9.857 | 0 | 0 | *** |
| Underweight | Pallidum | 0.0369 | 0.0079 | 4.659 | 0 | 0 | *** |
| Underweight | Putamen | -0.012 | 0.0043 | -2.793 | 0.005 | 0.006 | ** |
| Underweight | Thalamus | 0.0488 | 0.0057 | 8.508 | 0 | 0 | *** |
| Note. Age slopes were estimated for each CDC weight status group from the group × age model described in eTable 10 (restricted maximum likelihood) at the sample mean age (12.6 years) and are expressed in SD units of each outcome per year (coefficients per SD of age divided by the age SD, 2.47 years). Negative values indicate thinning or volume decline with age. P values are from Wald tests (z = slope/SE); FDR correction was applied within each group across 19 regions, and significance markers are based on unrounded q values. BMI, body mass index; CDC, Centers for Disease Control and Prevention; FDR, false discovery rate; SE, standard error; q, FDR-corrected P value. *q < .05; **q < .01; ***q < .001. | | | | | | | |

| **eTable 12. Differences in Age Slopes of Regional Brain Structure Between Each CDC Weight Status Group and the Healthy Weight Group.** | | | | | | | | |
| --- | --- | --- | --- | --- | --- | --- | --- | --- |
| group | measure | region | diff per year | SE | t | p | q | sig |
| Overweight | Cortical thickness | Fusiform | -0.0076 | 0.0033 | -2.314 | 0.021 | 0.196 |  |
| Overweight | Cortical thickness | Superior frontal | 0.0049 | 0.0033 | 1.47 | 0.142 | 0.538 |  |
| Overweight | Cortical thickness | Rostral ACC | -0.0041 | 0.0039 | -1.062 | 0.288 | 0.547 |  |
| Overweight | Cortical thickness | Lateral occipital | 0.0035 | 0.0031 | 1.122 | 0.262 | 0.547 |  |
| Overweight | Cortical thickness | Rostral mid-frontal | 0.0024 | 0.0034 | 0.705 | 0.481 | 0.824 |  |
| Overweight | Cortical thickness | Middle temporal | -0.0021 | 0.0033 | -0.642 | 0.521 | 0.824 |  |
| Overweight | Cortical thickness | Lateral OFC | -0.002 | 0.0036 | -0.541 | 0.589 | 0.86 |  |
| Overweight | Cortical thickness | Caudal ACC | 7e-04 | 0.0026 | 0.266 | 0.79 | 0.938 |  |
| Overweight | Cortical thickness | Insula | -0.0015 | 0.0042 | -0.363 | 0.716 | 0.938 |  |
| Overweight | Cortical thickness | Medial OFC | 4e-04 | 0.0037 | 0.105 | 0.916 | 0.943 |  |
| Overweight | Cortical thickness | Temporal pole | -8e-04 | 0.0047 | -0.179 | 0.858 | 0.943 |  |
| Overweight | Cortical thickness | Inferior temporal | -2e-04 | 0.0033 | -0.072 | 0.943 | 0.943 |  |
| Overweight | Subcortical volume | Putamen | -0.0041 | 0.0017 | -2.349 | 0.019 | 0.196 |  |
| Overweight | Subcortical volume | Hippocampus | -0.0029 | 0.0016 | -1.764 | 0.078 | 0.369 |  |
| Overweight | Subcortical volume | Thalamus | -0.0042 | 0.0023 | -1.817 | 0.069 | 0.369 |  |
| Overweight | Subcortical volume | Accumbens | 0.0037 | 0.0031 | 1.197 | 0.231 | 0.547 |  |
| Overweight | Subcortical volume | Amygdala | 0.0028 | 0.0025 | 1.096 | 0.273 | 0.547 |  |
| Overweight | Subcortical volume | Caudate | 0.0016 | 0.0014 | 1.148 | 0.251 | 0.547 |  |
| Overweight | Subcortical volume | Pallidum | 0.0011 | 0.0032 | 0.328 | 0.743 | 0.938 |  |
| Obese | Cortical thickness | Superior frontal | 0.0237 | 0.0035 | 6.767 | 0 | 0 | *** |
| Obese | Cortical thickness | Rostral mid-frontal | 0.0231 | 0.0036 | 6.448 | 0 | 0 | *** |
| Obese | Cortical thickness | Caudal ACC | 0.0089 | 0.0028 | 3.194 | 0.001 | 0.007 | ** |
| Obese | Cortical thickness | Medial OFC | 0.0102 | 0.0039 | 2.607 | 0.009 | 0.035 | * |
| Obese | Cortical thickness | Temporal pole | -0.0095 | 0.005 | -1.906 | 0.057 | 0.135 |  |
| Obese | Cortical thickness | Fusiform | -0.0066 | 0.0035 | -1.903 | 0.057 | 0.135 |  |
| Obese | Cortical thickness | Lateral OFC | 0.0044 | 0.0038 | 1.143 | 0.253 | 0.37 |  |
| Obese | Cortical thickness | Lateral occipital | 0.0023 | 0.0033 | 0.708 | 0.479 | 0.65 |  |
| Obese | Cortical thickness | Insula | -0.0024 | 0.0044 | -0.532 | 0.595 | 0.753 |  |
| Obese | Cortical thickness | Middle temporal | 0.0011 | 0.0034 | 0.333 | 0.739 | 0.878 |  |
| Obese | Cortical thickness | Rostral ACC | -2e-04 | 0.0041 | -0.044 | 0.965 | 0.965 |  |
| Obese | Cortical thickness | Inferior temporal | 3e-04 | 0.0035 | 0.084 | 0.933 | 0.965 |  |
| Obese | Subcortical volume | Accumbens | 0.0151 | 0.0033 | 4.612 | 0 | 0 | *** |
| Obese | Subcortical volume | Amygdala | 0.0067 | 0.0027 | 2.513 | 0.012 | 0.038 | * |
| Obese | Subcortical volume | Caudate | 0.0026 | 0.0015 | 1.807 | 0.071 | 0.149 |  |
| Obese | Subcortical volume | Hippocampus | 0.0024 | 0.0017 | 1.403 | 0.161 | 0.291 |  |
| Obese | Subcortical volume | Pallidum | -0.0047 | 0.0034 | -1.377 | 0.168 | 0.291 |  |
| Obese | Subcortical volume | Putamen | 0.0021 | 0.0018 | 1.148 | 0.251 | 0.37 |  |
| Obese | Subcortical volume | Thalamus | -1e-04 | 0.0025 | -0.048 | 0.962 | 0.965 |  |
| Underweight | Cortical thickness | Superior frontal | 0.0198 | 0.008 | 2.479 | 0.013 | 0.125 |  |
| Underweight | Cortical thickness | Medial OFC | -0.0201 | 0.0089 | -2.263 | 0.024 | 0.15 |  |
| Underweight | Cortical thickness | Insula | -0.0186 | 0.0102 | -1.834 | 0.067 | 0.253 |  |
| Underweight | Cortical thickness | Temporal pole | -0.0167 | 0.0114 | -1.472 | 0.141 | 0.383 |  |
| Underweight | Cortical thickness | Lateral occipital | 0.0097 | 0.0074 | 1.302 | 0.193 | 0.448 |  |
| Underweight | Cortical thickness | Lateral OFC | -0.01 | 0.0087 | -1.151 | 0.25 | 0.474 |  |
| Underweight | Cortical thickness | Fusiform | -0.0084 | 0.008 | -1.058 | 0.29 | 0.501 |  |
| Underweight | Cortical thickness | Rostral mid-frontal | 0.0068 | 0.0082 | 0.837 | 0.402 | 0.632 |  |
| Underweight | Cortical thickness | Middle temporal | 0.0056 | 0.0079 | 0.718 | 0.473 | 0.642 |  |
| Underweight | Cortical thickness | Rostral ACC | 0.0061 | 0.0093 | 0.651 | 0.515 | 0.653 |  |
| Underweight | Cortical thickness | Caudal ACC | 0.0011 | 0.0063 | 0.176 | 0.86 | 0.899 |  |
| Underweight | Cortical thickness | Inferior temporal | 0.001 | 0.0079 | 0.126 | 0.899 | 0.899 |  |
| Underweight | Subcortical volume | Hippocampus | 0.0098 | 0.004 | 2.486 | 0.013 | 0.125 |  |
| Underweight | Subcortical volume | Accumbens | 0.0142 | 0.0075 | 1.904 | 0.057 | 0.253 |  |
| Underweight | Subcortical volume | Thalamus | 0.0092 | 0.0056 | 1.64 | 0.101 | 0.32 |  |
| Underweight | Subcortical volume | Putamen | 0.0052 | 0.0042 | 1.248 | 0.212 | 0.448 |  |
| Underweight | Subcortical volume | Caudate | -0.0026 | 0.0033 | -0.785 | 0.432 | 0.632 |  |
| Underweight | Subcortical volume | Amygdala | -0.0022 | 0.0061 | -0.366 | 0.715 | 0.849 |  |
| Underweight | Subcortical volume | Pallidum | 0.0016 | 0.0077 | 0.211 | 0.833 | 0.899 |  |
| Note. Differences in age slope between each CDC weight status group and the healthy weight reference group are the group × age interaction coefficients from the model described in eTable 10 (restricted maximum likelihood), expressed in SD units of each outcome per year. Positive values indicate slower thinning or volume decline (or faster increase) than in the healthy weight group. P values are from Wald tests (z = difference/SE); FDR correction was applied within each group across 19 regions, and significance markers are based on unrounded q values. Contrasts are interpreted only for regions with a significant group × age likelihood ratio test (eTable 10); the obesity contrast for the amygdala is therefore not interpreted. BMI, body mass index; CDC, Centers for Disease Control and Prevention; FDR, false discovery rate; SE, standard error; q, FDR-corrected P value. *q < .05; **q < .01; ***q < .001. | | | | | | | | |

| **eTable 13. Differences in Regional Brain Structure Between Each CDC Weight Status Group and the Healthy Weight Group at Ages 10, 13, and 16 Years.** | | | | | | | | |
| --- | --- | --- | --- | --- | --- | --- | --- | --- |
| group | region | age | diff | SE | z | p | q | sig |
| Overweight | Caudal ACC | 10 | -0.011 | 0.024 | -0.452 | 0.651 | 0.728 |  |
| Overweight | Caudal ACC | 13 | -0.009 | 0.023 | -0.377 | 0.706 | 0.745 |  |
| Overweight | Caudal ACC | 16 | -0.007 | 0.025 | -0.268 | 0.789 | 0.789 |  |
| Overweight | Fusiform | 10 | 0.027 | 0.023 | 1.16 | 0.246 | 0.334 |  |
| Overweight | Fusiform | 13 | 0.004 | 0.022 | 0.171 | 0.864 | 0.864 |  |
| Overweight | Fusiform | 16 | -0.019 | 0.025 | -0.782 | 0.434 | 0.51 |  |
| Overweight | Inferior temporal | 10 | -0.017 | 0.023 | -0.735 | 0.462 | 0.585 |  |
| Overweight | Inferior temporal | 13 | -0.018 | 0.022 | -0.813 | 0.416 | 0.526 |  |
| Overweight | Inferior temporal | 16 | -0.018 | 0.025 | -0.745 | 0.456 | 0.51 |  |
| Overweight | Insula | 10 | 0.038 | 0.023 | 1.653 | 0.098 | 0.208 |  |
| Overweight | Insula | 13 | 0.034 | 0.021 | 1.616 | 0.106 | 0.174 |  |
| Overweight | Insula | 16 | 0.029 | 0.026 | 1.139 | 0.255 | 0.389 |  |
| Overweight | Lateral OFC | 10 | -0.033 | 0.022 | -1.498 | 0.134 | 0.232 |  |
| Overweight | Lateral OFC | 13 | -0.039 | 0.02 | -1.917 | 0.055 | 0.131 |  |
| Overweight | Lateral OFC | 16 | -0.045 | 0.024 | -1.868 | 0.062 | 0.147 |  |
| Overweight | Lateral occipital | 10 | 0.004 | 0.022 | 0.16 | 0.873 | 0.873 |  |
| Overweight | Lateral occipital | 13 | 0.014 | 0.021 | 0.659 | 0.51 | 0.57 |  |
| Overweight | Lateral occipital | 16 | 0.024 | 0.024 | 1.021 | 0.307 | 0.389 |  |
| Overweight | Medial OFC | 10 | -0.033 | 0.022 | -1.518 | 0.129 | 0.232 |  |
| Overweight | Medial OFC | 13 | -0.032 | 0.02 | -1.598 | 0.11 | 0.174 |  |
| Overweight | Medial OFC | 16 | -0.031 | 0.024 | -1.293 | 0.196 | 0.389 |  |
| Overweight | Middle temporal | 10 | -0.054 | 0.023 | -2.338 | 0.019 | 0.061 |  |
| Overweight | Middle temporal | 13 | -0.06 | 0.022 | -2.765 | 0.006 | 0.018 | * |
| Overweight | Middle temporal | 16 | -0.067 | 0.025 | -2.7 | 0.007 | 0.033 | * |
| Overweight | Rostral ACC | 10 | 0.033 | 0.024 | 1.414 | 0.157 | 0.249 |  |
| Overweight | Rostral ACC | 13 | 0.021 | 0.022 | 0.968 | 0.333 | 0.487 |  |
| Overweight | Rostral ACC | 16 | 0.009 | 0.026 | 0.341 | 0.733 | 0.773 |  |
| Overweight | Rostral mid-frontal | 10 | -0.114 | 0.021 | -5.382 | 0 | 0 | *** |
| Overweight | Rostral mid-frontal | 13 | -0.107 | 0.02 | -5.448 | 0 | 0 | *** |
| Overweight | Rostral mid-frontal | 16 | -0.1 | 0.023 | -4.347 | 0 | 0 | *** |
| Overweight | Superior frontal | 10 | -0.126 | 0.023 | -5.589 | 0 | 0 | *** |
| Overweight | Superior frontal | 13 | -0.112 | 0.021 | -5.266 | 0 | 0 | *** |
| Overweight | Superior frontal | 16 | -0.097 | 0.024 | -4.011 | 0 | 0.001 | *** |
| Overweight | Temporal pole | 10 | -0.057 | 0.024 | -2.338 | 0.019 | 0.061 |  |
| Overweight | Temporal pole | 13 | -0.06 | 0.022 | -2.76 | 0.006 | 0.018 | * |
| Overweight | Temporal pole | 16 | -0.062 | 0.027 | -2.287 | 0.022 | 0.065 |  |
| Overweight | Accumbens | 10 | 0.028 | 0.021 | 1.35 | 0.177 | 0.259 |  |
| Overweight | Accumbens | 13 | 0.039 | 0.019 | 2.015 | 0.044 | 0.119 |  |
| Overweight | Accumbens | 16 | 0.05 | 0.022 | 2.255 | 0.024 | 0.065 |  |
| Overweight | Amygdala | 10 | 0.006 | 0.02 | 0.317 | 0.751 | 0.793 |  |
| Overweight | Amygdala | 13 | 0.015 | 0.019 | 0.767 | 0.443 | 0.526 |  |
| Overweight | Amygdala | 16 | 0.023 | 0.021 | 1.089 | 0.276 | 0.389 |  |
| Overweight | Caudate | 10 | 0.014 | 0.023 | 0.617 | 0.537 | 0.638 |  |
| Overweight | Caudate | 13 | 0.019 | 0.022 | 0.834 | 0.404 | 0.526 |  |
| Overweight | Caudate | 16 | 0.023 | 0.023 | 1.021 | 0.307 | 0.389 |  |
| Overweight | Hippocampus | 10 | 0.041 | 0.02 | 2.074 | 0.038 | 0.09 |  |
| Overweight | Hippocampus | 13 | 0.033 | 0.02 | 1.67 | 0.095 | 0.174 |  |
| Overweight | Hippocampus | 16 | 0.024 | 0.02 | 1.177 | 0.239 | 0.389 |  |
| Overweight | Pallidum | 10 | 0.061 | 0.02 | 3.072 | 0.002 | 0.01 | * |
| Overweight | Pallidum | 13 | 0.064 | 0.018 | 3.495 | 0 | 0.002 | ** |
| Overweight | Pallidum | 16 | 0.068 | 0.022 | 3.129 | 0.002 | 0.011 | * |
| Overweight | Putamen | 10 | 0.047 | 0.021 | 2.204 | 0.027 | 0.075 |  |
| Overweight | Putamen | 13 | 0.034 | 0.021 | 1.657 | 0.098 | 0.174 |  |
| Overweight | Putamen | 16 | 0.022 | 0.022 | 1.025 | 0.306 | 0.389 |  |
| Overweight | Thalamus | 10 | 0.075 | 0.018 | 4.086 | 0 | 0 | *** |
| Overweight | Thalamus | 13 | 0.063 | 0.018 | 3.559 | 0 | 0.002 | ** |
| Overweight | Thalamus | 16 | 0.05 | 0.019 | 2.581 | 0.01 | 0.037 | * |
| Obese | Caudal ACC | 10 | -0.051 | 0.026 | -1.985 | 0.047 | 0.081 |  |
| Obese | Caudal ACC | 13 | -0.024 | 0.025 | -0.973 | 0.33 | 0.349 |  |
| Obese | Caudal ACC | 16 | 0.002 | 0.027 | 0.093 | 0.926 | 0.926 |  |
| Obese | Fusiform | 10 | -0.074 | 0.024 | -3.04 | 0.002 | 0.004 | ** |
| Obese | Fusiform | 13 | -0.094 | 0.023 | -4.083 | 0 | 0 | *** |
| Obese | Fusiform | 16 | -0.114 | 0.026 | -4.349 | 0 | 0 | *** |
| Obese | Inferior temporal | 10 | -0.078 | 0.024 | -3.207 | 0.001 | 0.003 | ** |
| Obese | Inferior temporal | 13 | -0.077 | 0.023 | -3.352 | 0.001 | 0.002 | ** |
| Obese | Inferior temporal | 16 | -0.076 | 0.026 | -2.915 | 0.004 | 0.008 | ** |
| Obese | Insula | 10 | -0.022 | 0.025 | -0.907 | 0.364 | 0.407 |  |
| Obese | Insula | 13 | -0.029 | 0.022 | -1.32 | 0.187 | 0.232 |  |
| Obese | Insula | 16 | -0.036 | 0.027 | -1.337 | 0.181 | 0.265 |  |
| Obese | Lateral OFC | 10 | -0.203 | 0.023 | -8.713 | 0 | 0 | *** |
| Obese | Lateral OFC | 13 | -0.19 | 0.022 | -8.803 | 0 | 0 | *** |
| Obese | Lateral OFC | 16 | -0.176 | 0.025 | -6.927 | 0 | 0 | *** |
| Obese | Lateral occipital | 10 | -0.043 | 0.024 | -1.814 | 0.07 | 0.108 |  |
| Obese | Lateral occipital | 13 | -0.036 | 0.023 | -1.602 | 0.109 | 0.148 |  |
| Obese | Lateral occipital | 16 | -0.029 | 0.025 | -1.15 | 0.25 | 0.317 |  |
| Obese | Medial OFC | 10 | -0.167 | 0.023 | -7.211 | 0 | 0 | *** |
| Obese | Medial OFC | 13 | -0.136 | 0.021 | -6.397 | 0 | 0 | *** |
| Obese | Medial OFC | 16 | -0.106 | 0.025 | -4.165 | 0 | 0 | *** |
| Obese | Middle temporal | 10 | -0.18 | 0.024 | -7.35 | 0 | 0 | *** |
| Obese | Middle temporal | 13 | -0.176 | 0.023 | -7.609 | 0 | 0 | *** |
| Obese | Middle temporal | 16 | -0.173 | 0.026 | -6.586 | 0 | 0 | *** |
| Obese | Rostral ACC | 10 | -0.042 | 0.025 | -1.7 | 0.089 | 0.121 |  |
| Obese | Rostral ACC | 13 | -0.043 | 0.023 | -1.858 | 0.063 | 0.102 |  |
| Obese | Rostral ACC | 16 | -0.043 | 0.027 | -1.592 | 0.111 | 0.176 |  |
| Obese | Rostral mid-frontal | 10 | -0.261 | 0.023 | -11.576 | 0 | 0 | *** |
| Obese | Rostral mid-frontal | 13 | -0.192 | 0.021 | -9.132 | 0 | 0 | *** |
| Obese | Rostral mid-frontal | 16 | -0.122 | 0.025 | -4.982 | 0 | 0 | *** |
| Obese | Superior frontal | 10 | -0.241 | 0.024 | -10.036 | 0 | 0 | *** |
| Obese | Superior frontal | 13 | -0.17 | 0.023 | -7.503 | 0 | 0 | *** |
| Obese | Superior frontal | 16 | -0.099 | 0.026 | -3.82 | 0 | 0 | *** |
| Obese | Temporal pole | 10 | -0.132 | 0.026 | -5.132 | 0 | 0 | *** |
| Obese | Temporal pole | 13 | -0.161 | 0.023 | -6.996 | 0 | 0 | *** |
| Obese | Temporal pole | 16 | -0.189 | 0.029 | -6.538 | 0 | 0 | *** |
| Obese | Accumbens | 10 | -0.008 | 0.022 | -0.353 | 0.724 | 0.724 |  |
| Obese | Accumbens | 13 | 0.038 | 0.021 | 1.812 | 0.07 | 0.102 |  |
| Obese | Accumbens | 16 | 0.083 | 0.024 | 3.482 | 0 | 0.001 | ** |
| Obese | Amygdala | 10 | -0.067 | 0.021 | -3.155 | 0.002 | 0.003 | ** |
| Obese | Amygdala | 13 | -0.047 | 0.02 | -2.3 | 0.021 | 0.041 | * |
| Obese | Amygdala | 16 | -0.027 | 0.022 | -1.184 | 0.237 | 0.317 |  |
| Obese | Caudate | 10 | -0.039 | 0.024 | -1.612 | 0.107 | 0.136 |  |
| Obese | Caudate | 13 | -0.031 | 0.024 | -1.295 | 0.195 | 0.232 |  |
| Obese | Caudate | 16 | -0.023 | 0.024 | -0.942 | 0.346 | 0.387 |  |
| Obese | Hippocampus | 10 | -0.028 | 0.021 | -1.315 | 0.189 | 0.224 |  |
| Obese | Hippocampus | 13 | -0.021 | 0.021 | -0.986 | 0.324 | 0.349 |  |
| Obese | Hippocampus | 16 | -0.013 | 0.022 | -0.609 | 0.543 | 0.573 |  |
| Obese | Pallidum | 10 | 0.075 | 0.021 | 3.514 | 0 | 0.001 | ** |
| Obese | Pallidum | 13 | 0.061 | 0.02 | 3.069 | 0.002 | 0.005 | ** |
| Obese | Pallidum | 16 | 0.047 | 0.023 | 2.012 | 0.044 | 0.084 |  |
| Obese | Putamen | 10 | 0.009 | 0.023 | 0.412 | 0.68 | 0.718 |  |
| Obese | Putamen | 13 | 0.016 | 0.022 | 0.705 | 0.481 | 0.481 |  |
| Obese | Putamen | 16 | 0.022 | 0.023 | 0.95 | 0.342 | 0.387 |  |
| Obese | Thalamus | 10 | 0.035 | 0.02 | 1.789 | 0.074 | 0.108 |  |
| Obese | Thalamus | 13 | 0.035 | 0.019 | 1.846 | 0.065 | 0.102 |  |
| Obese | Thalamus | 16 | 0.034 | 0.021 | 1.656 | 0.098 | 0.169 |  |
| Underweight | Caudal ACC | 10 | -0.087 | 0.055 | -1.576 | 0.115 | 0.199 |  |
| Underweight | Caudal ACC | 13 | -0.084 | 0.054 | -1.557 | 0.119 | 0.199 |  |
| Underweight | Caudal ACC | 16 | -0.081 | 0.059 | -1.369 | 0.171 | 0.25 |  |
| Underweight | Fusiform | 10 | -0.085 | 0.052 | -1.618 | 0.106 | 0.199 |  |
| Underweight | Fusiform | 13 | -0.11 | 0.05 | -2.212 | 0.027 | 0.073 |  |
| Underweight | Fusiform | 16 | -0.135 | 0.058 | -2.34 | 0.019 | 0.052 |  |
| Underweight | Inferior temporal | 10 | -0.063 | 0.053 | -1.193 | 0.233 | 0.34 |  |
| Underweight | Inferior temporal | 13 | -0.06 | 0.05 | -1.195 | 0.232 | 0.315 |  |
| Underweight | Inferior temporal | 16 | -0.057 | 0.058 | -0.978 | 0.328 | 0.389 |  |
| Underweight | Insula | 10 | -0.042 | 0.053 | -0.79 | 0.43 | 0.544 |  |
| Underweight | Insula | 13 | -0.098 | 0.048 | -2.023 | 0.043 | 0.102 |  |
| Underweight | Insula | 16 | -0.154 | 0.061 | -2.522 | 0.012 | 0.044 | * |
| Underweight | Lateral OFC | 10 | -0.006 | 0.05 | -0.127 | 0.899 | 0.899 |  |
| Underweight | Lateral OFC | 13 | -0.036 | 0.047 | -0.78 | 0.435 | 0.486 |  |
| Underweight | Lateral OFC | 16 | -0.067 | 0.057 | -1.175 | 0.24 | 0.304 |  |
| Underweight | Lateral occipital | 10 | -0.161 | 0.051 | -3.152 | 0.002 | 0.006 | ** |
| Underweight | Lateral occipital | 13 | -0.132 | 0.049 | -2.707 | 0.007 | 0.026 | * |
| Underweight | Lateral occipital | 16 | -0.103 | 0.056 | -1.834 | 0.067 | 0.141 |  |
| Underweight | Medial OFC | 10 | 0.034 | 0.05 | 0.668 | 0.504 | 0.599 |  |
| Underweight | Medial OFC | 13 | -0.027 | 0.047 | -0.578 | 0.563 | 0.595 |  |
| Underweight | Medial OFC | 16 | -0.087 | 0.057 | -1.535 | 0.125 | 0.216 |  |
| Underweight | Middle temporal | 10 | -0.115 | 0.053 | -2.184 | 0.029 | 0.079 |  |
| Underweight | Middle temporal | 13 | -0.098 | 0.05 | -1.957 | 0.05 | 0.106 |  |
| Underweight | Middle temporal | 16 | -0.081 | 0.058 | -1.401 | 0.161 | 0.25 |  |
| Underweight | Rostral ACC | 10 | -0.091 | 0.054 | -1.681 | 0.093 | 0.196 |  |
| Underweight | Rostral ACC | 13 | -0.073 | 0.05 | -1.444 | 0.149 | 0.218 |  |
| Underweight | Rostral ACC | 16 | -0.055 | 0.061 | -0.896 | 0.37 | 0.414 |  |
| Underweight | Rostral mid-frontal | 10 | 0.03 | 0.049 | 0.613 | 0.54 | 0.603 |  |
| Underweight | Rostral mid-frontal | 13 | 0.05 | 0.045 | 1.108 | 0.268 | 0.339 |  |
| Underweight | Rostral mid-frontal | 16 | 0.071 | 0.054 | 1.3 | 0.194 | 0.263 |  |
| Underweight | Superior frontal | 10 | -0.01 | 0.051 | -0.197 | 0.844 | 0.891 |  |
| Underweight | Superior frontal | 13 | 0.049 | 0.049 | 1.009 | 0.313 | 0.371 |  |
| Underweight | Superior frontal | 16 | 0.109 | 0.057 | 1.905 | 0.057 | 0.135 |  |
| Underweight | Temporal pole | 10 | 0.063 | 0.056 | 1.116 | 0.264 | 0.359 |  |
| Underweight | Temporal pole | 13 | 0.012 | 0.05 | 0.247 | 0.805 | 0.805 |  |
| Underweight | Temporal pole | 16 | -0.038 | 0.065 | -0.58 | 0.562 | 0.593 |  |
| Underweight | Accumbens | 10 | -0.111 | 0.047 | -2.348 | 0.019 | 0.06 |  |
| Underweight | Accumbens | 13 | -0.068 | 0.045 | -1.531 | 0.126 | 0.199 |  |
| Underweight | Accumbens | 16 | -0.026 | 0.053 | -0.49 | 0.624 | 0.624 |  |
| Underweight | Amygdala | 10 | -0.062 | 0.045 | -1.368 | 0.171 | 0.271 |  |
| Underweight | Amygdala | 13 | -0.068 | 0.043 | -1.576 | 0.115 | 0.199 |  |
| Underweight | Amygdala | 16 | -0.075 | 0.049 | -1.534 | 0.125 | 0.216 |  |
| Underweight | Caudate | 10 | -0.106 | 0.051 | -2.088 | 0.037 | 0.087 |  |
| Underweight | Caudate | 13 | -0.114 | 0.05 | -2.259 | 0.024 | 0.073 |  |
| Underweight | Caudate | 16 | -0.122 | 0.052 | -2.343 | 0.019 | 0.052 |  |
| Underweight | Hippocampus | 10 | -0.181 | 0.045 | -4.006 | 0 | 0.001 | *** |
| Underweight | Hippocampus | 13 | -0.151 | 0.044 | -3.404 | 0.001 | 0.004 | ** |
| Underweight | Hippocampus | 16 | -0.122 | 0.047 | -2.598 | 0.009 | 0.044 | * |
| Underweight | Pallidum | 10 | -0.17 | 0.045 | -3.743 | 0 | 0.001 | ** |
| Underweight | Pallidum | 13 | -0.165 | 0.042 | -3.895 | 0 | 0.001 | *** |
| Underweight | Pallidum | 16 | -0.16 | 0.051 | -3.14 | 0.002 | 0.024 | * |
| Underweight | Putamen | 10 | -0.16 | 0.048 | -3.361 | 0.001 | 0.004 | ** |
| Underweight | Putamen | 13 | -0.145 | 0.047 | -3.079 | 0.002 | 0.01 | ** |
| Underweight | Putamen | 16 | -0.129 | 0.05 | -2.604 | 0.009 | 0.044 | * |
| Underweight | Thalamus | 10 | -0.192 | 0.042 | -4.583 | 0 | 0 | *** |
| Underweight | Thalamus | 13 | -0.164 | 0.04 | -4.081 | 0 | 0.001 | *** |
| Underweight | Thalamus | 16 | -0.137 | 0.045 | -3.021 | 0.003 | 0.024 | * |
| Note. Differences between each CDC weight status group and the healthy weight reference group at ages 10, 13, and 16 years were estimated from the group × age model described in eTable 10 (restricted maximum likelihood) as linear combinations of the group and group × age coefficients, in SD units of each outcome. P values are from Wald tests (z = difference/SE); FDR correction was applied within each group and age across 19 regions, and significance markers are based on unrounded q values. BMI, body mass index; CDC, Centers for Disease Control and Prevention; FDR, false discovery rate; SE, standard error; q, FDR-corrected P value. *q < .05; **q < .01; ***q < .001. | | | | | | | | |

| **eTable 14. Likelihood Ratio Tests for Weight Status Group × Sex Interactions on Regional Brain Structure.** | | | | | | |
| --- | --- | --- | --- | --- | --- | --- |
| test | region | LR χ² | df | p | q | sig |
| Group x sex | Rostral ACC | 6 | 3 | 0.112 | 0.356 |  |
| Group x sex | Superior frontal | 5.76 | 3 | 0.124 | 0.356 |  |
| Group x sex | Rostral mid-frontal | 5.63 | 3 | 0.131 | 0.356 |  |
| Group x sex | Insula | 5.19 | 3 | 0.159 | 0.377 |  |
| Group x sex | Inferior temporal | 4.38 | 3 | 0.223 | 0.424 |  |
| Group x sex | Middle temporal | 4.5 | 3 | 0.212 | 0.424 |  |
| Group x sex | Temporal pole | 3.46 | 3 | 0.326 | 0.564 |  |
| Group x sex | Medial OFC | 2.8 | 3 | 0.423 | 0.574 |  |
| Group x sex | Fusiform | 2.36 | 3 | 0.5 | 0.634 |  |
| Group x sex | Lateral OFC | 1.32 | 3 | 0.725 | 0.818 |  |
| Group x sex | Caudal ACC | 1.05 | 3 | 0.79 | 0.833 |  |
| Group x sex | Lateral occipital | 0.65 | 3 | 0.884 | 0.884 |  |
| Group x sex | Accumbens | 15.19 | 3 | 0.002 | 0.032 | * |
| Group x sex | Hippocampus | 10.3 | 3 | 0.016 | 0.154 |  |
| Group x sex | Thalamus | 8.63 | 3 | 0.035 | 0.219 |  |
| Group x sex | Putamen | 6.38 | 3 | 0.094 | 0.356 |  |
| Group x sex | Caudate | 2.98 | 3 | 0.395 | 0.574 |  |
| Group x sex | Pallidum | 2.89 | 3 | 0.408 | 0.574 |  |
| Group x sex | Amygdala | 1.29 | 3 | 0.732 | 0.818 |  |
| Note. Whether differences between CDC weight status groups differed between male and female participants was tested across all participants by comparing the group model described in eTable 8 with and without weight status group × sex interactions (3 df) using likelihood ratio tests, with both models fitted by maximum likelihood. Models were adjusted for age, age², sex, pubertal development, socioeconomic status, race and ethnicity, and intracranial volume (subcortical outcomes), with random intercepts for family and participant. LR χ² is the likelihood ratio statistic. FDR correction was applied across 19 regions, and significance markers are based on unrounded q values. BMI, body mass index; CDC, Centers for Disease Control and Prevention; FDR, false discovery rate; LR, likelihood ratio; q, FDR-corrected P value. *q < .05; **q < .01; ***q < .001. | | | | | | |

| **eTable 15. Weight Status Group × Sex Interaction Terms for Regional Brain Structure.** | | | | | | | |
| --- | --- | --- | --- | --- | --- | --- | --- |
| group | region | β | SE | z | p | q | sig |
| Overweight | Temporal pole | 0.07 | 0.042 | 1.648 | 0.099 | 0.433 |  |
| Overweight | Superior frontal | 0.079 | 0.042 | 1.888 | 0.059 | 0.433 |  |
| Overweight | Insula | 0.072 | 0.041 | 1.767 | 0.077 | 0.433 |  |
| Overweight | Inferior temporal | 0.082 | 0.043 | 1.921 | 0.055 | 0.433 |  |
| Overweight | Middle temporal | 0.068 | 0.043 | 1.581 | 0.114 | 0.433 |  |
| Overweight | Lateral OFC | 0.035 | 0.04 | 0.889 | 0.374 | 0.71 |  |
| Overweight | Rostral ACC | 0.037 | 0.043 | 0.858 | 0.391 | 0.71 |  |
| Overweight | Rostral mid-frontal | 0.037 | 0.039 | 0.956 | 0.339 | 0.71 |  |
| Overweight | Medial OFC | 0.028 | 0.039 | 0.705 | 0.481 | 0.762 |  |
| Overweight | Caudal ACC | -0.02 | 0.046 | -0.434 | 0.664 | 0.864 |  |
| Overweight | Fusiform | 0.01 | 0.042 | 0.229 | 0.819 | 0.864 |  |
| Overweight | Lateral occipital | 0.014 | 0.042 | 0.324 | 0.746 | 0.864 |  |
| Overweight | Accumbens | -0.031 | 0.038 | -0.822 | 0.411 | 0.71 |  |
| Overweight | Amygdala | 0.037 | 0.037 | 0.996 | 0.319 | 0.71 |  |
| Overweight | Putamen | -0.048 | 0.041 | -1.165 | 0.244 | 0.71 |  |
| Overweight | Caudate | 0.021 | 0.044 | 0.476 | 0.634 | 0.864 |  |
| Overweight | Hippocampus | 0.01 | 0.039 | 0.265 | 0.791 | 0.864 |  |
| Overweight | Pallidum | 0.012 | 0.036 | 0.338 | 0.736 | 0.864 |  |
| Overweight | Thalamus | 0 | 0.035 | -0.012 | 0.991 | 0.991 |  |
| Obese | Rostral mid-frontal | 0.093 | 0.04 | 2.322 | 0.02 | 0.237 |  |
| Obese | Rostral ACC | 0.075 | 0.044 | 1.694 | 0.09 | 0.428 |  |
| Obese | Superior frontal | 0.078 | 0.043 | 1.812 | 0.07 | 0.428 |  |
| Obese | Fusiform | -0.058 | 0.044 | -1.333 | 0.183 | 0.496 |  |
| Obese | Medial OFC | 0.05 | 0.041 | 1.231 | 0.218 | 0.519 |  |
| Obese | Lateral OFC | -0.019 | 0.041 | -0.462 | 0.644 | 0.812 |  |
| Obese | Temporal pole | 0.015 | 0.044 | 0.35 | 0.726 | 0.812 |  |
| Obese | Insula | 0.021 | 0.042 | 0.489 | 0.625 | 0.812 |  |
| Obese | Inferior temporal | -0.016 | 0.044 | -0.363 | 0.717 | 0.812 |  |
| Obese | Middle temporal | 0.034 | 0.044 | 0.773 | 0.44 | 0.812 |  |
| Obese | Lateral occipital | -0.016 | 0.043 | -0.376 | 0.707 | 0.812 |  |
| Obese | Caudal ACC | -0.003 | 0.048 | -0.073 | 0.942 | 0.942 |  |
| Obese | Thalamus | -0.08 | 0.036 | -2.242 | 0.025 | 0.237 |  |
| Obese | Hippocampus | -0.063 | 0.04 | -1.571 | 0.116 | 0.441 |  |
| Obese | Putamen | -0.06 | 0.042 | -1.433 | 0.152 | 0.481 |  |
| Obese | Accumbens | -0.036 | 0.039 | -0.911 | 0.362 | 0.765 |  |
| Obese | Caudate | -0.027 | 0.045 | -0.604 | 0.546 | 0.812 |  |
| Obese | Pallidum | -0.014 | 0.037 | -0.369 | 0.712 | 0.812 |  |
| Obese | Amygdala | -0.005 | 0.039 | -0.119 | 0.905 | 0.942 |  |
| Underweight | Rostral ACC | -0.152 | 0.101 | -1.512 | 0.131 | 0.297 |  |
| Underweight | Insula | 0.152 | 0.097 | 1.579 | 0.114 | 0.297 |  |
| Underweight | Middle temporal | 0.151 | 0.1 | 1.5 | 0.134 | 0.297 |  |
| Underweight | Temporal pole | 0.1 | 0.101 | 0.995 | 0.32 | 0.573 |  |
| Underweight | Caudal ACC | -0.105 | 0.108 | -0.971 | 0.332 | 0.573 |  |
| Underweight | Medial OFC | -0.083 | 0.093 | -0.891 | 0.373 | 0.59 |  |
| Underweight | Superior frontal | 0.059 | 0.098 | 0.601 | 0.548 | 0.749 |  |
| Underweight | Fusiform | 0.048 | 0.099 | 0.481 | 0.631 | 0.749 |  |
| Underweight | Lateral occipital | 0.056 | 0.097 | 0.576 | 0.564 | 0.749 |  |
| Underweight | Lateral OFC | 0.03 | 0.093 | 0.318 | 0.75 | 0.753 |  |
| Underweight | Rostral mid-frontal | 0.029 | 0.091 | 0.315 | 0.753 | 0.753 |  |
| Underweight | Inferior temporal | 0.037 | 0.1 | 0.374 | 0.708 | 0.753 |  |
| Underweight | Accumbens | 0.321 | 0.089 | 3.592 | 0 | 0.006 | ** |
| Underweight | Hippocampus | 0.234 | 0.089 | 2.623 | 0.009 | 0.083 |  |
| Underweight | Caudate | 0.149 | 0.101 | 1.474 | 0.14 | 0.297 |  |
| Underweight | Pallidum | 0.135 | 0.085 | 1.593 | 0.111 | 0.297 |  |
| Underweight | Putamen | 0.16 | 0.094 | 1.7 | 0.089 | 0.297 |  |
| Underweight | Thalamus | 0.134 | 0.081 | 1.67 | 0.095 | 0.297 |  |
| Underweight | Amygdala | 0.042 | 0.087 | 0.481 | 0.63 | 0.749 |  |
| Note. Interaction coefficients from the group × sex model described in eTable 14 (restricted maximum likelihood; male participants as reference) give the difference between female and male participants in each group’s difference from the healthy weight group, in SD units of each outcome. Positive values indicate a less negative or more positive group difference in female participants. P values are from Wald tests (z = β/SE); FDR correction was applied within each group across 19 regions, and significance markers are based on unrounded q values. BMI, body mass index; CDC, Centers for Disease Control and Prevention; FDR, false discovery rate; SE, standard error; q, FDR-corrected P value. *q < .05; **q < .01; ***q < .001. | | | | | | | |

| **eTable 16. Sex-Specific Differences in Regional Brain Structure Between Each CDC Weight Status Group and the Healthy Weight Group.** | | | | | | | | |
| --- | --- | --- | --- | --- | --- | --- | --- | --- |
| group | sex | region | difference | SE | z | p | q | sig |
| Obese | Female | Medial OFC | -0.116 | 0.03 | -3.805 | 0 | 0 | *** |
| Obese | Female | Lateral OFC | -0.203 | 0.031 | -6.595 | 0 | 0 | *** |
| Obese | Female | Temporal pole | -0.147 | 0.033 | -4.484 | 0 | 0 | *** |
| Obese | Female | Superior frontal | -0.143 | 0.032 | -4.428 | 0 | 0 | *** |
| Obese | Female | Rostral mid-frontal | -0.156 | 0.03 | -5.225 | 0 | 0 | *** |
| Obese | Female | Middle temporal | -0.159 | 0.033 | -4.801 | 0 | 0 | *** |
| Obese | Female | Fusiform | -0.121 | 0.033 | -3.696 | 0 | 0.001 | *** |
| Obese | Female | Inferior temporal | -0.086 | 0.033 | -2.612 | 0.009 | 0.021 | * |
| Obese | Female | Lateral occipital | -0.046 | 0.032 | -1.433 | 0.152 | 0.24 |  |
| Obese | Female | Caudal ACC | -0.032 | 0.036 | -0.897 | 0.37 | 0.502 |  |
| Obese | Female | Insula | -0.017 | 0.032 | -0.533 | 0.594 | 0.705 |  |
| Obese | Female | Rostral ACC | -0.003 | 0.033 | -0.084 | 0.933 | 0.933 |  |
| Obese | Female | Pallidum | 0.056 | 0.028 | 2 | 0.045 | 0.096 |  |
| Obese | Female | Amygdala | -0.053 | 0.029 | -1.846 | 0.065 | 0.112 |  |
| Obese | Female | Hippocampus | -0.056 | 0.03 | -1.874 | 0.061 | 0.112 |  |
| Obese | Female | Caudate | -0.047 | 0.034 | -1.39 | 0.164 | 0.24 |  |
| Obese | Female | Putamen | -0.018 | 0.032 | -0.581 | 0.561 | 0.705 |  |
| Obese | Female | Accumbens | 0.009 | 0.03 | 0.298 | 0.766 | 0.808 |  |
| Obese | Female | Thalamus | -0.008 | 0.027 | -0.309 | 0.757 | 0.808 |  |
| Obese | Male | Medial OFC | -0.166 | 0.028 | -5.861 | 0 | 0 | *** |
| Obese | Male | Lateral OFC | -0.184 | 0.029 | -6.427 | 0 | 0 | *** |
| Obese | Male | Temporal pole | -0.162 | 0.03 | -5.335 | 0 | 0 | *** |
| Obese | Male | Superior frontal | -0.221 | 0.03 | -7.343 | 0 | 0 | *** |
| Obese | Male | Rostral mid-frontal | -0.249 | 0.028 | -8.936 | 0 | 0 | *** |
| Obese | Male | Middle temporal | -0.193 | 0.031 | -6.26 | 0 | 0 | *** |
| Obese | Male | Rostral ACC | -0.077 | 0.031 | -2.523 | 0.012 | 0.025 | * |
| Obese | Male | Inferior temporal | -0.07 | 0.031 | -2.285 | 0.022 | 0.042 | * |
| Obese | Male | Fusiform | -0.063 | 0.031 | -2.059 | 0.04 | 0.068 |  |
| Obese | Male | Insula | -0.038 | 0.029 | -1.274 | 0.202 | 0.256 |  |
| Obese | Male | Lateral occipital | -0.03 | 0.03 | -0.999 | 0.318 | 0.377 |  |
| Obese | Male | Caudal ACC | -0.028 | 0.033 | -0.859 | 0.39 | 0.436 |  |
| Obese | Male | Thalamus | 0.072 | 0.025 | 2.87 | 0.004 | 0.011 | * |
| Obese | Male | Pallidum | 0.07 | 0.026 | 2.672 | 0.008 | 0.018 | * |
| Obese | Male | Amygdala | -0.049 | 0.027 | -1.807 | 0.071 | 0.112 |  |
| Obese | Male | Accumbens | 0.045 | 0.028 | 1.623 | 0.105 | 0.153 |  |
| Obese | Male | Putamen | 0.042 | 0.03 | 1.424 | 0.154 | 0.21 |  |
| Obese | Male | Caudate | -0.02 | 0.032 | -0.622 | 0.534 | 0.564 |  |
| Obese | Male | Hippocampus | 0.007 | 0.028 | 0.242 | 0.809 | 0.809 |  |
| Overweight | Female | Rostral mid-frontal | -0.088 | 0.028 | -3.131 | 0.002 | 0.033 | * |
| Overweight | Female | Superior frontal | -0.072 | 0.03 | -2.385 | 0.017 | 0.065 |  |
| Overweight | Female | Insula | 0.072 | 0.03 | 2.424 | 0.015 | 0.065 |  |
| Overweight | Female | Rostral ACC | 0.043 | 0.031 | 1.377 | 0.169 | 0.457 |  |
| Overweight | Female | Medial OFC | -0.017 | 0.029 | -0.6 | 0.548 | 0.633 |  |
| Overweight | Female | Lateral OFC | -0.019 | 0.029 | -0.662 | 0.508 | 0.633 |  |
| Overweight | Female | Temporal pole | -0.023 | 0.031 | -0.745 | 0.456 | 0.633 |  |
| Overweight | Female | Caudal ACC | -0.019 | 0.034 | -0.574 | 0.566 | 0.633 |  |
| Overweight | Female | Inferior temporal | 0.025 | 0.031 | 0.811 | 0.417 | 0.633 |  |
| Overweight | Female | Middle temporal | -0.024 | 0.031 | -0.758 | 0.448 | 0.633 |  |
| Overweight | Female | Lateral occipital | 0.019 | 0.03 | 0.626 | 0.531 | 0.633 |  |
| Overweight | Female | Fusiform | 0.013 | 0.031 | 0.414 | 0.679 | 0.686 |  |
| Overweight | Female | Pallidum | 0.07 | 0.026 | 2.654 | 0.008 | 0.065 |  |
| Overweight | Female | Thalamus | 0.065 | 0.025 | 2.569 | 0.01 | 0.065 |  |
| Overweight | Female | Hippocampus | 0.04 | 0.028 | 1.413 | 0.158 | 0.457 |  |
| Overweight | Female | Amygdala | 0.033 | 0.027 | 1.199 | 0.23 | 0.547 |  |
| Overweight | Female | Accumbens | 0.021 | 0.028 | 0.757 | 0.449 | 0.633 |  |
| Overweight | Female | Caudate | 0.028 | 0.032 | 0.89 | 0.373 | 0.633 |  |
| Overweight | Female | Putamen | 0.012 | 0.03 | 0.404 | 0.686 | 0.686 |  |
| Overweight | Male | Superior frontal | -0.151 | 0.029 | -5.199 | 0 | 0 | *** |
| Overweight | Male | Rostral mid-frontal | -0.125 | 0.027 | -4.646 | 0 | 0 | *** |
| Overweight | Male | Temporal pole | -0.093 | 0.029 | -3.153 | 0.002 | 0.01 | * |
| Overweight | Male | Middle temporal | -0.091 | 0.03 | -3.061 | 0.002 | 0.01 | * |
| Overweight | Male | Lateral OFC | -0.054 | 0.028 | -1.97 | 0.049 | 0.103 |  |
| Overweight | Male | Inferior temporal | -0.057 | 0.03 | -1.911 | 0.056 | 0.106 |  |
| Overweight | Male | Medial OFC | -0.045 | 0.027 | -1.641 | 0.101 | 0.174 |  |
| Overweight | Male | Rostral ACC | 0.006 | 0.03 | 0.208 | 0.835 | 0.999 |  |
| Overweight | Male | Caudal ACC | 0.001 | 0.032 | 0.025 | 0.98 | 0.999 |  |
| Overweight | Male | Insula | 0 | 0.028 | -0.001 | 0.999 | 0.999 |  |
| Overweight | Male | Fusiform | 0.003 | 0.03 | 0.103 | 0.918 | 0.999 |  |
| Overweight | Male | Lateral occipital | 0.005 | 0.029 | 0.188 | 0.851 | 0.999 |  |
| Overweight | Male | Thalamus | 0.065 | 0.024 | 2.695 | 0.007 | 0.027 | * |
| Overweight | Male | Pallidum | 0.058 | 0.025 | 2.29 | 0.022 | 0.07 |  |
| Overweight | Male | Putamen | 0.06 | 0.029 | 2.088 | 0.037 | 0.1 |  |
| Overweight | Male | Accumbens | 0.052 | 0.027 | 1.971 | 0.049 | 0.103 |  |
| Overweight | Male | Hippocampus | 0.029 | 0.027 | 1.091 | 0.275 | 0.436 |  |
| Overweight | Male | Amygdala | -0.005 | 0.026 | -0.177 | 0.86 | 0.999 |  |
| Overweight | Male | Caudate | 0.007 | 0.031 | 0.243 | 0.808 | 0.999 |  |
| Underweight | Female | Rostral ACC | -0.166 | 0.076 | -2.175 | 0.03 | 0.513 |  |
| Underweight | Female | Caudal ACC | -0.144 | 0.081 | -1.773 | 0.076 | 0.513 |  |
| Underweight | Female | Lateral occipital | -0.108 | 0.073 | -1.468 | 0.142 | 0.513 |  |
| Underweight | Female | Medial OFC | -0.06 | 0.071 | -0.857 | 0.391 | 0.627 |  |
| Underweight | Female | Temporal pole | 0.081 | 0.077 | 1.063 | 0.288 | 0.627 |  |
| Underweight | Female | Superior frontal | 0.065 | 0.073 | 0.887 | 0.375 | 0.627 |  |
| Underweight | Female | Rostral mid-frontal | 0.06 | 0.069 | 0.871 | 0.384 | 0.627 |  |
| Underweight | Female | Fusiform | -0.076 | 0.075 | -1.01 | 0.313 | 0.627 |  |
| Underweight | Female | Inferior temporal | -0.04 | 0.075 | -0.529 | 0.596 | 0.81 |  |
| Underweight | Female | Middle temporal | -0.018 | 0.076 | -0.24 | 0.81 | 0.906 |  |
| Underweight | Female | Lateral OFC | -0.012 | 0.071 | -0.172 | 0.864 | 0.912 |  |
| Underweight | Female | Insula | 0.003 | 0.073 | 0.035 | 0.972 | 0.972 |  |
| Underweight | Female | Accumbens | 0.103 | 0.067 | 1.528 | 0.127 | 0.513 |  |
| Underweight | Female | Pallidum | -0.089 | 0.064 | -1.398 | 0.162 | 0.513 |  |
| Underweight | Female | Thalamus | -0.096 | 0.06 | -1.582 | 0.114 | 0.513 |  |
| Underweight | Female | Putamen | -0.06 | 0.07 | -0.849 | 0.396 | 0.627 |  |
| Underweight | Female | Amygdala | -0.044 | 0.065 | -0.669 | 0.503 | 0.736 |  |
| Underweight | Female | Caudate | -0.029 | 0.075 | -0.387 | 0.698 | 0.829 |  |
| Underweight | Female | Hippocampus | -0.029 | 0.066 | -0.443 | 0.657 | 0.829 |  |
| Underweight | Male | Middle temporal | -0.169 | 0.066 | -2.554 | 0.011 | 0.026 | * |
| Underweight | Male | Lateral occipital | -0.164 | 0.064 | -2.55 | 0.011 | 0.026 | * |
| Underweight | Male | Insula | -0.15 | 0.063 | -2.382 | 0.017 | 0.036 | * |
| Underweight | Male | Fusiform | -0.123 | 0.065 | -1.889 | 0.059 | 0.112 |  |
| Underweight | Male | Inferior temporal | -0.077 | 0.066 | -1.177 | 0.239 | 0.379 |  |
| Underweight | Male | Lateral OFC | -0.042 | 0.061 | -0.685 | 0.493 | 0.721 |  |
| Underweight | Male | Caudal ACC | -0.039 | 0.071 | -0.548 | 0.584 | 0.761 |  |
| Underweight | Male | Rostral mid-frontal | 0.031 | 0.06 | 0.523 | 0.601 | 0.761 |  |
| Underweight | Male | Medial OFC | 0.022 | 0.061 | 0.37 | 0.711 | 0.845 |  |
| Underweight | Male | Temporal pole | -0.019 | 0.065 | -0.285 | 0.776 | 0.867 |  |
| Underweight | Male | Rostral ACC | -0.014 | 0.066 | -0.207 | 0.836 | 0.882 |  |
| Underweight | Male | Superior frontal | 0.007 | 0.064 | 0.103 | 0.918 | 0.918 |  |
| Underweight | Male | Hippocampus | -0.263 | 0.059 | -4.431 | 0 | 0 | *** |
| Underweight | Male | Pallidum | -0.224 | 0.056 | -4.03 | 0 | 0 | *** |
| Underweight | Male | Thalamus | -0.23 | 0.053 | -4.312 | 0 | 0 | *** |
| Underweight | Male | Accumbens | -0.218 | 0.059 | -3.709 | 0 | 0.001 | *** |
| Underweight | Male | Putamen | -0.22 | 0.063 | -3.496 | 0 | 0.002 | ** |
| Underweight | Male | Caudate | -0.178 | 0.068 | -2.635 | 0.008 | 0.026 | * |
| Underweight | Male | Amygdala | -0.086 | 0.058 | -1.487 | 0.137 | 0.237 |  |
| Note. Sex-specific differences between each CDC weight status group and the healthy weight reference group were estimated by refitting the group × sex model (eTable 15) with each sex as the reference in turn, in SD units of each outcome. P values are from Wald tests (z = difference/SE); FDR correction was applied within each group and sex across 19 regions, and significance markers are based on unrounded q values. BMI, body mass index; CDC, Centers for Disease Control and Prevention; FDR, false discovery rate; SE, standard error; q, FDR-corrected P value. *q < .05; **q < .01; ***q < .001. | | | | | | | | |

| **eTable 17. Stratified Between-Person, Within-Person, and Contextual Associations of Extended BMI-for-Age z With Regional Brain Structure by CDC Weight Status Group.** | | | | | | | |
| --- | --- | --- | --- | --- | --- | --- | --- |
| group | region | β | SE | z | p | q | sig |
| **Section A:** Between-person (cross-sectional) associations | | | | | | | |
| Healthy weight | Lateral OFC | -0.039 | 0.014 | -2.803 | 0.005 | 0.012 | * |
| Healthy weight | Rostral mid-frontal | -0.037 | 0.014 | -2.693 | 0.007 | 0.015 | * |
| Healthy weight | Lateral occipital | 0.036 | 0.015 | 2.405 | 0.016 | 0.031 | * |
| Healthy weight | Superior frontal | -0.033 | 0.015 | -2.249 | 0.024 | 0.042 | * |
| Healthy weight | Fusiform | 0.031 | 0.015 | 2.057 | 0.04 | 0.063 |  |
| Healthy weight | Inferior temporal | 0.03 | 0.015 | 1.959 | 0.05 | 0.073 |  |
| Healthy weight | Medial OFC | -0.022 | 0.014 | -1.54 | 0.124 | 0.168 |  |
| Healthy weight | Insula | 0.018 | 0.015 | 1.201 | 0.23 | 0.291 |  |
| Healthy weight | Temporal pole | -0.017 | 0.015 | -1.131 | 0.258 | 0.306 |  |
| Healthy weight | Middle temporal | 0.012 | 0.015 | 0.827 | 0.408 | 0.456 |  |
| Healthy weight | Caudal ACC | -0.009 | 0.016 | -0.546 | 0.585 | 0.618 |  |
| Healthy weight | Rostral ACC | 0.005 | 0.015 | 0.331 | 0.74 | 0.74 |  |
| Healthy weight | Accumbens | 0.06 | 0.013 | 4.461 | 0 | 0 | *** |
| Healthy weight | Amygdala | 0.054 | 0.013 | 4.064 | 0 | 0 | *** |
| Healthy weight | Caudate | 0.061 | 0.016 | 3.962 | 0 | 0 | *** |
| Healthy weight | Hippocampus | 0.075 | 0.014 | 5.521 | 0 | 0 | *** |
| Healthy weight | Pallidum | 0.072 | 0.013 | 5.694 | 0 | 0 | *** |
| Healthy weight | Putamen | 0.066 | 0.015 | 4.573 | 0 | 0 | *** |
| Healthy weight | Thalamus | 0.089 | 0.012 | 7.446 | 0 | 0 | *** |
| Overweight | Rostral ACC | -0.134 | 0.108 | -1.241 | 0.215 | 0.816 |  |
| Overweight | Rostral mid-frontal | -0.128 | 0.099 | -1.29 | 0.197 | 0.816 |  |
| Overweight | Insula | 0.135 | 0.103 | 1.315 | 0.188 | 0.816 |  |
| Overweight | Lateral occipital | -0.154 | 0.108 | -1.434 | 0.152 | 0.816 |  |
| Overweight | Lateral OFC | -0.052 | 0.1 | -0.518 | 0.605 | 0.884 |  |
| Overweight | Temporal pole | -0.059 | 0.108 | -0.544 | 0.586 | 0.884 |  |
| Overweight | Caudal ACC | 0.061 | 0.115 | 0.529 | 0.597 | 0.884 |  |
| Overweight | Superior frontal | -0.068 | 0.108 | -0.63 | 0.529 | 0.884 |  |
| Overweight | Inferior temporal | -0.059 | 0.11 | -0.535 | 0.593 | 0.884 |  |
| Overweight | Middle temporal | -0.11 | 0.11 | -0.997 | 0.319 | 0.884 |  |
| Overweight | Fusiform | -0.045 | 0.109 | -0.408 | 0.684 | 0.928 |  |
| Overweight | Medial OFC | -0.005 | 0.1 | -0.05 | 0.96 | 0.992 |  |
| Overweight | Amygdala | -0.172 | 0.098 | -1.759 | 0.079 | 0.816 |  |
| Overweight | Hippocampus | -0.082 | 0.1 | -0.817 | 0.414 | 0.884 |  |
| Overweight | Putamen | -0.069 | 0.107 | -0.648 | 0.517 | 0.884 |  |
| Overweight | Caudate | 0.035 | 0.118 | 0.294 | 0.769 | 0.934 |  |
| Overweight | Pallidum | -0.026 | 0.095 | -0.271 | 0.786 | 0.934 |  |
| Overweight | Thalamus | -0.019 | 0.094 | -0.207 | 0.836 | 0.934 |  |
| Overweight | Accumbens | 0.001 | 0.101 | 0.01 | 0.992 | 0.992 |  |
| Obese | Medial OFC | -0.173 | 0.038 | -4.623 | 0 | 0 | *** |
| Obese | Lateral OFC | -0.206 | 0.039 | -5.358 | 0 | 0 | *** |
| Obese | Temporal pole | -0.218 | 0.042 | -5.139 | 0 | 0 | *** |
| Obese | Superior frontal | -0.164 | 0.04 | -4.068 | 0 | 0 | *** |
| Obese | Rostral mid-frontal | -0.139 | 0.037 | -3.7 | 0 | 0 | *** |
| Obese | Insula | -0.197 | 0.039 | -5.01 | 0 | 0 | *** |
| Obese | Fusiform | -0.27 | 0.041 | -6.609 | 0 | 0 | *** |
| Obese | Inferior temporal | -0.258 | 0.041 | -6.28 | 0 | 0 | *** |
| Obese | Middle temporal | -0.221 | 0.043 | -5.182 | 0 | 0 | *** |
| Obese | Lateral occipital | -0.208 | 0.04 | -5.218 | 0 | 0 | *** |
| Obese | Rostral ACC | -0.11 | 0.041 | -2.688 | 0.007 | 0.012 | * |
| Obese | Caudal ACC | -0.034 | 0.044 | -0.776 | 0.438 | 0.555 |  |
| Obese | Amygdala | -0.086 | 0.036 | -2.425 | 0.015 | 0.024 | * |
| Obese | Caudate | -0.064 | 0.043 | -1.489 | 0.137 | 0.2 |  |
| Obese | Putamen | -0.049 | 0.04 | -1.241 | 0.215 | 0.291 |  |
| Obese | Hippocampus | -0.019 | 0.037 | -0.515 | 0.607 | 0.72 |  |
| Obese | Thalamus | -0.013 | 0.034 | -0.385 | 0.701 | 0.783 |  |
| Obese | Accumbens | 0.012 | 0.037 | 0.329 | 0.742 | 0.784 |  |
| Obese | Pallidum | -0.007 | 0.036 | -0.195 | 0.846 | 0.846 |  |
| Underweight | Lateral occipital | 0.211 | 0.082 | 2.57 | 0.01 | 0.193 |  |
| Underweight | Fusiform | 0.191 | 0.085 | 2.253 | 0.024 | 0.23 |  |
| Underweight | Medial OFC | 0.139 | 0.08 | 1.736 | 0.082 | 0.281 |  |
| Underweight | Superior frontal | 0.138 | 0.074 | 1.874 | 0.061 | 0.281 |  |
| Underweight | Rostral mid-frontal | 0.124 | 0.073 | 1.703 | 0.089 | 0.281 |  |
| Underweight | Middle temporal | 0.161 | 0.087 | 1.855 | 0.064 | 0.281 |  |
| Underweight | Inferior temporal | 0.131 | 0.083 | 1.581 | 0.114 | 0.309 |  |
| Underweight | Lateral OFC | 0.096 | 0.074 | 1.293 | 0.196 | 0.414 |  |
| Underweight | Rostral ACC | 0.085 | 0.086 | 0.99 | 0.322 | 0.513 |  |
| Underweight | Insula | 0.058 | 0.08 | 0.721 | 0.471 | 0.596 |  |
| Underweight | Caudal ACC | 0.041 | 0.089 | 0.464 | 0.643 | 0.763 |  |
| Underweight | Temporal pole | -0.023 | 0.09 | -0.257 | 0.797 | 0.843 |  |
| Underweight | Accumbens | -0.105 | 0.073 | -1.446 | 0.148 | 0.352 |  |
| Underweight | Putamen | -0.089 | 0.079 | -1.124 | 0.261 | 0.496 |  |
| Underweight | Amygdala | -0.07 | 0.071 | -0.987 | 0.324 | 0.513 |  |
| Underweight | Hippocampus | -0.068 | 0.077 | -0.876 | 0.381 | 0.557 |  |
| Underweight | Pallidum | -0.057 | 0.071 | -0.809 | 0.418 | 0.568 |  |
| Underweight | Thalamus | 0.017 | 0.067 | 0.255 | 0.799 | 0.843 |  |
| Underweight | Caudate | 0.01 | 0.087 | 0.119 | 0.905 | 0.905 |  |
| **Section B:** Within-person (longitudinal) associations | | | | | | | |
| Healthy weight | Middle temporal | 0.03 | 0.007 | 4 | 0 | 0 | *** |
| Healthy weight | Lateral occipital | 0.036 | 0.007 | 4.979 | 0 | 0 | *** |
| Healthy weight | Fusiform | 0.02 | 0.008 | 2.647 | 0.008 | 0.026 | * |
| Healthy weight | Inferior temporal | 0.014 | 0.008 | 1.911 | 0.056 | 0.133 |  |
| Healthy weight | Temporal pole | -0.018 | 0.011 | -1.601 | 0.109 | 0.231 |  |
| Healthy weight | Rostral mid-frontal | -0.011 | 0.008 | -1.436 | 0.151 | 0.287 |  |
| Healthy weight | Medial OFC | 0.011 | 0.009 | 1.212 | 0.226 | 0.357 |  |
| Healthy weight | Caudal ACC | 0.008 | 0.006 | 1.231 | 0.218 | 0.357 |  |
| Healthy weight | Rostral ACC | 0.01 | 0.009 | 1.041 | 0.298 | 0.377 |  |
| Healthy weight | Superior frontal | -0.008 | 0.008 | -1.083 | 0.279 | 0.377 |  |
| Healthy weight | Lateral OFC | -0.007 | 0.008 | -0.842 | 0.4 | 0.462 |  |
| Healthy weight | Insula | 0.005 | 0.01 | 0.501 | 0.617 | 0.651 |  |
| Healthy weight | Hippocampus | 0.021 | 0.004 | 5.443 | 0 | 0 | *** |
| Healthy weight | Thalamus | 0.022 | 0.005 | 4.018 | 0 | 0 | *** |
| Healthy weight | Putamen | 0.013 | 0.004 | 3.295 | 0.001 | 0.004 | ** |
| Healthy weight | Pallidum | 0.02 | 0.008 | 2.583 | 0.01 | 0.027 | * |
| Healthy weight | Caudate | 0.004 | 0.003 | 1.104 | 0.27 | 0.377 |  |
| Healthy weight | Amygdala | 0.005 | 0.006 | 0.818 | 0.413 | 0.462 |  |
| Healthy weight | Accumbens | 0 | 0.007 | -0.025 | 0.98 | 0.98 |  |
| Overweight | Temporal pole | -0.076 | 0.029 | -2.576 | 0.01 | 0.095 |  |
| Overweight | Medial OFC | -0.04 | 0.023 | -1.784 | 0.074 | 0.226 |  |
| Overweight | Rostral ACC | -0.045 | 0.023 | -1.933 | 0.053 | 0.226 |  |
| Overweight | Caudal ACC | -0.027 | 0.016 | -1.705 | 0.088 | 0.226 |  |
| Overweight | Superior frontal | -0.041 | 0.02 | -1.993 | 0.046 | 0.226 |  |
| Overweight | Rostral mid-frontal | -0.037 | 0.021 | -1.728 | 0.084 | 0.226 |  |
| Overweight | Lateral OFC | -0.032 | 0.022 | -1.426 | 0.154 | 0.325 |  |
| Overweight | Middle temporal | 0.018 | 0.021 | 0.869 | 0.385 | 0.678 |  |
| Overweight | Insula | -0.016 | 0.025 | -0.631 | 0.528 | 0.779 |  |
| Overweight | Inferior temporal | 0.007 | 0.021 | 0.318 | 0.75 | 0.792 |  |
| Overweight | Lateral occipital | 0.006 | 0.019 | 0.32 | 0.749 | 0.792 |  |
| Overweight | Fusiform | -0.003 | 0.02 | -0.13 | 0.896 | 0.896 |  |
| Overweight | Thalamus | 0.038 | 0.014 | 2.692 | 0.007 | 0.095 |  |
| Overweight | Hippocampus | 0.016 | 0.01 | 1.669 | 0.095 | 0.226 |  |
| Overweight | Amygdala | -0.013 | 0.015 | -0.855 | 0.393 | 0.678 |  |
| Overweight | Pallidum | -0.012 | 0.019 | -0.624 | 0.533 | 0.779 |  |
| Overweight | Accumbens | -0.009 | 0.019 | -0.495 | 0.62 | 0.788 |  |
| Overweight | Putamen | 0.005 | 0.01 | 0.493 | 0.622 | 0.788 |  |
| Overweight | Caudate | 0.003 | 0.008 | 0.362 | 0.718 | 0.792 |  |
| Obese | Insula | -0.086 | 0.029 | -2.992 | 0.003 | 0.053 |  |
| Obese | Temporal pole | -0.098 | 0.037 | -2.655 | 0.008 | 0.075 |  |
| Obese | Lateral OFC | -0.045 | 0.028 | -1.639 | 0.101 | 0.277 |  |
| Obese | Rostral ACC | -0.043 | 0.026 | -1.642 | 0.101 | 0.277 |  |
| Obese | Inferior temporal | -0.044 | 0.025 | -1.745 | 0.081 | 0.277 |  |
| Obese | Rostral mid-frontal | 0.035 | 0.025 | 1.402 | 0.161 | 0.299 |  |
| Obese | Fusiform | -0.035 | 0.026 | -1.39 | 0.165 | 0.299 |  |
| Obese | Lateral occipital | -0.024 | 0.022 | -1.085 | 0.278 | 0.44 |  |
| Obese | Medial OFC | 0.02 | 0.027 | 0.742 | 0.458 | 0.58 |  |
| Obese | Caudal ACC | -0.014 | 0.018 | -0.79 | 0.43 | 0.58 |  |
| Obese | Superior frontal | -0.016 | 0.024 | -0.655 | 0.513 | 0.609 |  |
| Obese | Middle temporal | -0.004 | 0.024 | -0.162 | 0.872 | 0.872 |  |
| Obese | Caudate | -0.019 | 0.01 | -2.043 | 0.041 | 0.26 |  |
| Obese | Putamen | -0.02 | 0.012 | -1.636 | 0.102 | 0.277 |  |
| Obese | Amygdala | -0.026 | 0.017 | -1.499 | 0.134 | 0.299 |  |
| Obese | Thalamus | 0.022 | 0.016 | 1.362 | 0.173 | 0.299 |  |
| Obese | Hippocampus | 0.009 | 0.011 | 0.816 | 0.415 | 0.58 |  |
| Obese | Accumbens | -0.011 | 0.022 | -0.511 | 0.609 | 0.681 |  |
| Obese | Pallidum | 0.006 | 0.023 | 0.252 | 0.801 | 0.845 |  |
| Underweight | Medial OFC | 0.03 | 0.024 | 1.268 | 0.205 | 0.778 |  |
| Underweight | Fusiform | -0.027 | 0.021 | -1.269 | 0.204 | 0.778 |  |
| Underweight | Lateral OFC | 0.02 | 0.023 | 0.869 | 0.385 | 0.876 |  |
| Underweight | Temporal pole | 0.025 | 0.03 | 0.815 | 0.415 | 0.876 |  |
| Underweight | Rostral ACC | 0.02 | 0.024 | 0.826 | 0.409 | 0.876 |  |
| Underweight | Caudal ACC | 0.003 | 0.016 | 0.166 | 0.868 | 0.97 |  |
| Underweight | Superior frontal | 0.004 | 0.02 | 0.215 | 0.83 | 0.97 |  |
| Underweight | Rostral mid-frontal | -0.009 | 0.019 | -0.461 | 0.645 | 0.97 |  |
| Underweight | Insula | -0.01 | 0.028 | -0.365 | 0.715 | 0.97 |  |
| Underweight | Inferior temporal | 0.003 | 0.02 | 0.166 | 0.868 | 0.97 |  |
| Underweight | Middle temporal | -0.009 | 0.021 | -0.445 | 0.657 | 0.97 |  |
| Underweight | Lateral occipital | -0.004 | 0.019 | -0.208 | 0.835 | 0.97 |  |
| Underweight | Pallidum | 0.03 | 0.021 | 1.396 | 0.163 | 0.778 |  |
| Underweight | Putamen | -0.016 | 0.011 | -1.407 | 0.159 | 0.778 |  |
| Underweight | Thalamus | 0.022 | 0.014 | 1.557 | 0.119 | 0.778 |  |
| Underweight | Accumbens | -0.02 | 0.02 | -0.977 | 0.329 | 0.876 |  |
| Underweight | Amygdala | 0.004 | 0.017 | 0.235 | 0.814 | 0.97 |  |
| Underweight | Caudate | 0 | 0.008 | -0.029 | 0.977 | 0.986 |  |
| Underweight | Hippocampus | 0 | 0.009 | -0.017 | 0.986 | 0.986 |  |
| **Section C:** Contextual test (between- minus within-person difference) | | | | | | | |
| Healthy weight | Medial OFC | -0.032 | 0.016 | -1.962 | 0.05 | 0.105 |  |
| Healthy weight | Lateral OFC | -0.032 | 0.016 | -1.985 | 0.047 | 0.105 |  |
| Healthy weight | Rostral mid-frontal | -0.026 | 0.016 | -1.626 | 0.104 | 0.198 |  |
| Healthy weight | Superior frontal | -0.025 | 0.017 | -1.497 | 0.135 | 0.232 |  |
| Healthy weight | Middle temporal | -0.018 | 0.017 | -1.063 | 0.288 | 0.456 |  |
| Healthy weight | Caudal ACC | -0.016 | 0.017 | -0.947 | 0.344 | 0.499 |  |
| Healthy weight | Inferior temporal | 0.015 | 0.017 | 0.901 | 0.368 | 0.499 |  |
| Healthy weight | Insula | 0.012 | 0.018 | 0.701 | 0.484 | 0.613 |  |
| Healthy weight | Fusiform | 0.011 | 0.017 | 0.647 | 0.518 | 0.615 |  |
| Healthy weight | Rostral ACC | -0.005 | 0.018 | -0.264 | 0.791 | 0.885 |  |
| Healthy weight | Temporal pole | 0.001 | 0.018 | 0.054 | 0.957 | 0.979 |  |
| Healthy weight | Lateral occipital | 0 | 0.016 | -0.026 | 0.979 | 0.979 |  |
| Healthy weight | Thalamus | 0.067 | 0.013 | 5.137 | 0 | 0 | *** |
| Healthy weight | Accumbens | 0.06 | 0.015 | 3.949 | 0 | 0.001 | *** |
| Healthy weight | Caudate | 0.058 | 0.016 | 3.661 | 0 | 0.001 | ** |
| Healthy weight | Hippocampus | 0.054 | 0.014 | 3.849 | 0 | 0.001 | *** |
| Healthy weight | Pallidum | 0.053 | 0.015 | 3.572 | 0 | 0.001 | ** |
| Healthy weight | Putamen | 0.053 | 0.015 | 3.527 | 0 | 0.001 | ** |
| Healthy weight | Amygdala | 0.049 | 0.014 | 3.399 | 0.001 | 0.002 | ** |
| Overweight | Medial OFC | 0.035 | 0.103 | 0.344 | 0.731 | 0.92 |  |
| Overweight | Lateral OFC | -0.02 | 0.103 | -0.197 | 0.844 | 0.92 |  |
| Overweight | Temporal pole | 0.017 | 0.112 | 0.15 | 0.88 | 0.92 |  |
| Overweight | Rostral ACC | -0.089 | 0.11 | -0.809 | 0.418 | 0.92 |  |
| Overweight | Caudal ACC | 0.089 | 0.117 | 0.76 | 0.447 | 0.92 |  |
| Overweight | Superior frontal | -0.027 | 0.11 | -0.247 | 0.805 | 0.92 |  |
| Overweight | Rostral mid-frontal | -0.091 | 0.101 | -0.896 | 0.37 | 0.92 |  |
| Overweight | Insula | 0.151 | 0.106 | 1.429 | 0.153 | 0.92 |  |
| Overweight | Fusiform | -0.042 | 0.111 | -0.377 | 0.706 | 0.92 |  |
| Overweight | Inferior temporal | -0.065 | 0.111 | -0.585 | 0.559 | 0.92 |  |
| Overweight | Middle temporal | -0.128 | 0.112 | -1.14 | 0.254 | 0.92 |  |
| Overweight | Lateral occipital | -0.16 | 0.109 | -1.468 | 0.142 | 0.92 |  |
| Overweight | Accumbens | 0.01 | 0.103 | 0.1 | 0.92 | 0.92 |  |
| Overweight | Amygdala | -0.159 | 0.099 | -1.605 | 0.108 | 0.92 |  |
| Overweight | Caudate | 0.032 | 0.118 | 0.269 | 0.788 | 0.92 |  |
| Overweight | Hippocampus | -0.098 | 0.101 | -0.971 | 0.331 | 0.92 |  |
| Overweight | Pallidum | -0.014 | 0.097 | -0.145 | 0.885 | 0.92 |  |
| Overweight | Putamen | -0.074 | 0.107 | -0.691 | 0.489 | 0.92 |  |
| Overweight | Thalamus | -0.057 | 0.095 | -0.603 | 0.547 | 0.92 |  |
| Obese | Medial OFC | -0.193 | 0.046 | -4.185 | 0 | 0 | *** |
| Obese | Rostral mid-frontal | -0.174 | 0.045 | -3.844 | 0 | 0 | *** |
| Obese | Fusiform | -0.234 | 0.048 | -4.855 | 0 | 0 | *** |
| Obese | Inferior temporal | -0.214 | 0.048 | -4.451 | 0 | 0 | *** |
| Obese | Middle temporal | -0.218 | 0.049 | -4.42 | 0 | 0 | *** |
| Obese | Lateral occipital | -0.184 | 0.046 | -4.039 | 0 | 0 | *** |
| Obese | Lateral OFC | -0.161 | 0.047 | -3.399 | 0.001 | 0.002 | ** |
| Obese | Superior frontal | -0.148 | 0.047 | -3.165 | 0.002 | 0.004 | ** |
| Obese | Insula | -0.111 | 0.049 | -2.268 | 0.023 | 0.049 | * |
| Obese | Temporal pole | -0.12 | 0.056 | -2.125 | 0.034 | 0.064 |  |
| Obese | Rostral ACC | -0.067 | 0.049 | -1.369 | 0.171 | 0.271 |  |
| Obese | Caudal ACC | -0.02 | 0.047 | -0.422 | 0.673 | 0.71 |  |
| Obese | Amygdala | -0.06 | 0.04 | -1.518 | 0.129 | 0.223 |  |
| Obese | Caudate | -0.044 | 0.044 | -1.007 | 0.314 | 0.459 |  |
| Obese | Thalamus | -0.035 | 0.037 | -0.927 | 0.354 | 0.481 |  |
| Obese | Hippocampus | -0.028 | 0.038 | -0.734 | 0.463 | 0.568 |  |
| Obese | Putamen | -0.029 | 0.042 | -0.709 | 0.478 | 0.568 |  |
| Obese | Accumbens | 0.023 | 0.043 | 0.54 | 0.589 | 0.658 |  |
| Obese | Pallidum | -0.013 | 0.043 | -0.299 | 0.765 | 0.765 |  |
| Underweight | Fusiform | 0.218 | 0.087 | 2.492 | 0.013 | 0.121 |  |
| Underweight | Lateral occipital | 0.215 | 0.084 | 2.55 | 0.011 | 0.121 |  |
| Underweight | Superior frontal | 0.134 | 0.076 | 1.753 | 0.08 | 0.302 |  |
| Underweight | Rostral mid-frontal | 0.133 | 0.075 | 1.764 | 0.078 | 0.302 |  |
| Underweight | Middle temporal | 0.17 | 0.089 | 1.907 | 0.056 | 0.302 |  |
| Underweight | Inferior temporal | 0.128 | 0.085 | 1.497 | 0.134 | 0.425 |  |
| Underweight | Medial OFC | 0.109 | 0.084 | 1.308 | 0.191 | 0.518 |  |
| Underweight | Lateral OFC | 0.076 | 0.078 | 0.975 | 0.329 | 0.564 |  |
| Underweight | Insula | 0.068 | 0.085 | 0.801 | 0.423 | 0.574 |  |
| Underweight | Rostral ACC | 0.065 | 0.089 | 0.725 | 0.468 | 0.593 |  |
| Underweight | Temporal pole | -0.048 | 0.095 | -0.501 | 0.616 | 0.732 |  |
| Underweight | Caudal ACC | 0.038 | 0.09 | 0.426 | 0.67 | 0.749 |  |
| Underweight | Accumbens | -0.086 | 0.076 | -1.133 | 0.257 | 0.543 |  |
| Underweight | Pallidum | -0.087 | 0.074 | -1.175 | 0.24 | 0.543 |  |
| Underweight | Amygdala | -0.074 | 0.073 | -1.014 | 0.31 | 0.564 |  |
| Underweight | Hippocampus | -0.067 | 0.078 | -0.868 | 0.386 | 0.564 |  |
| Underweight | Putamen | -0.073 | 0.08 | -0.915 | 0.36 | 0.564 |  |
| Underweight | Caudate | 0.011 | 0.087 | 0.121 | 0.903 | 0.938 |  |
| Underweight | Thalamus | -0.005 | 0.068 | -0.077 | 0.938 | 0.938 |  |
| Note. Within-between linear mixed-effects models were fitted separately within each CDC weight status group, defined by each participant's mean extended BMI-for-age z across included visits. Section A shows between-person coefficients (stable differences among youth within the same weight status category); Section B, within-person coefficients (change within individuals across visits); and Section C, contextual effects (between- minus within-person coefficient, estimated as a linear contrast from the model covariance matrix). Models were adjusted for age, age², sex, pubertal development, socioeconomic status, race and ethnicity, and intracranial volume (subcortical outcomes), with random intercepts for family and participant. β is the standardized coefficient (SD change in the outcome per 1-SD increase in BMI-for-age z). P values are from Wald tests (z = β/SE); FDR correction was applied within each group and estimate type across 19 regions, and significance markers are based on unrounded *q* values. BMI, body mass index; CDC, Centers for Disease Control and Prevention; FDR, false discovery rate; SE, standard error; *q, FDR-corrected P value*. **q* < .05; ***q* < .01; ****q* < .001. | | | | | | | |

| **eTable 18. Formal Group Comparison: Interaction Tests for Between-Person and Within-Person BMI-for-Age z Associations Versus the Healthy Weight Reference Group.** | | | | | | | |
| --- | --- | --- | --- | --- | --- | --- | --- |
| measure | region | Δβ | SE | z | p | q | sig |
| **Section A:** Between-person, obesity vs healthy weight | | | | | | | |
| Cortical thickness | Lateral OFC | -0.153 | 0.04 | -3.869 | 0 | 0 | *** |
| Cortical thickness | Temporal pole | -0.184 | 0.042 | -4.323 | 0 | 0 | *** |
| Cortical thickness | Insula | -0.206 | 0.041 | -5.035 | 0 | 0 | *** |
| Cortical thickness | Fusiform | -0.278 | 0.042 | -6.63 | 0 | 0 | *** |
| Cortical thickness | Inferior temporal | -0.262 | 0.042 | -6.216 | 0 | 0 | *** |
| Cortical thickness | Middle temporal | -0.204 | 0.042 | -4.814 | 0 | 0 | *** |
| Cortical thickness | Lateral occipital | -0.233 | 0.041 | -5.631 | 0 | 0 | *** |
| Cortical thickness | Medial OFC | -0.141 | 0.039 | -3.586 | 0 | 0.001 | *** |
| Cortical thickness | Rostral ACC | -0.117 | 0.042 | -2.76 | 0.006 | 0.011 | * |
| Cortical thickness | Superior frontal | -0.097 | 0.041 | -2.349 | 0.019 | 0.027 | * |
| Cortical thickness | Rostral mid-frontal | -0.077 | 0.039 | -2.011 | 0.044 | 0.053 |  |
| Cortical thickness | Caudal ACC | -0.026 | 0.045 | -0.573 | 0.567 | 0.567 |  |
| Subcortical volume | Amygdala | -0.127 | 0.037 | -3.438 | 0.001 | 0.001 | ** |
| Subcortical volume | Putamen | -0.106 | 0.04 | -2.623 | 0.009 | 0.015 | * |
| Subcortical volume | Thalamus | -0.081 | 0.034 | -2.367 | 0.018 | 0.027 | * |
| Subcortical volume | Hippocampus | -0.088 | 0.038 | -2.306 | 0.021 | 0.029 | * |
| Subcortical volume | Caudate | -0.089 | 0.043 | -2.05 | 0.04 | 0.051 |  |
| Subcortical volume | Pallidum | -0.059 | 0.036 | -1.634 | 0.102 | 0.114 |  |
| Subcortical volume | Accumbens | -0.023 | 0.038 | -0.6 | 0.548 | 0.567 |  |
| **Section B:** Between-person, overweight vs healthy weight | | | | | | | |
| Cortical thickness | Lateral occipital | -0.194 | 0.105 | -1.847 | 0.065 | 0.41 |  |
| Cortical thickness | Middle temporal | -0.153 | 0.108 | -1.412 | 0.158 | 0.5 |  |
| Cortical thickness | Insula | 0.118 | 0.103 | 1.146 | 0.252 | 0.598 |  |
| Cortical thickness | Inferior temporal | -0.129 | 0.108 | -1.199 | 0.231 | 0.598 |  |
| Cortical thickness | Rostral ACC | -0.112 | 0.108 | -1.03 | 0.303 | 0.64 |  |
| Cortical thickness | Fusiform | -0.093 | 0.107 | -0.868 | 0.385 | 0.666 |  |
| Cortical thickness | Rostral mid-frontal | -0.077 | 0.098 | -0.787 | 0.431 | 0.683 |  |
| Cortical thickness | Caudal ACC | 0.076 | 0.117 | 0.65 | 0.516 | 0.7 |  |
| Cortical thickness | Lateral OFC | -0.042 | 0.1 | -0.422 | 0.673 | 0.853 |  |
| Cortical thickness | Temporal pole | -0.021 | 0.107 | -0.193 | 0.847 | 0.921 |  |
| Cortical thickness | Superior frontal | -0.017 | 0.105 | -0.16 | 0.873 | 0.921 |  |
| Cortical thickness | Medial OFC | -0.001 | 0.1 | -0.013 | 0.989 | 0.989 |  |
| Subcortical volume | Amygdala | -0.208 | 0.094 | -2.205 | 0.027 | 0.41 |  |
| Subcortical volume | Hippocampus | -0.186 | 0.097 | -1.912 | 0.056 | 0.41 |  |
| Subcortical volume | Putamen | -0.162 | 0.103 | -1.578 | 0.115 | 0.435 |  |
| Subcortical volume | Thalamus | -0.144 | 0.087 | -1.647 | 0.1 | 0.435 |  |
| Subcortical volume | Pallidum | -0.079 | 0.091 | -0.868 | 0.385 | 0.666 |  |
| Subcortical volume | Accumbens | -0.067 | 0.096 | -0.7 | 0.484 | 0.7 |  |
| Subcortical volume | Caudate | -0.031 | 0.111 | -0.276 | 0.783 | 0.921 |  |
| **Section C:** Between-person, underweight vs healthy weight | | | | | | | |
| Cortical thickness | Lateral occipital | 0.153 | 0.082 | 1.859 | 0.063 | 0.171 |  |
| Cortical thickness | Superior frontal | 0.145 | 0.083 | 1.754 | 0.079 | 0.189 |  |
| Cortical thickness | Medial OFC | 0.131 | 0.08 | 1.628 | 0.104 | 0.197 |  |
| Cortical thickness | Rostral mid-frontal | 0.129 | 0.078 | 1.653 | 0.098 | 0.197 |  |
| Cortical thickness | Fusiform | 0.117 | 0.085 | 1.384 | 0.166 | 0.287 |  |
| Cortical thickness | Middle temporal | 0.113 | 0.085 | 1.325 | 0.185 | 0.293 |  |
| Cortical thickness | Inferior temporal | 0.091 | 0.085 | 1.064 | 0.287 | 0.42 |  |
| Cortical thickness | Lateral OFC | 0.069 | 0.08 | 0.86 | 0.39 | 0.529 |  |
| Cortical thickness | Rostral ACC | 0.056 | 0.087 | 0.648 | 0.517 | 0.655 |  |
| Cortical thickness | Temporal pole | -0.042 | 0.088 | -0.48 | 0.631 | 0.73 |  |
| Cortical thickness | Caudal ACC | 0.041 | 0.091 | 0.45 | 0.653 | 0.73 |  |
| Cortical thickness | Insula | 0.031 | 0.084 | 0.37 | 0.712 | 0.751 |  |
| Subcortical volume | Hippocampus | -0.172 | 0.074 | -2.335 | 0.02 | 0.159 |  |
| Subcortical volume | Pallidum | -0.171 | 0.072 | -2.364 | 0.018 | 0.159 |  |
| Subcortical volume | Putamen | -0.174 | 0.078 | -2.239 | 0.025 | 0.159 |  |
| Subcortical volume | Accumbens | -0.142 | 0.076 | -1.87 | 0.062 | 0.171 |  |
| Subcortical volume | Amygdala | -0.146 | 0.073 | -2.001 | 0.045 | 0.171 |  |
| Subcortical volume | Thalamus | -0.136 | 0.068 | -2.013 | 0.044 | 0.171 |  |
| Subcortical volume | Caudate | -0.006 | 0.083 | -0.068 | 0.946 | 0.946 |  |
| **Section D:** Within-person, obesity vs healthy weight | | | | | | | |
| Cortical thickness | Insula | -0.092 | 0.031 | -3.007 | 0.003 | 0.018 | * |
| Cortical thickness | Fusiform | -0.068 | 0.024 | -2.898 | 0.004 | 0.018 | * |
| Cortical thickness | Lateral occipital | -0.065 | 0.022 | -2.945 | 0.003 | 0.018 | * |
| Cortical thickness | Temporal pole | -0.093 | 0.034 | -2.709 | 0.007 | 0.021 | * |
| Cortical thickness | Inferior temporal | -0.065 | 0.023 | -2.771 | 0.006 | 0.021 | * |
| Cortical thickness | Rostral mid-frontal | 0.056 | 0.024 | 2.314 | 0.021 | 0.049 | * |
| Cortical thickness | Rostral ACC | -0.053 | 0.028 | -1.911 | 0.056 | 0.118 |  |
| Cortical thickness | Lateral OFC | -0.046 | 0.026 | -1.78 | 0.075 | 0.141 |  |
| Cortical thickness | Middle temporal | -0.039 | 0.023 | -1.697 | 0.09 | 0.142 |  |
| Cortical thickness | Medial OFC | 0.012 | 0.027 | 0.46 | 0.646 | 0.818 |  |
| Cortical thickness | Caudal ACC | -0.007 | 0.019 | -0.379 | 0.705 | 0.832 |  |
| Cortical thickness | Superior frontal | 0.007 | 0.024 | 0.295 | 0.768 | 0.832 |  |
| Subcortical volume | Putamen | -0.038 | 0.012 | -3.111 | 0.002 | 0.018 | * |
| Subcortical volume | Caudate | -0.024 | 0.01 | -2.493 | 0.013 | 0.034 | * |
| Subcortical volume | Amygdala | -0.031 | 0.018 | -1.741 | 0.082 | 0.141 |  |
| Subcortical volume | Pallidum | -0.025 | 0.023 | -1.105 | 0.269 | 0.394 |  |
| Subcortical volume | Hippocampus | -0.012 | 0.012 | -1.008 | 0.314 | 0.426 |  |
| Subcortical volume | Accumbens | 0.006 | 0.022 | 0.269 | 0.788 | 0.832 |  |
| Subcortical volume | Thalamus | -0.002 | 0.016 | -0.134 | 0.893 | 0.893 |  |
| **Section E:** Within-person, overweight vs healthy weight | | | | | | | |
| Cortical thickness | Medial OFC | -0.047 | 0.024 | -2.012 | 0.044 | 0.31 |  |
| Cortical thickness | Rostral ACC | -0.051 | 0.025 | -2.091 | 0.037 | 0.31 |  |
| Cortical thickness | Caudal ACC | -0.032 | 0.016 | -1.969 | 0.049 | 0.31 |  |
| Cortical thickness | Lateral OFC | -0.031 | 0.023 | -1.346 | 0.178 | 0.334 |  |
| Cortical thickness | Temporal pole | -0.052 | 0.031 | -1.707 | 0.088 | 0.334 |  |
| Cortical thickness | Superior frontal | -0.03 | 0.021 | -1.409 | 0.159 | 0.334 |  |
| Cortical thickness | Rostral mid-frontal | -0.03 | 0.022 | -1.397 | 0.162 | 0.334 |  |
| Cortical thickness | Fusiform | -0.028 | 0.021 | -1.333 | 0.183 | 0.334 |  |
| Cortical thickness | Lateral occipital | -0.025 | 0.019 | -1.301 | 0.193 | 0.334 |  |
| Cortical thickness | Insula | -0.019 | 0.027 | -0.684 | 0.494 | 0.626 |  |
| Cortical thickness | Middle temporal | -0.012 | 0.021 | -0.591 | 0.555 | 0.659 |  |
| Cortical thickness | Inferior temporal | -0.009 | 0.021 | -0.42 | 0.674 | 0.754 |  |
| Subcortical volume | Pallidum | -0.028 | 0.02 | -1.396 | 0.163 | 0.334 |  |
| Subcortical volume | Putamen | -0.018 | 0.011 | -1.693 | 0.09 | 0.334 |  |
| Subcortical volume | Amygdala | -0.016 | 0.016 | -1.019 | 0.308 | 0.45 |  |
| Subcortical volume | Thalamus | 0.015 | 0.015 | 1.039 | 0.299 | 0.45 |  |
| Subcortical volume | Hippocampus | -0.007 | 0.01 | -0.704 | 0.482 | 0.626 |  |
| Subcortical volume | Accumbens | -0.006 | 0.02 | -0.319 | 0.75 | 0.791 |  |
| Subcortical volume | Caudate | -0.001 | 0.009 | -0.094 | 0.925 | 0.925 |  |
| **Section F:** Within-person, underweight vs healthy weight | | | | | | | |
| Cortical thickness | Fusiform | -0.056 | 0.023 | -2.477 | 0.013 | 0.14 |  |
| Cortical thickness | Middle temporal | -0.04 | 0.022 | -1.789 | 0.074 | 0.357 |  |
| Cortical thickness | Lateral occipital | -0.037 | 0.021 | -1.78 | 0.075 | 0.357 |  |
| Cortical thickness | Medial OFC | 0.014 | 0.025 | 0.553 | 0.58 | 0.804 |  |
| Cortical thickness | Lateral OFC | 0.017 | 0.025 | 0.668 | 0.504 | 0.804 |  |
| Cortical thickness | Temporal pole | 0.026 | 0.033 | 0.784 | 0.433 | 0.804 |  |
| Cortical thickness | Rostral ACC | 0.012 | 0.027 | 0.461 | 0.644 | 0.804 |  |
| Cortical thickness | Superior frontal | 0.026 | 0.023 | 1.135 | 0.256 | 0.804 |  |
| Cortical thickness | Insula | -0.017 | 0.029 | -0.578 | 0.563 | 0.804 |  |
| Cortical thickness | Inferior temporal | -0.012 | 0.022 | -0.544 | 0.586 | 0.804 |  |
| Cortical thickness | Rostral mid-frontal | 0.006 | 0.023 | 0.27 | 0.787 | 0.88 |  |
| Cortical thickness | Caudal ACC | -0.003 | 0.018 | -0.155 | 0.877 | 0.926 |  |
| Subcortical volume | Putamen | -0.028 | 0.012 | -2.439 | 0.015 | 0.14 |  |
| Subcortical volume | Hippocampus | -0.016 | 0.011 | -1.473 | 0.141 | 0.535 |  |
| Subcortical volume | Accumbens | -0.01 | 0.021 | -0.484 | 0.629 | 0.804 |  |
| Subcortical volume | Caudate | -0.004 | 0.009 | -0.417 | 0.677 | 0.804 |  |
| Subcortical volume | Pallidum | 0.01 | 0.022 | 0.462 | 0.644 | 0.804 |  |
| Subcortical volume | Thalamus | 0.008 | 0.016 | 0.507 | 0.612 | 0.804 |  |
| Subcortical volume | Amygdala | 0 | 0.017 | 0.019 | 0.985 | 0.985 |  |
| Note. Interaction models were fitted across all participants with healthy weight as the reference group, including BMI-for-age z × weight status group interactions for both the between- and within-person components. Models were adjusted for age, age², sex, pubertal development, socioeconomic status, race and ethnicity, and intracranial volume (subcortical outcomes), with random intercepts for family and participant. Each coefficient is the difference in slope between the indicated group and the healthy weight group (Δβ). P values are from Wald tests (z = Δβ/SE); FDR correction was applied across 19 regions separately for each set of interaction terms, and significance markers are based on unrounded *q* values. BMI, body mass index; CDC, Centers for Disease Control and Prevention; FDR, false discovery rate; SE, standard error; *q, FDR-corrected P value*. **q* < .05; ***q* < .01; ****q* < .001. | | | | | | | |

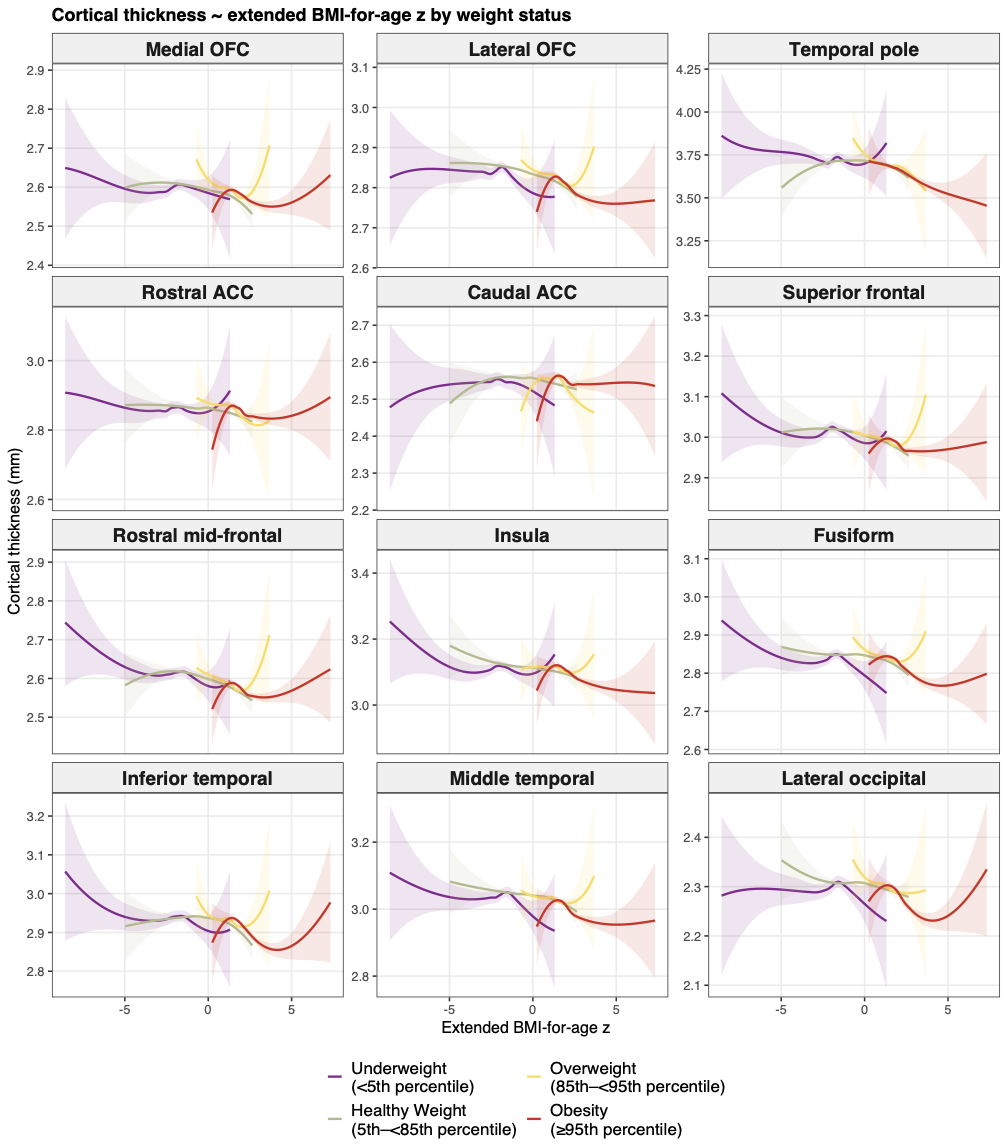

**eFigure 13.** LOESS associations between extended BMI-for-age z and cortical thickness by CDC weight status group. Locally weighted scatterplot smoothing (LOESS) curves of cortical thickness (mm; y-axis) against extended BMI-for-age z (x-axis) for each of the 12 a priori cortical regions, by CDC weight status group (color). Shaded bands indicate 95% confidence intervals. Observations from all 4 waves of the analytic sample are pooled. The curves illustrate the unadjusted within-group relationships underlying the stratified estimates in eTable 17, Section A.

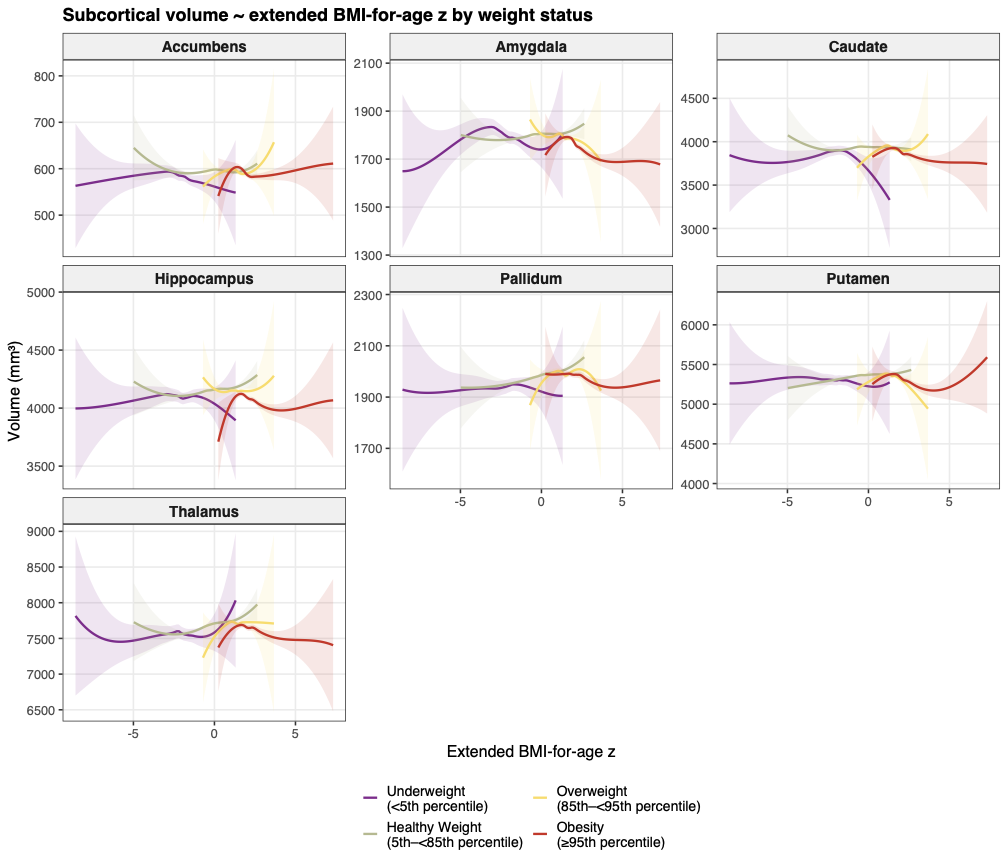

**eFigure 14.** LOESS associations between extended BMI-for-age z and subcortical volume by CDC weight status group. Locally weighted scatterplot smoothing (LOESS) curves of subcortical volume (mm³; y-axis) against extended BMI-for-age z (x-axis) for each of the 7 a priori subcortical regions, by CDC weight status group (color). Shaded bands indicate 95% confidence intervals. Observations from all 4 waves of the analytic sample are pooled. The positive association in the healthy weight group and the absent or reversed pattern in the obesity group are visible in the unadjusted data, consistent with the stratified estimates in eTable 17, Section A.

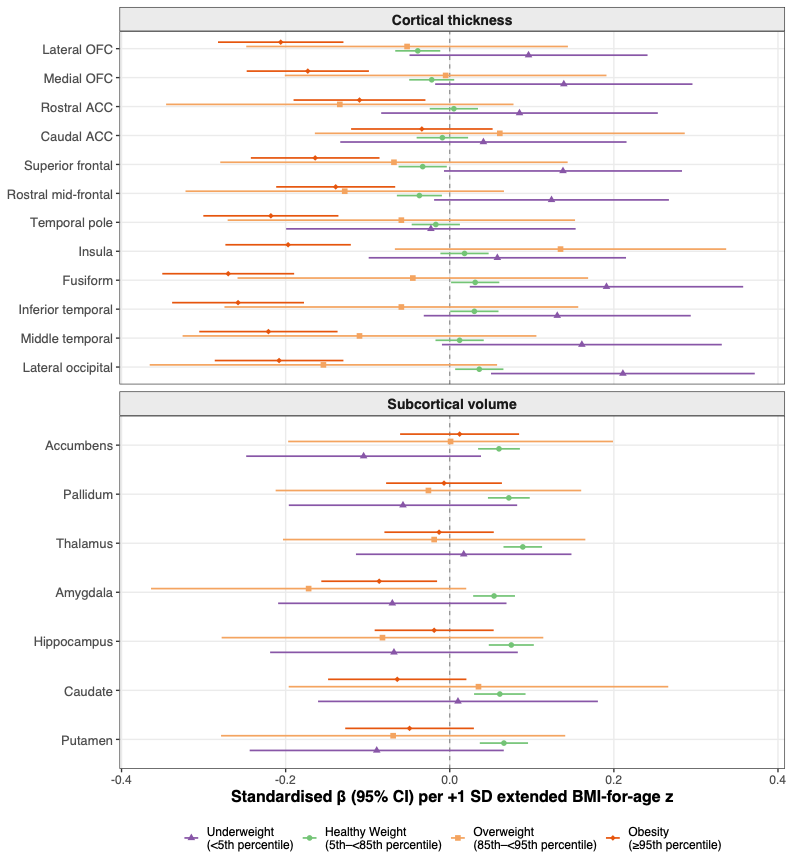
**eFigure 15.** Between-person BMI-for-age z associations with brain structure across all CDC weight status groups. Point-range forest plot of standardized between-person β coefficients (95% CIs) per 1-SD increase in extended BMI-for-age z for all 19 a priori regions (y-axis), with the 4 CDC weight status groups shown in separate colors and shapes (dodged). The upper panel shows cortical thickness and the lower panel subcortical volume; regions are ordered anatomically, and the dashed vertical line indicates β = 0. Models were adjusted for age, age², sex, pubertal development, socioeconomic status, race and ethnicity, and intracranial volume (subcortical outcomes). The figure complements the brain maps in Figure 3 of the main manuscript; full estimates are in eTable 17, Section A.

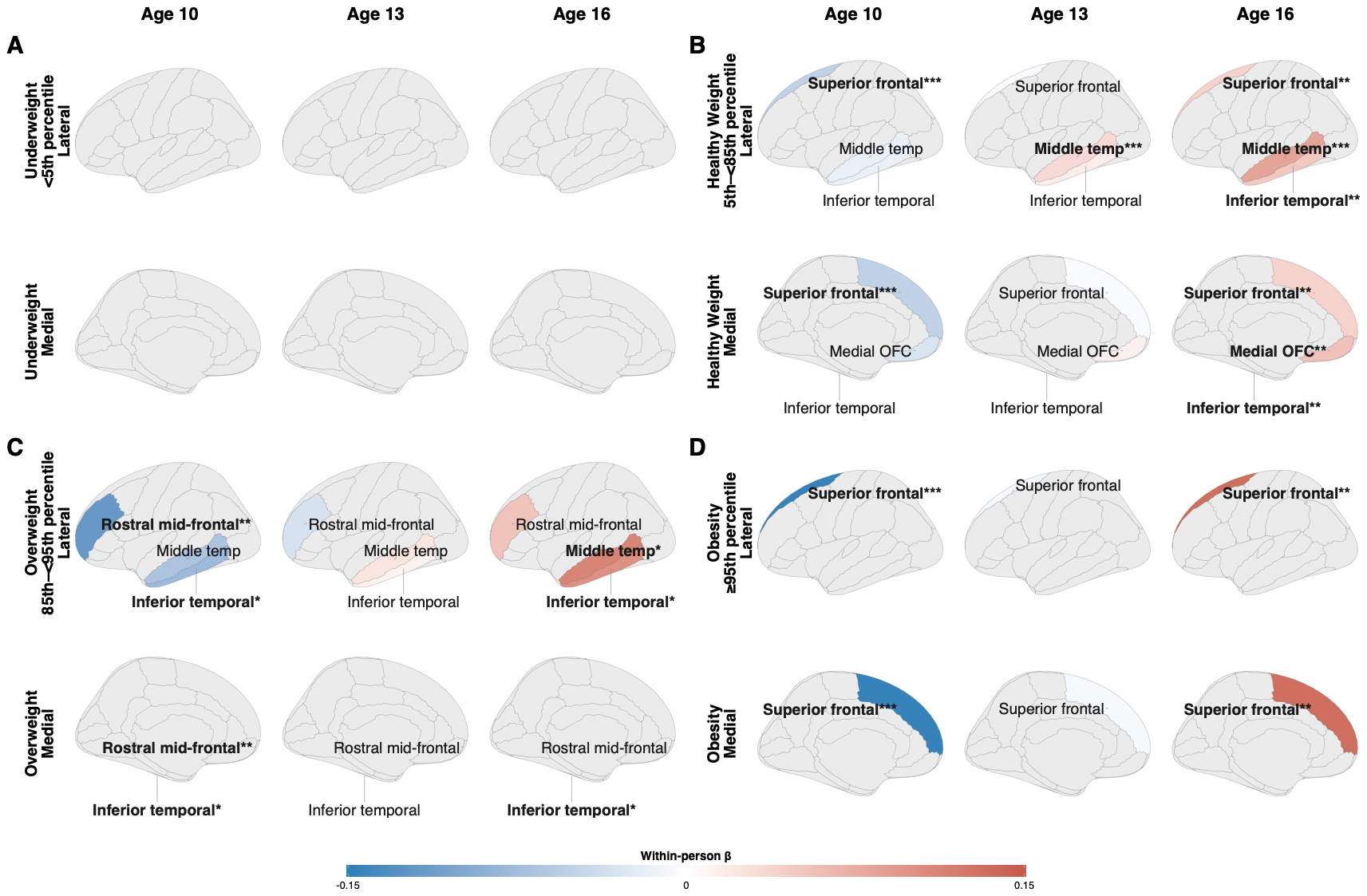

**eFigure 16.** Age moderation of within-person BMI-for-age z associations with cortical thickness across adolescence by CDC weight status group. Standardized within-person β per 1-SD increase in extended BMI-for-age z estimated at ages 10, 13, and 16 years (columns), shown in lateral and medial left-hemisphere views for each group: (A) Underweight (<5th percentile); (B) Healthy weight (5th to <85th percentile); (C) Overweight (85th to <95th percentile); (D) Obesity (≥95th percentile). Simple slopes were estimated by recentering age at each target age in models with a within-person BMI-for-age z × age interaction, adjusted for age², sex, pubertal development, socioeconomic status, and race and ethnicity. Regions are shown only where age moderation was significant (likelihood ratio test, *q* < .05; eTable 19). Blue indicates a negative within-person association (lower thickness at visits when a participant’s BMI-for-age z was above their own mean); red indicates a positive association. Bold labels denote FDR-significant simple slopes (*q* < .05) at that age (eTable 20). The color scale is shared across panels (−0.15 to 0.15). All 3 groups with significant age moderation (B, C, D) show within-person associations shifting from negative at age 10 to positive by age 16. At age 10, the strongest negative estimates were in superior frontal cortex in the healthy weight (β = −0.051) and obesity groups (β = −0.146) and in rostral middle frontal cortex in the overweight group (β = −0.118). No age moderation was observed in the underweight group (A).

| **eTable 19. Likelihood Ratio Tests for Age Moderation of Within-Person BMI-for-Age z Associations With Regional Brain Structure by CDC Weight Status Group.** | | | | | |
| --- | --- | --- | --- | --- | --- |
| group | region | LR_X_2 stat | P | q | sig |
| Healthy weight | Middle temporal | 18.539 | 0 | 0 | *** |
| Healthy weight | Superior frontal | 14.439 | 0 | 0.001 | ** |
| Healthy weight | Medial OFC | 9.855 | 0.002 | 0.011 | * |
| Healthy weight | Inferior temporal | 7.857 | 0.005 | 0.024 | * |
| Healthy weight | Lateral occipital | 4.891 | 0.027 | 0.103 |  |
| Healthy weight | Rostral mid-frontal | 4.182 | 0.041 | 0.118 |  |
| Healthy weight | Fusiform | 4.006 | 0.045 | 0.118 |  |
| Healthy weight | Caudal ACC | 3.251 | 0.071 | 0.136 |  |
| Healthy weight | Lateral OFC | 2.095 | 0.148 | 0.255 |  |
| Healthy weight | Rostral ACC | 1.062 | 0.303 | 0.449 |  |
| Healthy weight | Temporal pole | 0.204 | 0.652 | 0.729 |  |
| Healthy weight | Insula | 0.045 | 0.832 | 0.832 |  |
| Healthy weight | Thalamus | 3.858 | 0.05 | 0.118 |  |
| Healthy weight | Caudate | 3.277 | 0.07 | 0.136 |  |
| Healthy weight | Putamen | 1.044 | 0.307 | 0.449 |  |
| Healthy weight | Hippocampus | 0.89 | 0.345 | 0.469 |  |
| Healthy weight | Accumbens | 0.773 | 0.379 | 0.481 |  |
| Healthy weight | Pallidum | 0.552 | 0.457 | 0.543 |  |
| Healthy weight | Amygdala | 0.061 | 0.805 | 0.832 |  |
| Overweight | Inferior temporal | 7.992 | 0.005 | 0.045 | * |
| Overweight | Middle temporal | 7.962 | 0.005 | 0.045 | * |
| Overweight | Rostral mid-frontal | 7.144 | 0.008 | 0.048 | * |
| Overweight | Medial OFC | 3.263 | 0.071 | 0.224 |  |
| Overweight | Lateral occipital | 3.717 | 0.054 | 0.224 |  |
| Overweight | Fusiform | 2.969 | 0.085 | 0.23 |  |
| Overweight | Superior frontal | 2.508 | 0.113 | 0.269 |  |
| Overweight | Lateral OFC | 0.95 | 0.33 | 0.569 |  |
| Overweight | Temporal pole | 0.81 | 0.368 | 0.569 |  |
| Overweight | Rostral ACC | 0.672 | 0.412 | 0.569 |  |
| Overweight | Caudal ACC | 0.572 | 0.45 | 0.569 |  |
| Overweight | Insula | 0.497 | 0.481 | 0.571 |  |
| Overweight | Hippocampus | 3.378 | 0.066 | 0.224 |  |
| Overweight | Caudate | 1.688 | 0.194 | 0.409 |  |
| Overweight | Amygdala | 1.244 | 0.265 | 0.503 |  |
| Overweight | Thalamus | 0.625 | 0.429 | 0.569 |  |
| Overweight | Putamen | 0.244 | 0.621 | 0.694 |  |
| Overweight | Accumbens | 0.131 | 0.718 | 0.758 |  |
| Overweight | Pallidum | 0 | 0.989 | 0.989 |  |
| Obese | Superior frontal | 13.65 | 0 | 0.004 | ** |
| Obese | Rostral mid-frontal | 7.484 | 0.006 | 0.059 |  |
| Obese | Fusiform | 3.766 | 0.052 | 0.331 |  |
| Obese | Caudal ACC | 1.808 | 0.179 | 0.583 |  |
| Obese | Lateral OFC | 1.024 | 0.312 | 0.707 |  |
| Obese | Temporal pole | 0.929 | 0.335 | 0.707 |  |
| Obese | Lateral occipital | 1.036 | 0.309 | 0.707 |  |
| Obese | Insula | 0.515 | 0.473 | 0.749 |  |
| Obese | Middle temporal | 0.623 | 0.43 | 0.749 |  |
| Obese | Medial OFC | 0.102 | 0.75 | 0.924 |  |
| Obese | Rostral ACC | 0.025 | 0.875 | 0.924 |  |
| Obese | Inferior temporal | 0.066 | 0.797 | 0.924 |  |
| Obese | Hippocampus | 2.61 | 0.106 | 0.505 |  |
| Obese | Pallidum | 1.765 | 0.184 | 0.583 |  |
| Obese | Thalamus | 0.59 | 0.442 | 0.749 |  |
| Obese | Amygdala | 0.227 | 0.633 | 0.881 |  |
| Obese | Caudate | 0.207 | 0.649 | 0.881 |  |
| Obese | Accumbens | 0.027 | 0.87 | 0.924 |  |
| Obese | Putamen | 0.002 | 0.964 | 0.964 |  |
| Underweight | Lateral occipital | 8.87 | 0.003 | 0.055 |  |
| Underweight | Temporal pole | 4.9 | 0.027 | 0.102 |  |
| Underweight | Rostral mid-frontal | 5.127 | 0.024 | 0.102 |  |
| Underweight | Fusiform | 5.502 | 0.019 | 0.102 |  |
| Underweight | Middle temporal | 4.895 | 0.027 | 0.102 |  |
| Underweight | Inferior temporal | 3.71 | 0.054 | 0.171 |  |
| Underweight | Insula | 2.516 | 0.113 | 0.306 |  |
| Underweight | Superior frontal | 2.136 | 0.144 | 0.342 |  |
| Underweight | Rostral ACC | 1.868 | 0.172 | 0.362 |  |
| Underweight | Medial OFC | 0.026 | 0.873 | 0.951 |  |
| Underweight | Lateral OFC | 0.063 | 0.801 | 0.951 |  |
| Underweight | Caudal ACC | 0.058 | 0.81 | 0.951 |  |
| Underweight | Caudate | 0.63 | 0.427 | 0.812 |  |
| Underweight | Pallidum | 0.475 | 0.491 | 0.847 |  |
| Underweight | Accumbens | 0.006 | 0.941 | 0.951 |  |
| Underweight | Amygdala | 0.007 | 0.935 | 0.951 |  |
| Underweight | Hippocampus | 0.111 | 0.739 | 0.951 |  |
| Underweight | Putamen | 0.009 | 0.926 | 0.951 |  |
| Underweight | Thalamus | 0.004 | 0.951 | 0.951 |  |
| Note. Age moderation was tested within each CDC weight status group by comparing models with and without a within-person BMI-for-age z × age interaction using likelihood ratio tests (1 df). Both models were fitted by maximum likelihood and adjusted for age², sex, pubertal development, socioeconomic status, race and ethnicity, and intracranial volume (subcortical outcomes), with random intercepts for family and participant. LR χ² is the likelihood ratio statistic. FDR correction was applied within each group across 19 regions, and significance markers are based on unrounded *q* values. BMI, body mass index; CDC, Centers for Disease Control and Prevention; FDR, false discovery rate; LR, likelihood ratio; *q, FDR-corrected P value*. **q* < .05; ***q* < .01; ****q* < .001. | | | | | |

| **eTable 20. Simple Slopes of Within-Person BMI-for-Age z Associations With Cortical Thickness at Ages 10, 13, and 16 Years for Regions Showing Significant Age Moderation, by CDC Weight Status Group.** | | | | | | | | |
| --- | --- | --- | --- | --- | --- | --- | --- | --- |
| group | region | age | β | SE | z | p | q | sig |
| Healthy weight | Inferior temporal | 10 | -0.017 | 0.013 | -1.237 | 0.216 | 0.216 |  |
| Healthy weight | Inferior temporal | 13 | 0.016 | 0.008 | 2.167 | 0.03 | 0.061 |  |
| Healthy weight | Inferior temporal | 16 | 0.049 | 0.015 | 3.387 | 0.001 | 0.001 | ** |
| Healthy weight | Medial OFC | 10 | -0.029 | 0.015 | -1.873 | 0.061 | 0.122 |  |
| Healthy weight | Medial OFC | 13 | 0.013 | 0.009 | 1.496 | 0.135 | 0.18 |  |
| Healthy weight | Medial OFC | 16 | 0.055 | 0.017 | 3.306 | 0.001 | 0.001 | ** |
| Healthy weight | Middle temporal | 10 | -0.017 | 0.013 | -1.297 | 0.194 | 0.216 |  |
| Healthy weight | Middle temporal | 13 | 0.033 | 0.008 | 4.39 | 0 | 0 | *** |
| Healthy weight | Middle temporal | 16 | 0.083 | 0.014 | 5.755 | 0 | 0 | *** |
| Healthy weight | Superior frontal | 10 | -0.051 | 0.014 | -3.746 | 0 | 0.001 | *** |
| Healthy weight | Superior frontal | 13 | -0.006 | 0.008 | -0.722 | 0.47 | 0.47 |  |
| Healthy weight | Superior frontal | 16 | 0.04 | 0.015 | 2.684 | 0.007 | 0.007 | ** |
| Overweight | Inferior temporal | 10 | -0.078 | 0.036 | -2.142 | 0.032 | 0.048 | * |
| Overweight | Inferior temporal | 13 | 0.012 | 0.021 | 0.558 | 0.577 | 0.577 |  |
| Overweight | Inferior temporal | 16 | 0.101 | 0.039 | 2.57 | 0.01 | 0.015 | * |
| Overweight | Middle temporal | 10 | -0.066 | 0.036 | -1.823 | 0.068 | 0.068 |  |
| Overweight | Middle temporal | 13 | 0.023 | 0.021 | 1.107 | 0.268 | 0.402 |  |
| Overweight | Middle temporal | 16 | 0.112 | 0.039 | 2.854 | 0.004 | 0.013 | * |
| Overweight | Rostral mid-frontal | 10 | -0.118 | 0.037 | -3.181 | 0.001 | 0.004 | ** |
| Overweight | Rostral mid-frontal | 13 | -0.032 | 0.021 | -1.499 | 0.134 | 0.402 |  |
| Overweight | Rostral mid-frontal | 16 | 0.054 | 0.04 | 1.34 | 0.18 | 0.18 |  |
| Obese | Superior frontal | 10 | -0.146 | 0.042 | -3.429 | 0.001 | 0.001 | *** |
| Obese | Superior frontal | 13 | -0.007 | 0.024 | -0.295 | 0.768 | 0.768 |  |
| Obese | Superior frontal | 16 | 0.132 | 0.046 | 2.837 | 0.005 | 0.005 | ** |
| Note. Simple slopes are within-person BMI-for-age z associations at ages 10, 13, and 16 years, estimated from the age moderation models by recentering age at each target age. Only regions with significant age moderation in eTable 19 (*q* < .05) are shown; all were cortical. β is the standardized within-person coefficient at the indicated age. P values are from Wald tests (z = β/SE); FDR correction was applied within each group and age across the regions shown, and significance markers are based on unrounded *q* values. BMI, body mass index; CDC, Centers for Disease Control and Prevention; FDR, false discovery rate; SE, standard error; *q, FDR-corrected P value*. **q* < .05; ***q* < .01; ****q* < .001. | | | | | | | | |

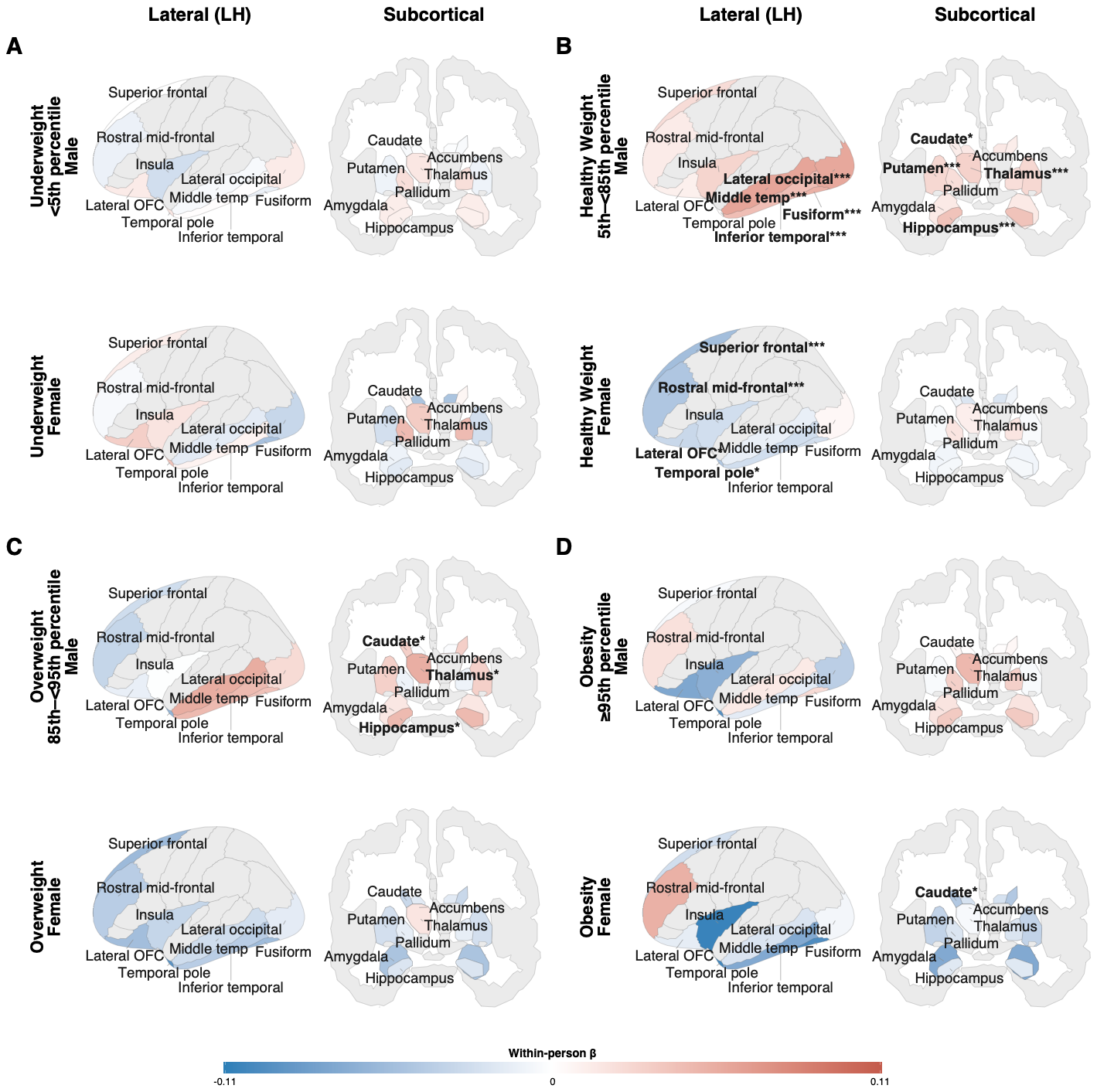

**eFigure 17.** Sex moderation of within-person BMI-for-age z associations with brain structure by CDC weight status group. Standardized within-person β per 1-SD increase in extended BMI-for-age z shown separately for male and female participants within each group, mapped to the left-hemisphere lateral cortical surface and a subcortical coronal section: (A) Underweight (<5th percentile); (B) Healthy weight (5th to <85th percentile); (C) Overweight (85th to <95th percentile); (D) Obesity (≥95th percentile). Sex-specific slopes were estimated by refitting models with a within-person BMI-for-age z × sex interaction with each sex as the reference in turn, adjusted for age, age², pubertal development, socioeconomic status, race and ethnicity, and intracranial volume (subcortical outcomes) (eTables 21 and 22). Blue indicates a negative within-person association; red indicates a positive association. Bold labels denote FDR-significant sex-specific associations (*q* < .05). The color scale is shared across panels (−0.11 to 0.11). In healthy weight youth, male participants show positive within-person associations in temporal cortex and subcortical structures, whereas female participants show negative associations in frontal cortex; within-person × sex interactions are significant in 13 of 19 regions (B). In the overweight group, sex moderation is present in 3 subcortical regions, with positive slopes in male participants (C). In the obesity and underweight groups, no within-person × sex interaction survived correction (A, D), although the female-specific caudate slope was significant in the obesity group.

| **eTable 21. Within-Person × Sex Interaction Terms for BMI-for-Age z Associations With Brain Structure by CDC Weight Status Group.** | | | | | | | |
| --- | --- | --- | --- | --- | --- | --- | --- |
| group | region | β | SE | z | p | q | sig |
| Healthy weight | Superior frontal | -0.077 | 0.015 | -4.98 | 0 | 0 | *** |
| Healthy weight | Rostral mid-frontal | -0.06 | 0.016 | -3.807 | 0 | 0 | *** |
| Healthy weight | Fusiform | -0.079 | 0.015 | -5.218 | 0 | 0 | *** |
| Healthy weight | Inferior temporal | -0.071 | 0.015 | -4.714 | 0 | 0 | *** |
| Healthy weight | Middle temporal | -0.073 | 0.015 | -4.875 | 0 | 0 | *** |
| Healthy weight | Lateral occipital | -0.053 | 0.014 | -3.703 | 0 | 0.001 | *** |
| Healthy weight | Caudal ACC | -0.038 | 0.012 | -3.114 | 0.002 | 0.004 | ** |
| Healthy weight | Insula | -0.055 | 0.02 | -2.671 | 0.008 | 0.013 | * |
| Healthy weight | Lateral OFC | -0.043 | 0.017 | -2.546 | 0.011 | 0.017 | * |
| Healthy weight | Temporal pole | -0.046 | 0.022 | -2.067 | 0.039 | 0.053 |  |
| Healthy weight | Medial OFC | -0.021 | 0.017 | -1.195 | 0.232 | 0.259 |  |
| Healthy weight | Rostral ACC | -0.003 | 0.019 | -0.163 | 0.871 | 0.871 |  |
| Healthy weight | Hippocampus | -0.046 | 0.008 | -5.997 | 0 | 0 | *** |
| Healthy weight | Putamen | -0.03 | 0.008 | -3.701 | 0 | 0.001 | *** |
| Healthy weight | Caudate | -0.02 | 0.006 | -3.164 | 0.002 | 0.003 | ** |
| Healthy weight | Accumbens | -0.036 | 0.015 | -2.436 | 0.015 | 0.022 | * |
| Healthy weight | Amygdala | -0.021 | 0.012 | -1.825 | 0.068 | 0.086 |  |
| Healthy weight | Thalamus | -0.019 | 0.011 | -1.724 | 0.085 | 0.101 |  |
| Healthy weight | Pallidum | -0.005 | 0.015 | -0.321 | 0.748 | 0.79 |  |
| Overweight | Fusiform | -0.098 | 0.04 | -2.445 | 0.014 | 0.069 |  |
| Overweight | Inferior temporal | -0.087 | 0.041 | -2.133 | 0.033 | 0.104 |  |
| Overweight | Middle temporal | -0.083 | 0.041 | -2.051 | 0.04 | 0.109 |  |
| Overweight | Medial OFC | -0.06 | 0.045 | -1.357 | 0.175 | 0.34 |  |
| Overweight | Rostral ACC | -0.062 | 0.046 | -1.356 | 0.175 | 0.34 |  |
| Overweight | Caudal ACC | -0.032 | 0.032 | -1.004 | 0.315 | 0.499 |  |
| Overweight | Lateral occipital | -0.039 | 0.037 | -1.052 | 0.293 | 0.499 |  |
| Overweight | Lateral OFC | -0.041 | 0.044 | -0.93 | 0.353 | 0.515 |  |
| Overweight | Superior frontal | -0.029 | 0.04 | -0.714 | 0.475 | 0.645 |  |
| Overweight | Insula | -0.033 | 0.05 | -0.65 | 0.516 | 0.653 |  |
| Overweight | Temporal pole | -0.03 | 0.058 | -0.519 | 0.604 | 0.717 |  |
| Overweight | Rostral mid-frontal | -0.007 | 0.042 | -0.173 | 0.863 | 0.9 |  |
| Overweight | Caudate | -0.064 | 0.016 | -4.083 | 0 | 0.001 | *** |
| Overweight | Hippocampus | -0.067 | 0.019 | -3.585 | 0 | 0.003 | ** |
| Overweight | Putamen | -0.057 | 0.02 | -2.854 | 0.004 | 0.027 | * |
| Overweight | Amygdala | -0.066 | 0.03 | -2.16 | 0.031 | 0.104 |  |
| Overweight | Thalamus | -0.037 | 0.028 | -1.344 | 0.179 | 0.34 |  |
| Overweight | Accumbens | -0.015 | 0.037 | -0.395 | 0.693 | 0.774 |  |
| Overweight | Pallidum | -0.005 | 0.037 | -0.126 | 0.9 | 0.9 |  |
| Obese | Fusiform | -0.122 | 0.05 | -2.459 | 0.014 | 0.076 |  |
| Obese | Rostral ACC | -0.086 | 0.051 | -1.679 | 0.093 | 0.253 |  |
| Obese | Caudal ACC | -0.059 | 0.034 | -1.705 | 0.088 | 0.253 |  |
| Obese | Lateral OFC | 0.059 | 0.053 | 1.099 | 0.272 | 0.47 |  |
| Obese | Inferior temporal | -0.056 | 0.049 | -1.157 | 0.247 | 0.47 |  |
| Obese | Medial OFC | -0.031 | 0.052 | -0.588 | 0.556 | 0.632 |  |
| Obese | Rostral mid-frontal | 0.032 | 0.049 | 0.652 | 0.514 | 0.632 |  |
| Obese | Insula | -0.039 | 0.056 | -0.688 | 0.492 | 0.632 |  |
| Obese | Middle temporal | -0.036 | 0.047 | -0.759 | 0.448 | 0.632 |  |
| Obese | Lateral occipital | 0.034 | 0.043 | 0.789 | 0.43 | 0.632 |  |
| Obese | Superior frontal | -0.022 | 0.046 | -0.473 | 0.636 | 0.672 |  |
| Obese | Temporal pole | -0.007 | 0.072 | -0.1 | 0.92 | 0.92 |  |
| Obese | Amygdala | -0.09 | 0.034 | -2.658 | 0.008 | 0.075 |  |
| Obese | Caudate | -0.05 | 0.018 | -2.7 | 0.007 | 0.075 |  |
| Obese | Hippocampus | -0.053 | 0.022 | -2.407 | 0.016 | 0.076 |  |
| Obese | Thalamus | -0.053 | 0.031 | -1.717 | 0.086 | 0.253 |  |
| Obese | Putamen | -0.032 | 0.024 | -1.351 | 0.177 | 0.42 |  |
| Obese | Pallidum | -0.052 | 0.045 | -1.164 | 0.245 | 0.47 |  |
| Obese | Accumbens | -0.024 | 0.042 | -0.575 | 0.565 | 0.632 |  |
| Underweight | Medial OFC | -0.02 | 0.048 | -0.424 | 0.671 | 0.889 |  |
| Underweight | Lateral OFC | 0.019 | 0.047 | 0.4 | 0.69 | 0.889 |  |
| Underweight | Temporal pole | 0.024 | 0.061 | 0.39 | 0.696 | 0.889 |  |
| Underweight | Rostral ACC | 0.048 | 0.05 | 0.953 | 0.34 | 0.889 |  |
| Underweight | Caudal ACC | 0.012 | 0.033 | 0.35 | 0.726 | 0.889 |  |
| Underweight | Superior frontal | 0.013 | 0.041 | 0.311 | 0.756 | 0.889 |  |
| Underweight | Rostral mid-frontal | 0.005 | 0.039 | 0.14 | 0.889 | 0.889 |  |
| Underweight | Insula | 0.045 | 0.057 | 0.794 | 0.427 | 0.889 |  |
| Underweight | Fusiform | -0.039 | 0.043 | -0.908 | 0.364 | 0.889 |  |
| Underweight | Inferior temporal | 0.007 | 0.041 | 0.181 | 0.857 | 0.889 |  |
| Underweight | Middle temporal | -0.01 | 0.042 | -0.239 | 0.811 | 0.889 |  |
| Underweight | Lateral occipital | -0.046 | 0.039 | -1.179 | 0.238 | 0.889 |  |
| Underweight | Accumbens | -0.047 | 0.041 | -1.142 | 0.253 | 0.889 |  |
| Underweight | Amygdala | -0.022 | 0.035 | -0.631 | 0.528 | 0.889 |  |
| Underweight | Caudate | 0.01 | 0.017 | 0.57 | 0.569 | 0.889 |  |
| Underweight | Hippocampus | -0.027 | 0.019 | -1.431 | 0.152 | 0.889 |  |
| Underweight | Pallidum | 0.024 | 0.043 | 0.549 | 0.583 | 0.889 |  |
| Underweight | Putamen | -0.015 | 0.023 | -0.667 | 0.505 | 0.889 |  |
| Underweight | Thalamus | 0.018 | 0.029 | 0.62 | 0.535 | 0.889 |  |
| Note. Within-person BMI-for-age z × sex interactions (male participants as reference) were added to the primary within-between model within each CDC weight status group. Models were adjusted for age, age², pubertal development, socioeconomic status, race and ethnicity, and intracranial volume (subcortical outcomes), with random intercepts for family and participant. Negative coefficients indicate a more negative within-person slope in female than in male participants. P values are from Wald tests (z = β/SE); FDR correction was applied within each group across 19 regions, and significance markers are based on unrounded *q* values. BMI, body mass index; CDC, Centers for Disease Control and Prevention; FDR, false discovery rate; SE, standard error; *q, FDR-corrected P value*. **q* < .05; ***q* < .01; ****q* < .001. | | | | | | | |

| **eTable 22. Sex-Specific Within-Person BMI-for-Age z Associations With Brain Structure by CDC Weight Status Group.** | | | | | | |
| --- | --- | --- | --- | --- | --- | --- |
| group | region | sex | β | SE | q | sig |
| Healthy weight | Superior frontal | Female | -0.054 | 0.012 | 0 | *** |
| Healthy weight | Rostral mid-frontal | Female | -0.047 | 0.012 | 0.001 | *** |
| Healthy weight | Temporal pole | Female | -0.045 | 0.017 | 0.029 | * |
| Healthy weight | Lateral OFC | Female | -0.032 | 0.013 | 0.038 | * |
| Healthy weight | Inferior temporal | Female | -0.027 | 0.012 | 0.051 |  |
| Healthy weight | Fusiform | Female | -0.026 | 0.012 | 0.057 |  |
| Healthy weight | Insula | Female | -0.027 | 0.016 | 0.149 |  |
| Healthy weight | Caudal ACC | Female | -0.015 | 0.009 | 0.182 |  |
| Healthy weight | Middle temporal | Female | -0.013 | 0.012 | 0.343 |  |
| Healthy weight | Rostral ACC | Female | 0.008 | 0.014 | 0.632 |  |
| Healthy weight | Lateral occipital | Female | 0.005 | 0.011 | 0.725 |  |
| Healthy weight | Medial OFC | Female | -0.002 | 0.014 | 0.925 |  |
| Healthy weight | Fusiform | Male | 0.052 | 0.01 | 0 | *** |
| Healthy weight | Inferior temporal | Male | 0.043 | 0.01 | 0 | *** |
| Healthy weight | Middle temporal | Male | 0.06 | 0.01 | 0 | *** |
| Healthy weight | Lateral occipital | Male | 0.058 | 0.009 | 0 | *** |
| Healthy weight | Caudal ACC | Male | 0.023 | 0.008 | 0.013 | * |
| Healthy weight | Superior frontal | Male | 0.023 | 0.01 | 0.052 |  |
| Healthy weight | Insula | Male | 0.028 | 0.013 | 0.079 |  |
| Healthy weight | Medial OFC | Male | 0.019 | 0.011 | 0.149 |  |
| Healthy weight | Rostral mid-frontal | Male | 0.013 | 0.01 | 0.268 |  |
| Healthy weight | Lateral OFC | Male | 0.01 | 0.011 | 0.412 |  |
| Healthy weight | Rostral ACC | Male | 0.011 | 0.012 | 0.432 |  |
| Healthy weight | Temporal pole | Male | 0.001 | 0.014 | 0.94 |  |
| Healthy weight | Accumbens | Female | -0.021 | 0.011 | 0.123 |  |
| Healthy weight | Caudate | Female | -0.008 | 0.005 | 0.149 |  |
| Healthy weight | Pallidum | Female | 0.017 | 0.012 | 0.224 |  |
| Healthy weight | Thalamus | Female | 0.011 | 0.008 | 0.268 |  |
| Healthy weight | Hippocampus | Female | -0.006 | 0.006 | 0.376 |  |
| Healthy weight | Amygdala | Female | -0.008 | 0.009 | 0.448 |  |
| Healthy weight | Putamen | Female | -0.004 | 0.006 | 0.55 |  |
| Healthy weight | Hippocampus | Male | 0.04 | 0.005 | 0 | *** |
| Healthy weight | Putamen | Male | 0.026 | 0.005 | 0 | *** |
| Healthy weight | Thalamus | Male | 0.03 | 0.007 | 0 | *** |
| Healthy weight | Caudate | Male | 0.012 | 0.004 | 0.015 | * |
| Healthy weight | Pallidum | Male | 0.022 | 0.01 | 0.062 |  |
| Healthy weight | Amygdala | Male | 0.014 | 0.008 | 0.14 |  |
| Healthy weight | Accumbens | Male | 0.015 | 0.01 | 0.193 |  |
| Overweight | Medial OFC | Female | -0.074 | 0.033 | 0.141 |  |
| Overweight | Temporal pole | Female | -0.092 | 0.043 | 0.141 |  |
| Overweight | Rostral ACC | Female | -0.079 | 0.034 | 0.141 |  |
| Overweight | Caudal ACC | Female | -0.045 | 0.024 | 0.178 |  |
| Overweight | Superior frontal | Female | -0.057 | 0.03 | 0.178 |  |
| Overweight | Fusiform | Female | -0.056 | 0.03 | 0.178 |  |
| Overweight | Lateral OFC | Female | -0.054 | 0.033 | 0.235 |  |
| Overweight | Inferior temporal | Female | -0.041 | 0.03 | 0.335 |  |
| Overweight | Rostral mid-frontal | Female | -0.041 | 0.032 | 0.352 |  |
| Overweight | Insula | Female | -0.034 | 0.037 | 0.533 |  |
| Overweight | Middle temporal | Female | -0.028 | 0.03 | 0.533 |  |
| Overweight | Lateral occipital | Female | -0.016 | 0.028 | 0.696 |  |
| Overweight | Middle temporal | Male | 0.055 | 0.027 | 0.168 |  |
| Overweight | Inferior temporal | Male | 0.046 | 0.028 | 0.235 |  |
| Overweight | Temporal pole | Male | -0.062 | 0.039 | 0.255 |  |
| Overweight | Fusiform | Male | 0.041 | 0.027 | 0.265 |  |
| Overweight | Rostral mid-frontal | Male | -0.034 | 0.029 | 0.413 |  |
| Overweight | Superior frontal | Male | -0.028 | 0.027 | 0.511 |  |
| Overweight | Lateral occipital | Male | 0.024 | 0.025 | 0.533 |  |
| Overweight | Caudal ACC | Male | -0.013 | 0.022 | 0.687 |  |
| Overweight | Rostral ACC | Male | -0.017 | 0.031 | 0.696 |  |
| Overweight | Medial OFC | Male | -0.013 | 0.03 | 0.722 |  |
| Overweight | Lateral OFC | Male | -0.013 | 0.03 | 0.722 |  |
| Overweight | Insula | Male | -0.001 | 0.034 | 0.968 |  |
| Overweight | Caudate | Female | -0.032 | 0.012 | 0.055 |  |
| Overweight | Amygdala | Female | -0.049 | 0.023 | 0.141 |  |
| Overweight | Putamen | Female | -0.026 | 0.015 | 0.211 |  |
| Overweight | Hippocampus | Female | -0.021 | 0.014 | 0.265 |  |
| Overweight | Thalamus | Female | 0.017 | 0.021 | 0.568 |  |
| Overweight | Accumbens | Female | -0.017 | 0.028 | 0.687 |  |
| Overweight | Pallidum | Female | -0.014 | 0.028 | 0.699 |  |
| Overweight | Hippocampus | Male | 0.046 | 0.013 | 0.011 | * |
| Overweight | Caudate | Male | 0.032 | 0.011 | 0.046 | * |
| Overweight | Thalamus | Male | 0.054 | 0.019 | 0.046 | * |
| Overweight | Putamen | Male | 0.031 | 0.014 | 0.141 |  |
| Overweight | Amygdala | Male | 0.016 | 0.021 | 0.579 |  |
| Overweight | Pallidum | Male | -0.01 | 0.025 | 0.741 |  |
| Overweight | Accumbens | Male | -0.003 | 0.025 | 0.939 |  |
| Obese | Fusiform | Female | -0.098 | 0.036 | 0.083 |  |
| Obese | Insula | Female | -0.106 | 0.041 | 0.087 |  |
| Obese | Rostral ACC | Female | -0.087 | 0.037 | 0.143 |  |
| Obese | Inferior temporal | Female | -0.072 | 0.035 | 0.169 |  |
| Obese | Temporal pole | Female | -0.102 | 0.052 | 0.179 |  |
| Obese | Caudal ACC | Female | -0.044 | 0.025 | 0.228 |  |
| Obese | Rostral mid-frontal | Female | 0.052 | 0.036 | 0.37 |  |
| Obese | Superior frontal | Female | -0.027 | 0.034 | 0.791 |  |
| Obese | Middle temporal | Female | -0.022 | 0.034 | 0.793 |  |
| Obese | Lateral OFC | Female | -0.015 | 0.039 | 0.866 |  |
| Obese | Lateral occipital | Female | -0.007 | 0.031 | 0.929 |  |
| Obese | Medial OFC | Female | 0.004 | 0.038 | 0.962 |  |
| Obese | Lateral OFC | Male | -0.074 | 0.038 | 0.179 |  |
| Obese | Temporal pole | Male | -0.094 | 0.051 | 0.201 |  |
| Obese | Insula | Male | -0.067 | 0.04 | 0.245 |  |
| Obese | Lateral occipital | Male | -0.04 | 0.03 | 0.436 |  |
| Obese | Medial OFC | Male | 0.035 | 0.037 | 0.692 |  |
| Obese | Fusiform | Male | 0.024 | 0.035 | 0.793 |  |
| Obese | Caudal ACC | Male | 0.015 | 0.024 | 0.801 |  |
| Obese | Rostral mid-frontal | Male | 0.02 | 0.035 | 0.803 |  |
| Obese | Inferior temporal | Male | -0.016 | 0.034 | 0.866 |  |
| Obese | Middle temporal | Male | 0.014 | 0.033 | 0.866 |  |
| Obese | Superior frontal | Male | -0.005 | 0.033 | 0.957 |  |
| Obese | Rostral ACC | Male | -0.001 | 0.036 | 0.979 |  |
| Obese | Caudate | Female | -0.045 | 0.013 | 0.03 | * |
| Obese | Amygdala | Female | -0.072 | 0.025 | 0.063 |  |
| Obese | Putamen | Female | -0.036 | 0.017 | 0.164 |  |
| Obese | Hippocampus | Female | -0.018 | 0.016 | 0.592 |  |
| Obese | Accumbens | Female | -0.024 | 0.031 | 0.791 |  |
| Obese | Pallidum | Female | -0.021 | 0.033 | 0.793 |  |
| Obese | Thalamus | Female | -0.005 | 0.022 | 0.929 |  |
| Obese | Hippocampus | Male | 0.035 | 0.016 | 0.155 |  |
| Obese | Thalamus | Male | 0.047 | 0.022 | 0.163 |  |
| Obese | Pallidum | Male | 0.031 | 0.032 | 0.686 |  |
| Obese | Amygdala | Male | 0.018 | 0.024 | 0.791 |  |
| Obese | Caudate | Male | 0.005 | 0.013 | 0.866 |  |
| Obese | Putamen | Male | -0.004 | 0.017 | 0.929 |  |
| Obese | Accumbens | Male | 0.001 | 0.03 | 0.979 |  |
| Underweight | Medial OFC | Female | 0.017 | 0.038 | 0.915 |  |
| Underweight | Lateral OFC | Female | 0.032 | 0.038 | 0.915 |  |
| Underweight | Temporal pole | Female | 0.04 | 0.049 | 0.915 |  |
| Underweight | Rostral ACC | Female | 0.05 | 0.04 | 0.915 |  |
| Underweight | Caudal ACC | Female | 0.01 | 0.027 | 0.915 |  |
| Underweight | Superior frontal | Female | 0.012 | 0.033 | 0.915 |  |
| Underweight | Insula | Female | 0.018 | 0.046 | 0.915 |  |
| Underweight | Fusiform | Female | -0.051 | 0.034 | 0.915 |  |
| Underweight | Middle temporal | Female | -0.016 | 0.034 | 0.915 |  |
| Underweight | Lateral occipital | Female | -0.033 | 0.031 | 0.915 |  |
| Underweight | Rostral mid-frontal | Female | -0.005 | 0.031 | 0.988 |  |
| Underweight | Inferior temporal | Female | 0.008 | 0.033 | 0.988 |  |
| Underweight | Medial OFC | Male | 0.037 | 0.03 | 0.915 |  |
| Underweight | Lateral OFC | Male | 0.013 | 0.029 | 0.915 |  |
| Underweight | Temporal pole | Male | 0.016 | 0.038 | 0.915 |  |
| Underweight | Rostral mid-frontal | Male | -0.011 | 0.024 | 0.915 |  |
| Underweight | Insula | Male | -0.027 | 0.035 | 0.915 |  |
| Underweight | Fusiform | Male | -0.012 | 0.026 | 0.915 |  |
| Underweight | Lateral occipital | Male | 0.013 | 0.024 | 0.915 |  |
| Underweight | Rostral ACC | Male | 0.003 | 0.031 | 0.988 |  |
| Underweight | Caudal ACC | Male | -0.002 | 0.02 | 0.988 |  |
| Underweight | Superior frontal | Male | 0 | 0.025 | 0.988 |  |
| Underweight | Inferior temporal | Male | 0.001 | 0.025 | 0.988 |  |
| Underweight | Middle temporal | Male | -0.005 | 0.026 | 0.988 |  |
| Underweight | Accumbens | Female | -0.049 | 0.033 | 0.915 |  |
| Underweight | Amygdala | Female | -0.01 | 0.028 | 0.915 |  |
| Underweight | Caudate | Female | 0.006 | 0.013 | 0.915 |  |
| Underweight | Hippocampus | Female | -0.017 | 0.015 | 0.915 |  |
| Underweight | Pallidum | Female | 0.045 | 0.035 | 0.915 |  |
| Underweight | Putamen | Female | -0.026 | 0.018 | 0.915 |  |
| Underweight | Thalamus | Female | 0.034 | 0.023 | 0.915 |  |
| Underweight | Amygdala | Male | 0.012 | 0.021 | 0.915 |  |
| Underweight | Caudate | Male | -0.004 | 0.01 | 0.915 |  |
| Underweight | Hippocampus | Male | 0.01 | 0.011 | 0.915 |  |
| Underweight | Pallidum | Male | 0.021 | 0.027 | 0.915 |  |
| Underweight | Putamen | Male | -0.01 | 0.014 | 0.915 |  |
| Underweight | Thalamus | Male | 0.016 | 0.018 | 0.915 |  |
| Underweight | Accumbens | Male | -0.002 | 0.025 | 0.988 |  |
| Note. Sex-specific within-person slopes were estimated by refitting each sex moderation model (eTable 21) with each sex as the reference in turn. β is the standardized within-person coefficient within the indicated sex. P values are from Wald tests; FDR correction was applied within each group across 19 regions and both sexes (38 tests), and significance markers are based on unrounded *q* values. BMI, body mass index; CDC, Centers for Disease Control and Prevention; FDR, false discovery rate; SE, standard error; *q, FDR-corrected P value*. **q* < .05; ***q* < .01; ****q* < .001. | | | | | | |

| **eTable 23. Sensitivity Analysis: Within-Person Estimates With Random Slope for Age, by CDC Weight Status Group.** | | | | | | |
| --- | --- | --- | --- | --- | --- | --- |
| group | region | β primary | q primary | β rs | q rs | Δ |
| Healthy weight | Middle temporal | 0.03 | 0 | 0.032 | 0 | 0.002 |
| Healthy weight | Lateral occipital | 0.036 | 0 | 0.037 | 0 | 0.001 |
| Healthy weight | Fusiform | 0.02 | 0.026 | 0.021 | 0.022 | 0.001 |
| Healthy weight | Inferior temporal | 0.014 | 0.133 | 0.017 | 0.068 | 0.003 |
| Healthy weight | Temporal pole | -0.018 | 0.231 | -0.016 | 0.272 | 0.002 |
| Healthy weight | Rostral mid-frontal | -0.011 | 0.287 | -0.011 | 0.292 | 0 |
| Healthy weight | Medial OFC | 0.011 | 0.357 | 0.012 | 0.292 | 0.001 |
| Healthy weight | Caudal ACC | 0.008 | 0.357 | 0.008 | 0.309 | 0 |
| Healthy weight | Rostral ACC | 0.01 | 0.377 | 0.01 | 0.347 | 0 |
| Healthy weight | Superior frontal | -0.008 | 0.377 | -0.009 | 0.347 | -0.001 |
| Healthy weight | Lateral OFC | -0.007 | 0.462 | -0.006 | 0.556 | 0.001 |
| Healthy weight | Insula | 0.005 | 0.651 | 0.006 | 0.611 | 0.001 |
| Healthy weight | Hippocampus | 0.021 | 0 | 0.022 | 0 | 0.001 |
| Healthy weight | Thalamus | 0.022 | 0 | 0.022 | 0 | 0 |
| Healthy weight | Putamen | 0.013 | 0.004 | 0.013 | 0.005 | 0 |
| Healthy weight | Pallidum | 0.02 | 0.027 | 0.019 | 0.04 | -0.001 |
| Healthy weight | Caudate | 0.004 | 0.377 | 0.005 | 0.272 | 0.001 |
| Healthy weight | Amygdala | 0.005 | 0.462 | 0.003 | 0.694 | -0.002 |
| Healthy weight | Accumbens | 0 | 0.98 | 0.001 | 0.944 | 0.001 |
| Overweight | Temporal pole | -0.076 | 0.095 | -0.075 | 0.222 | 0.001 |
| Overweight | Medial OFC | -0.04 | 0.226 | -0.037 | 0.327 | 0.003 |
| Overweight | Rostral ACC | -0.045 | 0.226 | -0.037 | 0.327 | 0.008 |
| Overweight | Caudal ACC | -0.027 | 0.226 | -0.025 | 0.327 | 0.002 |
| Overweight | Superior frontal | -0.041 | 0.226 | -0.037 | 0.327 | 0.004 |
| Overweight | Rostral mid-frontal | -0.037 | 0.226 | -0.032 | 0.327 | 0.005 |
| Overweight | Lateral OFC | -0.032 | 0.325 | -0.028 | 0.452 | 0.004 |
| Overweight | Middle temporal | 0.018 | 0.678 | 0.013 | 0.793 | -0.005 |
| Overweight | Insula | -0.016 | 0.779 | -0.01 | 0.793 | 0.006 |
| Overweight | Inferior temporal | 0.007 | 0.792 | 0.008 | 0.793 | 0.001 |
| Overweight | Lateral occipital | 0.006 | 0.792 | 0.007 | 0.793 | 0.001 |
| Overweight | Fusiform | -0.003 | 0.896 | -0.002 | 0.905 | 0.001 |
| Overweight | Thalamus | 0.038 | 0.095 | 0.032 | 0.226 | -0.006 |
| Overweight | Hippocampus | 0.016 | 0.226 | 0.015 | 0.327 | -0.001 |
| Overweight | Amygdala | -0.013 | 0.678 | -0.016 | 0.582 | -0.003 |
| Overweight | Pallidum | -0.012 | 0.779 | -0.012 | 0.793 | 0 |
| Overweight | Accumbens | -0.009 | 0.788 | -0.01 | 0.793 | -0.001 |
| Overweight | Putamen | 0.005 | 0.788 | 0.003 | 0.82 | -0.002 |
| Overweight | Caudate | 0.003 | 0.792 | -0.004 | 0.793 | -0.007 |
| Obese | Insula | -0.086 | 0.053 | -0.093 | 0.025 | -0.007 |
| Obese | Temporal pole | -0.098 | 0.075 | -0.097 | 0.08 | 0.001 |
| Obese | Lateral OFC | -0.045 | 0.277 | -0.044 | 0.283 | 0.001 |
| Obese | Rostral ACC | -0.043 | 0.277 | -0.044 | 0.283 | -0.001 |
| Obese | Inferior temporal | -0.044 | 0.277 | -0.046 | 0.283 | -0.002 |
| Obese | Rostral mid-frontal | 0.035 | 0.299 | 0.035 | 0.295 | 0 |
| Obese | Fusiform | -0.035 | 0.299 | -0.036 | 0.295 | -0.001 |
| Obese | Lateral occipital | -0.024 | 0.44 | -0.027 | 0.344 | -0.003 |
| Obese | Medial OFC | 0.02 | 0.58 | 0.019 | 0.599 | -0.001 |
| Obese | Caudal ACC | -0.014 | 0.58 | -0.016 | 0.525 | -0.002 |
| Obese | Superior frontal | -0.016 | 0.609 | -0.019 | 0.572 | -0.003 |
| Obese | Middle temporal | -0.004 | 0.872 | -0.005 | 0.828 | -0.001 |
| Obese | Caudate | -0.019 | 0.26 | -0.018 | 0.283 | 0.001 |
| Obese | Putamen | -0.02 | 0.277 | -0.019 | 0.283 | 0.001 |
| Obese | Amygdala | -0.026 | 0.299 | -0.025 | 0.295 | 0.001 |
| Obese | Thalamus | 0.022 | 0.299 | 0.028 | 0.283 | 0.006 |
| Obese | Hippocampus | 0.009 | 0.58 | 0.007 | 0.63 | -0.002 |
| Obese | Accumbens | -0.011 | 0.681 | -0.013 | 0.63 | -0.002 |
| Obese | Pallidum | 0.006 | 0.845 | 0.007 | 0.818 | 0.001 |
| Underweight | Medial OFC | 0.03 | 0.778 | 0.032 | 0.752 | 0.002 |
| Underweight | Fusiform | -0.027 | 0.778 | -0.027 | 0.752 | 0 |
| Underweight | Lateral OFC | 0.02 | 0.876 | 0.021 | 0.863 | 0.001 |
| Underweight | Temporal pole | 0.025 | 0.876 | 0.024 | 0.877 | -0.001 |
| Underweight | Rostral ACC | 0.02 | 0.876 | 0.029 | 0.752 | 0.009 |
| Underweight | Caudal ACC | 0.003 | 0.97 | 0.004 | 0.949 | 0.001 |
| Underweight | Superior frontal | 0.004 | 0.97 | 0.004 | 0.949 | 0 |
| Underweight | Rostral mid-frontal | -0.009 | 0.97 | -0.01 | 0.949 | -0.001 |
| Underweight | Insula | -0.01 | 0.97 | -0.01 | 0.949 | 0 |
| Underweight | Inferior temporal | 0.003 | 0.97 | 0.003 | 0.949 | 0 |
| Underweight | Middle temporal | -0.009 | 0.97 | -0.009 | 0.949 | 0 |
| Underweight | Lateral occipital | -0.004 | 0.97 | -0.004 | 0.949 | 0 |
| Underweight | Pallidum | 0.03 | 0.778 | 0.026 | 0.752 | -0.004 |
| Underweight | Putamen | -0.016 | 0.778 | -0.016 | 0.752 | 0 |
| Underweight | Thalamus | 0.022 | 0.778 | 0.025 | 0.752 | 0.003 |
| Underweight | Accumbens | -0.02 | 0.876 | -0.02 | 0.862 | 0 |
| Underweight | Amygdala | 0.004 | 0.97 | 0.003 | 0.949 | -0.001 |
| Underweight | Caudate | 0 | 0.986 | -0.002 | 0.949 | -0.002 |
| Underweight | Hippocampus | 0 | 0.986 | 0 | 0.966 | 0 |
| Note. Primary within-person estimates were refitted with an added participant-level random slope for age; all other model terms were unchanged. Columns show the primary estimate and its *q* value, the random-slope estimate and its *q* value, and their difference (Δ, random-slope minus primary estimate). P values are from Wald tests; FDR correction was applied within each group across 19 regions. BMI, body mass index; CDC, Centers for Disease Control and Prevention; FDR, false discovery rate; *q, FDR-corrected P value*. | | | | | | |

| **eTable 24. Sensitivity Analysis: Within-Person Estimates After Excluding Biologically Implausible BMI Values, by CDC Weight Status Group.** | | | | | | |
| --- | --- | --- | --- | --- | --- | --- |
| group | region | β primary | q primary | β biv | q biv | Δ |
| Healthy weight | Middle temporal | 0.03 | 0 | 0.03 | 0 | 0 |
| Healthy weight | Lateral occipital | 0.036 | 0 | 0.036 | 0 | 0 |
| Healthy weight | Fusiform | 0.02 | 0.026 | 0.02 | 0.026 | 0 |
| Healthy weight | Inferior temporal | 0.014 | 0.133 | 0.014 | 0.133 | 0 |
| Healthy weight | Temporal pole | -0.018 | 0.231 | -0.018 | 0.231 | 0 |
| Healthy weight | Rostral mid-frontal | -0.011 | 0.287 | -0.011 | 0.287 | 0 |
| Healthy weight | Medial OFC | 0.011 | 0.357 | 0.011 | 0.357 | 0 |
| Healthy weight | Caudal ACC | 0.008 | 0.357 | 0.008 | 0.357 | 0 |
| Healthy weight | Rostral ACC | 0.01 | 0.377 | 0.01 | 0.377 | 0 |
| Healthy weight | Superior frontal | -0.008 | 0.377 | -0.008 | 0.377 | 0 |
| Healthy weight | Lateral OFC | -0.007 | 0.462 | -0.007 | 0.462 | 0 |
| Healthy weight | Insula | 0.005 | 0.651 | 0.005 | 0.651 | 0 |
| Healthy weight | Hippocampus | 0.021 | 0 | 0.021 | 0 | 0 |
| Healthy weight | Thalamus | 0.022 | 0 | 0.022 | 0 | 0 |
| Healthy weight | Putamen | 0.013 | 0.004 | 0.013 | 0.004 | 0 |
| Healthy weight | Pallidum | 0.02 | 0.027 | 0.02 | 0.027 | 0 |
| Healthy weight | Caudate | 0.004 | 0.377 | 0.004 | 0.377 | 0 |
| Healthy weight | Amygdala | 0.005 | 0.462 | 0.005 | 0.462 | 0 |
| Healthy weight | Accumbens | 0 | 0.98 | 0 | 0.98 | 0 |
| Overweight | Temporal pole | -0.076 | 0.095 | -0.076 | 0.095 | 0 |
| Overweight | Medial OFC | -0.04 | 0.226 | -0.04 | 0.226 | 0 |
| Overweight | Rostral ACC | -0.045 | 0.226 | -0.045 | 0.226 | 0 |
| Overweight | Caudal ACC | -0.027 | 0.226 | -0.027 | 0.226 | 0 |
| Overweight | Superior frontal | -0.041 | 0.226 | -0.041 | 0.226 | 0 |
| Overweight | Rostral mid-frontal | -0.037 | 0.226 | -0.037 | 0.226 | 0 |
| Overweight | Lateral OFC | -0.032 | 0.325 | -0.032 | 0.325 | 0 |
| Overweight | Middle temporal | 0.018 | 0.678 | 0.018 | 0.678 | 0 |
| Overweight | Insula | -0.016 | 0.779 | -0.016 | 0.779 | 0 |
| Overweight | Inferior temporal | 0.007 | 0.792 | 0.007 | 0.792 | 0 |
| Overweight | Lateral occipital | 0.006 | 0.792 | 0.006 | 0.792 | 0 |
| Overweight | Fusiform | -0.003 | 0.896 | -0.003 | 0.896 | 0 |
| Overweight | Thalamus | 0.038 | 0.095 | 0.038 | 0.095 | 0 |
| Overweight | Hippocampus | 0.016 | 0.226 | 0.016 | 0.226 | 0 |
| Overweight | Amygdala | -0.013 | 0.678 | -0.013 | 0.678 | 0 |
| Overweight | Pallidum | -0.012 | 0.779 | -0.012 | 0.779 | 0 |
| Overweight | Accumbens | -0.009 | 0.788 | -0.009 | 0.788 | 0 |
| Overweight | Putamen | 0.005 | 0.788 | 0.005 | 0.788 | 0 |
| Overweight | Caudate | 0.003 | 0.792 | 0.003 | 0.792 | 0 |
| Obese | Insula | -0.086 | 0.053 | -0.093 | 0.024 | -0.007 |
| Obese | Temporal pole | -0.098 | 0.075 | -0.112 | 0.024 | -0.014 |
| Obese | Lateral OFC | -0.045 | 0.277 | -0.055 | 0.202 | -0.01 |
| Obese | Rostral ACC | -0.043 | 0.277 | -0.048 | 0.216 | -0.005 |
| Obese | Inferior temporal | -0.044 | 0.277 | -0.049 | 0.202 | -0.005 |
| Obese | Rostral mid-frontal | 0.035 | 0.299 | 0.034 | 0.325 | -0.001 |
| Obese | Fusiform | -0.035 | 0.299 | -0.041 | 0.232 | -0.006 |
| Obese | Lateral occipital | -0.024 | 0.44 | -0.028 | 0.342 | -0.004 |
| Obese | Medial OFC | 0.02 | 0.58 | 0.013 | 0.696 | -0.007 |
| Obese | Caudal ACC | -0.014 | 0.58 | -0.012 | 0.652 | 0.002 |
| Obese | Superior frontal | -0.016 | 0.609 | -0.016 | 0.652 | 0 |
| Obese | Middle temporal | -0.004 | 0.872 | -0.008 | 0.73 | -0.004 |
| Obese | Caudate | -0.019 | 0.26 | -0.021 | 0.191 | -0.002 |
| Obese | Putamen | -0.02 | 0.277 | -0.021 | 0.216 | -0.001 |
| Obese | Amygdala | -0.026 | 0.299 | -0.03 | 0.216 | -0.004 |
| Obese | Thalamus | 0.022 | 0.299 | 0.021 | 0.325 | -0.001 |
| Obese | Hippocampus | 0.009 | 0.58 | 0.009 | 0.609 | 0 |
| Obese | Accumbens | -0.011 | 0.681 | -0.011 | 0.696 | 0 |
| Obese | Pallidum | 0.006 | 0.845 | 0.008 | 0.73 | 0.002 |
| Underweight | Medial OFC | 0.03 | 0.778 | 0.043 | 0.758 | 0.013 |
| Underweight | Fusiform | -0.027 | 0.778 | -0.026 | 0.794 | 0.001 |
| Underweight | Lateral OFC | 0.02 | 0.876 | 0.017 | 0.858 | -0.003 |
| Underweight | Temporal pole | 0.025 | 0.876 | 0.012 | 0.858 | -0.013 |
| Underweight | Rostral ACC | 0.02 | 0.876 | 0.041 | 0.758 | 0.021 |
| Underweight | Caudal ACC | 0.003 | 0.97 | 0.006 | 0.858 | 0.003 |
| Underweight | Superior frontal | 0.004 | 0.97 | 0.007 | 0.858 | 0.003 |
| Underweight | Rostral mid-frontal | -0.009 | 0.97 | -0.008 | 0.858 | 0.001 |
| Underweight | Insula | -0.01 | 0.97 | 0 | 0.997 | 0.01 |
| Underweight | Inferior temporal | 0.003 | 0.97 | 0.012 | 0.858 | 0.009 |
| Underweight | Middle temporal | -0.009 | 0.97 | -0.007 | 0.858 | 0.002 |
| Underweight | Lateral occipital | -0.004 | 0.97 | -0.013 | 0.858 | -0.009 |
| Underweight | Pallidum | 0.03 | 0.778 | 0.032 | 0.758 | 0.002 |
| Underweight | Putamen | -0.016 | 0.778 | -0.015 | 0.793 | 0.001 |
| Underweight | Thalamus | 0.022 | 0.778 | 0.026 | 0.758 | 0.004 |
| Underweight | Accumbens | -0.02 | 0.876 | -0.03 | 0.758 | -0.01 |
| Underweight | Amygdala | 0.004 | 0.97 | 0.011 | 0.858 | 0.007 |
| Underweight | Caudate | 0 | 0.986 | 0.005 | 0.858 | 0.005 |
| Underweight | Hippocampus | 0 | 0.986 | -0.001 | 0.968 | -0.001 |
| Note. Biologically implausible values were defined by the CDC modified BMI z-score (< −4 or > 8). In the analytic sample, 6 observations were flagged: 2 high values in the obesity group and 4 low values in the underweight group. No observations were flagged in the healthy weight or overweight groups, whose estimates are therefore unchanged. Flagged observations were excluded, within- and between-person components were recomputed, and group membership was kept as in the primary analysis. Columns show the primary estimate and its *q* value, the estimate after exclusion and its *q* value, and their difference (Δ, after exclusion minus primary estimate). P values are from Wald tests; FDR correction was applied within each group across 19 regions. BIV, biologically implausible value; BMI, body mass index; CDC, Centers for Disease Control and Prevention; FDR, false discovery rate; *q, FDR-corrected P value*. | | | | | | |

| **eTable 25. Model Diagnostics: Maximum Variance Inflation Factor and Residual Kurtosis by CDC Weight Status Group.** | | | |
| --- | --- | --- | --- |
| group | region | resid kurt | max VIF |
| Healthy weight | Middle temporal | 8.78 | 4.02 |
| Healthy weight | Inferior temporal | 5.92 | 4.02 |
| Healthy weight | Fusiform | 4.49 | 4.02 |
| Healthy weight | Temporal pole | 4.39 | 4.02 |
| Healthy weight | Lateral occipital | 2.94 | 4.02 |
| Healthy weight | Superior frontal | 2.93 | 4.02 |
| Healthy weight | Rostral mid-frontal | 2.46 | 4.02 |
| Healthy weight | Caudal ACC | 1.78 | 4.02 |
| Healthy weight | Rostral ACC | 1.73 | 4.02 |
| Healthy weight | Lateral OFC | 1.63 | 4.02 |
| Healthy weight | Insula | 1.03 | 4.02 |
| Healthy weight | Medial OFC | 0.96 | 4.02 |
| Healthy weight | Caudate | 31.46 | 4.02 |
| Healthy weight | Putamen | 7.16 | 4.02 |
| Healthy weight | Pallidum | 4.41 | 4.02 |
| Healthy weight | Hippocampus | 3.68 | 4.02 |
| Healthy weight | Accumbens | 1.82 | 4.02 |
| Healthy weight | Amygdala | 1.76 | 4.02 |
| Healthy weight | Thalamus | 1.2 | 4.02 |
| Overweight | Middle temporal | 4.93 | 3.62 |
| Overweight | Temporal pole | 4.19 | 3.62 |
| Overweight | Inferior temporal | 4 | 3.62 |
| Overweight | Fusiform | 3.91 | 3.62 |
| Overweight | Superior frontal | 3.09 | 3.62 |
| Overweight | Lateral occipital | 2.8 | 3.62 |
| Overweight | Rostral mid-frontal | 2.57 | 3.62 |
| Overweight | Caudal ACC | 2.03 | 3.62 |
| Overweight | Lateral OFC | 1.9 | 3.62 |
| Overweight | Medial OFC | 1.12 | 3.62 |
| Overweight | Rostral ACC | 1.1 | 3.62 |
| Overweight | Insula | 0.72 | 3.62 |
| Overweight | Caudate | 4.73 | 3.63 |
| Overweight | Putamen | 3.73 | 3.63 |
| Overweight | Pallidum | 3.29 | 3.63 |
| Overweight | Hippocampus | 2.34 | 3.63 |
| Overweight | Thalamus | 1.07 | 3.63 |
| Overweight | Amygdala | 0.87 | 3.63 |
| Overweight | Accumbens | 0.47 | 3.63 |
| Obese | Inferior temporal | 7.14 | 3.42 |
| Obese | Fusiform | 4.64 | 3.42 |
| Obese | Temporal pole | 4.06 | 3.42 |
| Obese | Middle temporal | 3.68 | 3.42 |
| Obese | Lateral OFC | 3.15 | 3.42 |
| Obese | Lateral occipital | 2.74 | 3.42 |
| Obese | Superior frontal | 2.28 | 3.42 |
| Obese | Rostral mid-frontal | 2.12 | 3.42 |
| Obese | Rostral ACC | 1.37 | 3.42 |
| Obese | Medial OFC | 1.23 | 3.42 |
| Obese | Caudal ACC | 1.17 | 3.42 |
| Obese | Insula | 0.77 | 3.42 |
| Obese | Caudate | 11.28 | 3.44 |
| Obese | Accumbens | 6.12 | 3.44 |
| Obese | Putamen | 5.35 | 3.44 |
| Obese | Hippocampus | 5.25 | 3.44 |
| Obese | Pallidum | 4.23 | 3.44 |
| Obese | Amygdala | 1.17 | 3.44 |
| Obese | Thalamus | 1.11 | 3.44 |
| Underweight | Middle temporal | 8.17 | 3.67 |
| Underweight | Fusiform | 8.09 | 3.67 |
| Underweight | Inferior temporal | 7.77 | 3.67 |
| Underweight | Temporal pole | 5.41 | 3.67 |
| Underweight | Lateral occipital | 1.9 | 3.67 |
| Underweight | Lateral OFC | 1.4 | 3.67 |
| Underweight | Superior frontal | 1.12 | 3.67 |
| Underweight | Rostral mid-frontal | 0.8 | 3.67 |
| Underweight | Medial OFC | 0.66 | 3.67 |
| Underweight | Caudal ACC | 0.65 | 3.67 |
| Underweight | Rostral ACC | 0.33 | 3.67 |
| Underweight | Insula | 0.3 | 3.67 |
| Underweight | Pallidum | 6.59 | 3.68 |
| Underweight | Putamen | 5.39 | 3.68 |
| Underweight | Caudate | 2.9 | 3.68 |
| Underweight | Amygdala | 1.93 | 3.68 |
| Underweight | Hippocampus | 0.87 | 3.68 |
| Underweight | Thalamus | 0.56 | 3.68 |
| Underweight | Accumbens | 0.04 | 3.68 |
| Note. Variance inflation factors (VIFs) were computed from linear-model approximations of the fixed effects of each primary within-between model. Because race and ethnicity has 4 df, its generalized VIF was converted to the VIF scale as (GVIF^(1/(2·df)))². The maximum VIF across model terms is shown; values below 5 indicate acceptable multicollinearity. The maximum VIF in the full range model was 3.82. Excess residual kurtosis was computed from normalized residuals of the primary mixed-effects models. BMI, body mass index; CDC, Centers for Disease Control and Prevention; GVIF, generalized variance inflation factor; VIF, variance inflation factor. | | | |

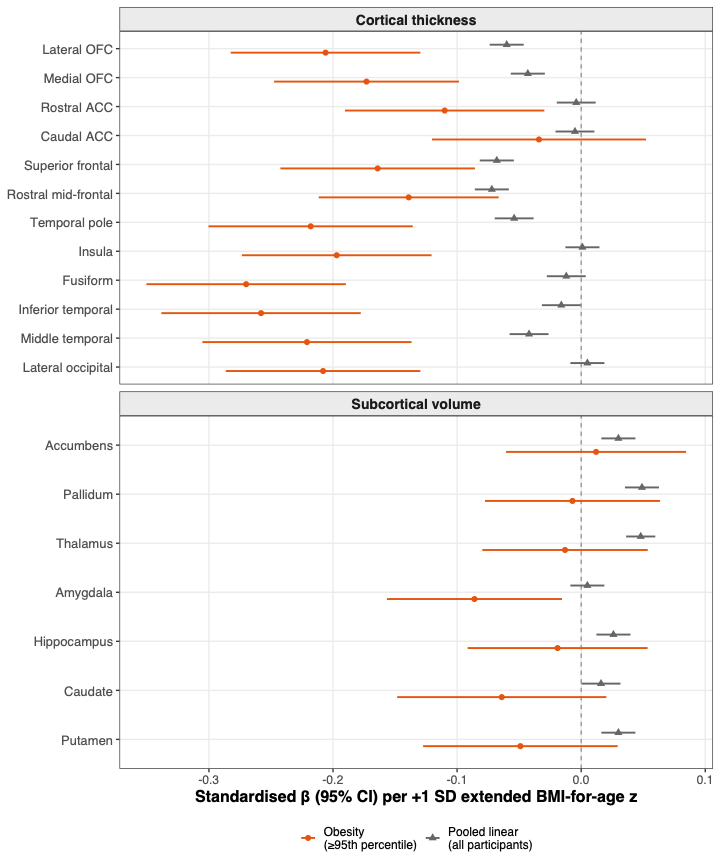

**eFigure 18.** Comparison of full-range (pooled) vs obesity-specific between-person BMI-for-age z associations with brain structure. Point-range forest plot of standardized between-person β coefficients (95% CIs) per 1-SD increase in extended BMI-for-age z for all 19 a priori regions (y-axis), comparing the obesity group–specific estimate (orange circles) with the pooled model estimate across all participants (gray triangles) for each region. The upper panel shows cortical thickness and the lower panel subcortical volume; regions are ordered anatomically, and the dashed vertical line indicates β = 0. Models were adjusted for age, age², sex, pubertal development, socioeconomic status, race and ethnicity, and intracranial volume (subcortical outcomes). The figure illustrates the attenuation of cortical thinning in the pooled model relative to the obesity-specific estimates (eg, fusiform β = −0.012 pooled vs −0.270 in obesity), and significant positive pooled estimates in 5 subcortical regions (accumbens, hippocampus, pallidum, putamen, and thalamus) that reflect the healthy weight group and are absent in the obesity group. Pooled estimates are in eTable 2; obesity-specific estimates are in eTable 17, Section A.

**eReferences**

1.	Freedman D. cdcanthro: Sex- and Age-Standardized Metrics from the Centers for Disease Control and Prevention (CDC) Growth Charts. Published online May 22, 2026. Accessed June 22, 2026. https://cran.r-project.org/web/packages/cdcanthro/index.html

2.	CDC. Growth Charts - CDC Extended BMI-for-Age Growth Charts. October 15, 2024. Accessed June 22, 2026. https://www.cdc.gov/growthcharts/extended-bmi.htm

3.	Freedman DS, Lawman HG, Pan L, et al. The prevalence and validity of high, biologically implausible values of weight, height, and BMI among 8.8 million children. *Obesity*. 2016;24(5):1132-1139. doi:10.1002/oby.21446

4.	Willette AA, Kapogiannis D. Does the brain shrink as the waist expands? *Ageing Res Rev*. 2015;20:86-97. doi:10.1016/j.arr.2014.03.007

5.	Gómez-Apo E, Mondragón-Maya A, Ferrari-Díaz M, Silva-Pereyra J. Structural Brain Changes Associated with Overweight and Obesity. *J Obes*. 2021;2021(1):6613385. doi:10.1155/2021/6613385

6.	Laurent JS, Watts R, Adise S, et al. Associations Among Body Mass Index, Cortical Thickness, and Executive Function in Children. *JAMA Pediatr*. 2020;174(2):170-177. doi:10.1001/jamapediatrics.2019.4708

7.	Ronan L, Alexander-Bloch AF, Wagstyl K, et al. Obesity associated with increased brain age from midlife. *Neurobiol Aging*. 2016;47:63-70. doi:10.1016/j.neurobiolaging.2016.07.010

8.	Kaltenhauser S, Weber CF, Lin H, et al. Association of Body Mass Index and Waist Circumference With Imaging Metrics of Brain Integrity and Functional Connectivity in Children Aged 9 to 10 Years in the US, 2016-2018. *JAMA Netw Open*. 2023;6(5):e2314193. doi:10.1001/jamanetworkopen.2023.14193

9.	Rapuano KM, Laurent JS, Hagler DJ, et al. Nucleus accumbens cytoarchitecture predicts weight gain in children. *Proc Natl Acad Sci*. 2020;117(43):26977-26984. doi:10.1073/pnas.2007918117

10.	de Groot CJ, van den Akker ELT, Rings EHHM, Delemarre-van de Waal HA, van der Grond J. Brain structure, executive function and appetitive traits in adolescent obesity. *Pediatr Obes*. 2017;12(4):e33-e36. doi:10.1111/ijpo.12149

11.	Perlaki G, Molnar D, Smeets PAM, et al. Volumetric gray matter measures of amygdala and accumbens in childhood overweight/obesity. *PLOS ONE*. 2018;13(10):e0205331. doi:10.1371/journal.pone.0205331

12.	Morys F, Tremblay C, Rahayel S, et al. Neural correlates of obesity across the lifespan. *Commun Biol*. 2024;7(1):656. doi:10.1038/s42003-024-06361-9

13.	Ronan L, Alexander-Bloch A, Fletcher PC. Childhood Obesity, Cortical Structure, and Executive Function in Healthy Children. *Cereb Cortex*. 2020;30(4):2519-2528. doi:10.1093/cercor/bhz257

14.	Cheng TW, Magis-Weinberg L, Guazzelli Williamson V, et al. A Researcher’s Guide to the Measurement and Modeling of Puberty in the ABCD Study® at Baseline. *Front Endocrinol*. 2021;12:471. doi:10.3389/fendo.2021.608575

15.	Beer JC, Tustison NJ, Cook PA, et al. Longitudinal ComBat: A method for harmonizing longitudinal multi-scanner imaging data. *NeuroImage*. 2020;220:117129. doi:10.1016/j.neuroimage.2020.117129
